# Estimating Disorder Probability Based on Polygenic Prediction Using the BPC Approach

**DOI:** 10.1101/2024.01.12.24301157

**Authors:** Emil Uffelmann, Major Depressive Disorder Working Group of the Psychiatric Genomics Consortium, Schizophrenia Working Group of the Psychiatric Genomics Consortium, Alkes L. Price, Danielle Posthuma, Wouter J. Peyrot

## Abstract

Polygenic Scores (PGSs) summarize an individual’s genetic propensity for a given trait in a single value based on SNP effect sizes derived from Genome-Wide Association Study (GWAS) results. Methods have been developed that apply Bayesian approaches to improve the prediction accuracy of PGSs through optimization of estimated effect sizes. While these methods are generally well-calibrated for continuous traits (implying the predicted values are, on average, equal to the true trait values), they are not well-calibrated for binary disorder traits in ascertained samples. This is a problem because well-calibrated PGSs are needed to reliably compute the absolute disorder probability for an individual to facilitate future clinical implementation. Here, we introduce the Bayesian polygenic score Probability Conversion (BPC) approach, which computes an individual’s predicted disorder probability using GWAS summary statistics, an existing Bayesian PGS method (e.g., PRScs, SBayesR), the individual’s genotype data, and a prior disorder probability (which can be specified flexibly, based on e.g., literature, small reference samples, or prior elicitation). The BPC approach transforms the PGS to its underlying liability scale, computes the variances of the PGS in cases and controls, and applies Bayes’ Theorem to compute the absolute disorder probability; it is practical in its application as it does not require a tuning sample with both genotype and phenotype data. We applied the BPC approach to extensive simulated data and empirical data of nine disorders. The BPC approach yielded well-calibrated results that were consistently better than the results of another recently published approach.

## Introduction

Polygenic Scores (PGSs)^1^ are per-individual estimates of the total contribution of common genetic variants to a trait or disorder liability based on SNP effect sizes (betas) from Genome-Wide Association Studies (GWAS)^2^. PGSs for several traits show increasing clinical potential that rivals that of conventional clinical predictors^3–6^. While summarizing an individual’s genetic risk for a disorder in a single value has the potential to be a simple and informative metric, PGS applications are limited because they are generally only interpretable at the group level. Accordingly, PGSs are commonly evaluated using the coefficient of determination (*R*^2^)^7^ or the Area Under the Curve (AUC)^8^, metrics that are blind to the scale of the PGS. Moreover, risk estimates based on PGSs are often reported in quantiles (e.g., a PGS falls in the top 5% of a given distribution), which can be challenging to interpret in terms of personal absolute risk of disease.

To make PGSs directly interpretable to individuals, they can be transformed into probabilities. For example, if an individual receives a PGS of 0.5 for multiple sclerosis, then this should correspond to a 50% probability of that individual developing multiple sclerosis in their lifetime. With access to a sufficiently large population-representative tuning sample with relevant pheno- and genotype data, such a transformation can be achieved with existing methods^9,10^. However, in most clinical settings, such samples are not readily available. Ideally, a single individual’s genotype data and publicly available resources should be sufficient to achieve such a transformation.

Bayesian PGS methods are known to be well-calibrated for continuous traits^11–13^, meaning the slope equals 1 when regressing the true phenotype on the PGS (implying the predicted values are, on average, equal to the true trait values). This offers a unique opportunity to achieve well-calibrated probabilities for binary disorder traits. However, when samples are over-ascertained for cases, Bayesian PGSs can become miscalibrated and, therefore, require a transformation.

Here, we introduce Bayesian polygenic score Probability Conversion (BPC), an approach to transform PGSs based on Bayesian methods (e.g. PRScs^12^ and SBayesR^11^), that only requires a single individual’s genotype data, GWAS summary statistics, and a prior disorder probability. We confirm that the resulting probabilities are well-calibrated in simulations and empirical analyses of nine disorders and that the BPC approach performs better than a recently published approach ^14^.

### Glossary

|  |  |
| --- | --- |
| Calibration | The degree to which a predicted value is equal to the true trait value on average. It is often assessed with the calibration slope for continuous traits<sup>13</sup> and with the ICI for disorder traits<sup>15</sup>. |
| Ascertainment | We use this term to refer to the oversampling of cases in genetic studies, compared to the lifetime population prevalence. |
| Prior disorder probability | The chance to be a case prior to genetic testing: equal to the lifetime population prevalence for a random individual (e.g. 1%), and much larger for help-seeking individuals (e.g. 50%). |
| Posterior mean beta | The SNP-effect size resulting from a Bayesian prediction method (e.g. PRScs and SBayesR). Polygenic risk score resulting from the effect sizes are well-calibrated for continuous traits<sup>11–13</sup>. |
| Effective sample size ($N_{eff}$) | The sample size with a case-control ratio of 0.5 that provides equivalent power as an observed study with a different case-control ratio<sup>16</sup>.<br>$N_{eff} = \frac{4}{\frac{1}{N_{case}} + \frac{1}{N_{control}}}$ |
| Liability | A latent continuous risk phenotype, modeled to underly binary disorder traits<sup>7,17,18</sup>. The liability is modeled following a standard normal distribution, and the threshold is defined such that the proportion of individuals exceeding the threshold is equal to the population lifetime prevalence. |
| $R^2$ | The variance explained in the phenotype by the PGS, which is also referred to as the coefficient of determination. $R^2$ has different values depending on the scale of the phenotype (e.g. observed scale with 50% case-control ascertainment vs. liability scale)<sup>7</sup>. |

## Methods

### Bayesian polygenic score Probability Conversion (BPC) approach

We developed the BPC approach to achieve calibration for binary disorder traits in ascertained samples, using the existing Bayesian Polygenic Score (PGS) methods PRScs^12^ and SBayesR^11^. The BPC approach follows four steps (see Figure 1).

**Figure 1.**
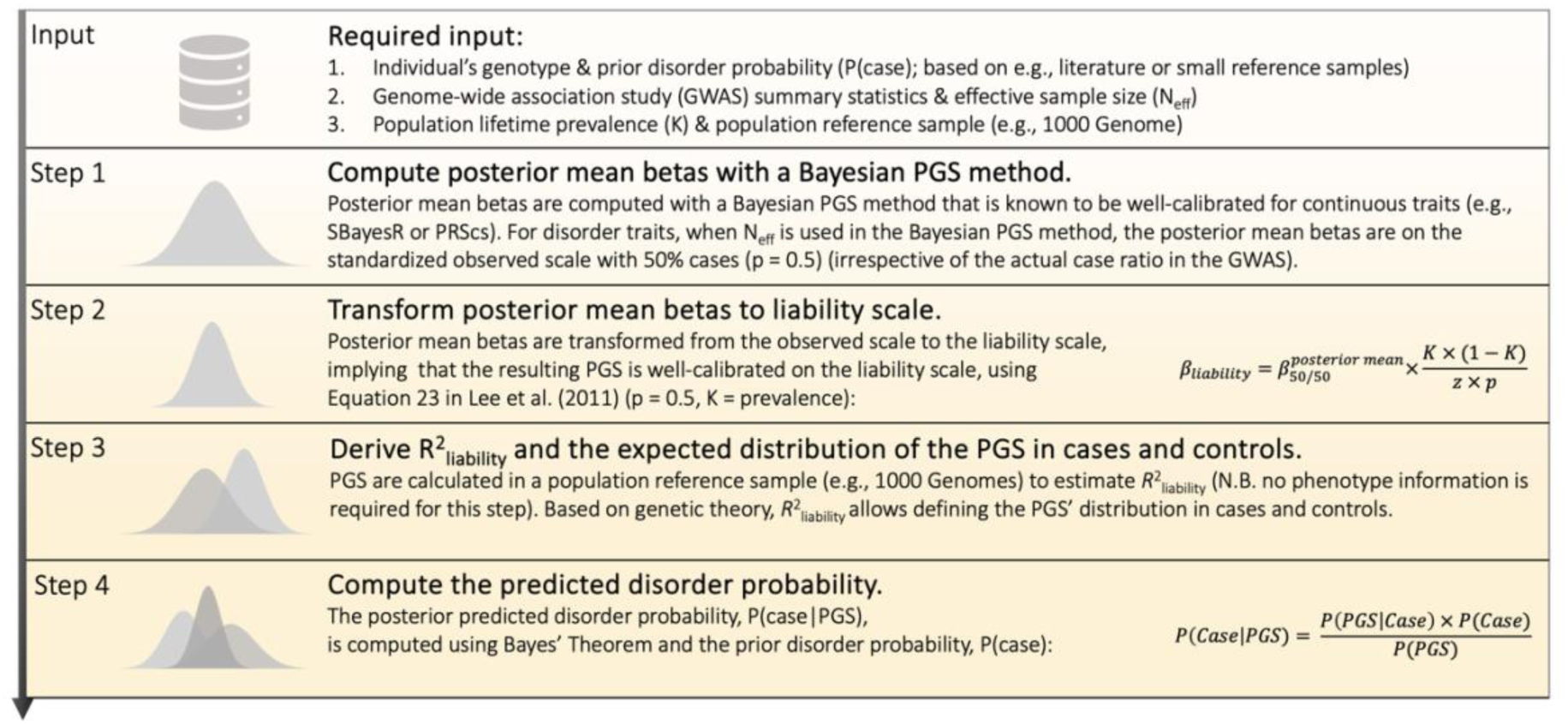
Overview of the Bayesian polygenic score Probability Conversion (BPC) approach. The BPC approach transforms an individual’s Polygenic Score (PGS) into a well-calibrated disorder probability. See the glossary for the definition of key terms.

#### Input

First, the BPC approach requires as input an individual’s genotype data and prior disorder probability. The prior can be based on context-specific prevalence estimates from published literature^19^, small reference samples, or prior elicitation (see **Discussion** for a detailed discussion on approaches to set the prior). For convenience, we mostly report results for a prior of 0.50. Second, the BPC approach requires the GWAS summary statistics (training sample) and the effective sample size (N_eff_, see Glossary and Supplementary Note 1) of the training sample (i.e., the sum of N_eff_ of all cohorts contributing to the meta-analysis^16^). The GWAS betas are assumed to be age-independent. Third, the population lifetime prevalence of the disorder of interest and an ancestry-matched population reference sample (e.g., 1000G) are required. No tuning sample with both genotype and phenotype data is required. We note that instead of an individual-level population reference sample, summary-level LD and allele frequency information could, in principle, be used as well. It is important to use the same set of SNPs across the training sample, reference sample, and the individual’s genotype data to ensure optimal prediction accuracy and well-calibrated BPC predictions.

#### Step 1 Compute posterior mean betas with a Bayesian PGS method

The BPC approach requires the posterior mean betas to be on the standardized observed scale with 50% case ascertainment (*p* = 0.5). For PRScs, this is achieved by simply using N_eff_ (i.e. the effective sample size)^16^ as input because PRScs is based on the GWAS Z-scores, noting that 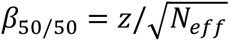 ^20^ (see Supplementary Note 2). (We note that, as long as N_eff_ is used, the proportion of cases in the discovery GWAS can have different values from 50%.) In contrast, SBayesR is based on the GWAS effect sizes (typically on the log-odds scale), which first need to be transformed to 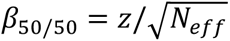 before applying SBayesR, while also setting N_eff_ as sample size.

#### Step 2 Transform posterior mean betas to liability scale

The posterior mean betas are transformed from the standardized observed scale with 50% case ascertainment to the continuous liability scale (*β*_*liability*_)^18^ (see Supplementary Note 3):

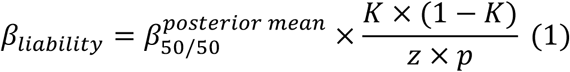

where *K* denotes the disorder population lifetime prevalence and *z* is the height of the standard normal probability density function at a threshold corresponding to *K*^18^. Subsequently, a PGS is constructed using *β*_*liability*_ and an individual’s genotype data.

#### Step 3 Derive R^2^_liability_ and the expected distribution of the PGS in cases and controls

To define the standard normal probability density function of the PGS in both cases and controls, an estimate of *R*^2^_liability_, the coefficient of determination on the liability scale^7^, is required. When a PGS is well-calibrated for a standardized phenotype with variance 1 (here the liability^21^), the variance of the PGS equals the variance explained by the PGS in the phenotype:

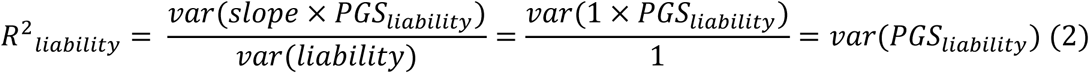

where *slope* refers to the regression of the liability on *PGS*_*liability*_ (which is equal to 1 due to the PGS being well-calibrated). Thus, *R*^2^_*liability*_ can be estimated by computing *var*(*PGS*_*liability*_) in an ancestry-matched population reference sample without the need for phenotype data. Given *R*^2^_*liability*_, the expected mean and variance of the PGS can be estimated in cases and in controls using normal theory^22,23^ (see Supplementary Note 4). Thus, the expected conditional probabilities *P*(*PGS*_*i*_|*D*_*i*_ = *case*) and *P*(*PGS*_*i*_|*D*_*i*_ = *control*) can be estimated for every individual *i* with PGS value *PGS*_*i*_ and disease status *D*_*i*_.

#### Step 4 Compute the genetically informed disorder probability

Finally, we use Bayes’ theorem to update the prior disorder probability to the posterior probability:

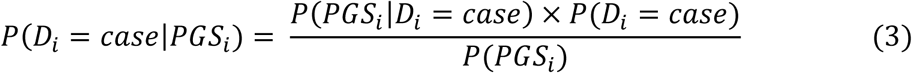

where *P*(*D*_*i*_ = *case*) is the prior disorder probability for individual *i*, *P*(*PGS*_*i*_|*D*_*i*_ = *case*) is the conditional probability, and *P*(*PGS*_*i*_) is the normalization factor corresponding to *P*(*PGS*_*i*_|*D*_*i*_ = *case*) × *P*(*D*_*i*_ = *case*) + *P*(*PGS*_*i*_|*D*_*i*_ = *control*) × (1 − *P*(*D*_*i*_ = *case*)). Thus, the BPC approach provides predicted disorder probabilities for individuals based on GWAS summary statistics, individual genotype data, and a prior disorder probability. (See **Code Availability** for R code to implement the BPC approach.). We note the prior disorder probability can be specified flexibly and does not depend on the case ratio in the training GWAS sample (see **Discussion** for a detailed discussion on how to set the prior).

### Alternative approaches to obtain disorder probabilities from PGS

The BPC approach transforms a single individual’s genotype data to the predicted disorder probability based on only publicly available data without requiring tuning samples that include both pheno- and genotype data, making it practical in its application. We are aware of only one other published approach that computes disorder probabilities only based on publicly available data, introduced in Pain et al. (2022)^14^. In addition, we describe the linear rescaling approach, an unpublished alternative to the BPC approach.

Briefly, the approach of Pain et al. (2022)^14^ works as follows. First, the difference in mean PGS between cases and controls is computed based on an estimate of the *R*^2^ (which is transformed to the AUC^24,25^), assuming the PGS have the same variance in cases and controls (scaled to 1). The *R*^2^ is estimated based on the GWAS summary statistics using lassosum^26^. Second, the PGS distribution across cases and controls is divided into quantiles, and third, the disorder probabilities per PGS quantile are assessed based on the testing sample’s case-control ratio (i.e. the prior disorder probability). For individual *i*, the predicted disorder probability follows by finding which quantile contains its PGS Z-value (standardized based on the distribution of the PGS in 1000 Genomes).

The approach of Pain et al. (2022) differs in three important ways from the BPC approach. First, it implicitly assumes that the variance and the mean of the PGS in the full population are the same as in the target sample. However, if the target sample is over-ascertained for cases, the variance and the mean are larger than in the full population (see Figure S1). As such, PGS Z-values based on the full population (i.e., 1000 Genomes) will overestimate the PGS Z-values in the ascertained target sample and, consequently, also the predicted disorder probabilities. Second, Pain et al. (2022) suggest using lassosum^26^ to estimate the *R*^2^ from summary statistics, while the BPC approach achieves this by estimating the variance of a well-calibrated PGS in a population reference sample (see **Methods**: *Step 3 Derive R^2^_liability_ and the expected distribution of the PGS in cases and controls*). Third, the Pain et al. (2022) approach assumes *var*(*PGS*|*case*) = *var*(*PGS*|*control*), while the BPC approach models more precisely the fact that *var*(*PGS*|*case*) < *var*(*PGS*|*control*), which has the most impact for disorders with low population lifetime prevalence (K) and large 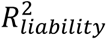 values (see **Results** & Table S1 for a summary of these differences).

We developed an alternative approach, the linear rescaling approach, to obtain well-calibrated predicted disorder probabilities, that does not apply Bayes’ Theorem but a linear rescaling of the *PGS*_*liability*_ instead. The linear rescaling approach follows steps 1-3 of the BPC approach described above and in Figure 1. Subsequently, the expected variance of the *PGS*_*liability*_ in the ascertained sample, *var*(*PGS*_*liability*_ | *ascertained sample*), is computed based on the prior disorder probability (i.e., the case-control ratio in the testing sample, *P*(*case*)) and the distribution of *PGS*_*liability*_in cases and controls (see **Methods**: *Step 3 Derive R^2^_liability_ and the expected distribution of the PGS in cases and controls*). Next, the PGS is scaled to PGS’ with the property that 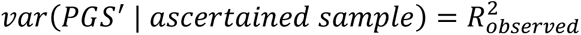 in the ascertained sample (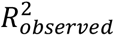 is computed based on *R*^2^_liability_ and the transformation introduced in Lee et al. (2012)^7^), resulting in *PGS*′ that is well-calibrated on the standardized observed scale (see Equation 2). Lastly, we scale the *PGS*′ (which is based on a standardized phenotype) to the observed scale with cases coded 1 and controls 0, 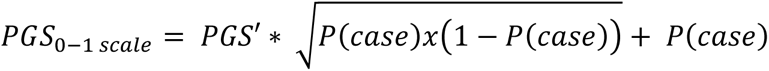, resulting in PGSs that represent the predicted disorder probability. We note the linear rescaling approach can lead to predicted disorder probabilities that are larger than 1 and smaller than 0, which we truncate to 1 and 0 before evaluating its calibration.

### Approaches using tuning samples

We developed an alternative BPC-tuned approach that is conceptually similar to the standard BPC approach outlined above. Instead of deriving them theoretically, it uses empirical estimates of the variances and means of the PGS in cases and controls derived from a tuning sample with both genotype and phenotype data. As such, the BPC-tuned approach skips steps 1 and 2 described above and in Figure 1.

The Logit-tuned approach, as applied in ref. ^10^, computes predicted disorder probabilities in three steps. First, the slope and intercept are estimated from a logistic regression model in the tuning sample: *D* ∼ *PGS*, where *D* ∈ {0,1} is a vector of binary disease status. Second, the PGSs in the testing sample are used to compute logit(*D̂*): *PGS* ∗ *slope* + *intercept*. Third, the predicted disorder probabilities are computed as the inverse logit transformation of 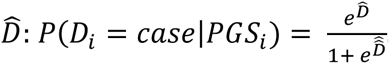

### Untransformed PGS

We also evaluated the calibration of untransformed PGSs. These are constructed using the posterior mean betas of step 1 (see Figure 1 and **Methods**: *Step 1 Compute posterior mean betas with a Bayesian PGS method*), which are on the standardized observed scale with 50% case ascertainment when N_eff_ is used as input in the Bayesian PGS methods. The resulting PGSs are centered around zero and cannot be interpreted as disorder probabilities.

### Metrics of performance

To assess calibration, we compute the Integrated Calibration Index (ICI): the weighted average of the absolute difference between the real and predicted disorder probability^15^. (The real disorder probability is computed using the loess smoothing function in R; thus, the ICI can be intuitively understood as the weighted difference between the calibration curve and the diagonal line in a calibration plot (see **Results**)). Lower values of the ICI indicate better calibration and perfect calibration implies ICI=0.

The calibration slope is another metric to assess calibration that is often used in the literature^11–13^, which refers to the slope from a linear regression of the phenotype of interest on the PGS. If the slope equals 1 and the intercept 0, the predictor is said to be well-calibrated. A downside of this metric is that a PGS with values outside the range of 0 and 1 can still have a calibration slope of 1, and the ICI has been proposed as a superior metric because the ICI is robust to sparse subregions of poor calibration^15^. Typically, untransformed Bayesian PGSs are centered around 0, and while they may have a calibration slope of 1, they cannot be interpreted as disorder probabilities and cannot be evaluated with the ICI.

To assess the prediction accuracy of the PGSs, we use the Area Under the Curve (AUC) and the coefficient of determination (*R*^2^) (we note the AUC and *R*^2^ can be transformed into one another^7^).

### Simulation analysis

We simulated individual-level data for 1,000 SNPs in Linkage Equilibrium based on the liability threshold model^17^ (see Supplementary Note 5 for details). We simulated a relatively small number of SNPs (M) because this allows the simulation of smaller training sample sizes (N), which reduces the computational cost. The PGS’s *R*^2^ primarily depends on 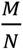, such that simulations at reduced values of both *M* and *N* are appropriate ^27^. To further reduce the computational cost, we did not simulate Linkage Disequilibrium (LD), which has no impact on the scale of the PGS as it aggregates all SNP effects into a single score. We repeated the simulations 100 times for eight different parameter settings where we varied the power of the training sample and thereby the coefficient of determination (*R*^2^) of the PGS (*R*^2^_*liability*_ = {0.01, 0.05, 0.10, 0.15}), as well as the disorder population lifetime prevalence (K = {0.01, 0.15}). The disorder’s SNP-based heritability was set to 0.2. We simulated three independent samples: a training sample with case-control information used to estimate SNP effects with a GWAS (varying N; see below), a population reference sample without case-control information to estimate *R*^2^_*liability*_ as described above (N = 503), and a testing sample with case control-information to evaluate model performance (N_case_=1,000 and N_control_=1,000). To achieve the desired *R*^2^_*liability*_in the testing sample, we approximated the required sample size of the training sample using the avengeme package in R^28^ (e.g. *N*_training_ = 2,759 when *R*^2^_*liability*_ = 0.1 and K = 0.01). We computed the posterior mean betas using Bpred, the version of LDPred that assumes linkage equilibrium^13^, with GWAS betas on the standardized observed scale with 50% case ascertainment and therefore used N_eff_ as input. We applied the BPC approach to estimate predicted disorder probabilities and compared it to the existing approach introduced in Pain et al. (2022)^14^.

### Empirical analysis

We analyzed nine phenotypes based on large training samples of GWAS meta-analyses, namely schizophrenia (SCZ)^29^, major depression (MD)^30^, breast cancer (BC)^31^, coronary artery disease (CAD; we note that 23% of the training sample included individuals from non-European populations)^32^, inflammatory bowel disease (IBD)^33^, multiple sclerosis (MS)^34^, prostate cancer (PC)^35^, rheumatoid arthritis (RA)^36^, and type 2 diabetes (T2D)^37^. We computed the PGSs in three testing samples that were fully independent of the respective training samples (Table 1). For SCZ and MD, 62 and 22 testing cohorts, respectively, were used, and PGSs were computed based on the GWAS results that excluded the testing cohort from the Psychiatric Genomics Consortium (PGC). In evaluating the ICI, we concatenated all individual cohorts. Testing data from the UK Biobank^38^ was used for BC, CAD, IBD, MS, PC, RA, and T2D. If SNP-wise N_eff_ values were available in the GWAS results, the maximum N_eff_ across all SNPs was used as input to the BPC approach (MD and SCZ). Alternatively, N_eff_ was calculated as the sum of N_eff_ of all contributing cohorts (CAD, IBD, MS, RA)^16^. If neither information was available, the SNP-wise N_eff_ were estimated analytically with 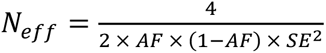^16^, where AF = effect allele frequency and SE = standard error (PC, BC). Because the analytically derived N_eff_ can produce large outliers, we used the 90^th^ percentile across all SNPs instead of the maximum as input to the BPC approach.

**Table 1.** Phenotype Summary. PGC-MD = Major Depression Working Group of the Psychiatric Genomics Consortium; PGC-SCZ = Schizophrenia Working Group of the Psychiatric Genomics Consortium; UKB = UK Biobank

| Phenotype | Abbreviation | Population lifetime prevalence | Training sample |  | Testing sample |  |
| --- | --- | --- | --- | --- | --- | --- |
| | | | Effective sample size<br>($N_{\text{case}}$ / $N_{\text{control}}$) | GWAS reference | Effective sample size*<br>($N_{\text{case}}$ / $N_{\text{control}}$) | Individual-level dataset |
| Major Depression | MD | 16.00%<sup>39</sup> | 133,299**<br>(50,968 / 96,399) | Wray et al.<br>(2018)<sup>30</sup> | 25,184***<br>(12,592 / 12,592) | PGC-MD<sup>30</sup> |
| Schizophrenia | SCZ | 1.00% | 115,996**<br>(48,650 / 70,612) | Trubetskoy et al.<br>(2022)<sup>29</sup> | 85,340***<br>(42,670 / 42,670) | PGC-SCZ<sup>29</sup> |
| Breast Cancer | BC | 12.50% | 231,040<br>(133,384 / 113,789) | Zhang et al.<br>(2020)<sup>31</sup> | 18,456<br>(9,228 / 9,228) | UKB<sup>38</sup> |
| Coronary Artery Disease | CAD | 3.00% | 129,014<br>(61,289 / 12,6310) | Nikpay et al.<br>(2015)<sup>32</sup> | 20,000<br>(10,000 / 10,000) | UKB<sup>38</sup> |
| Inflammatory Bowel Disease | IBD | 1.30% | 30,273<br>(12,924 / 21,770) | Liu et al.<br>(2015)<sup>33</sup> | 5,924<br>(2,962 / 2,962) | UKB<sup>38</sup> |
| Multiple Sclerosis | MS | 0.16% | 35,828<br>(14,802 / 26,703) | International Multiple Sclerosis Genetics Consortium<br>(2019)<sup>34</sup> | 2,368<br>(1,184 / 1,184) | UKB<sup>38</sup> |
| Prostate Cancer | PC | 12.50% | 125,417<br>(79,148 / 61,106) | Schumacher et al.<br>(2018)<sup>35</sup> | 7,026<br>(3,513 / 3,513) | UKB<sup>38</sup> |
| Rheumatoid Arthritis | RA | 0.50% | 58,012<br>(22,350 / 74,823) | Ishigaki et al.<br>(2022)<sup>36</sup> | 5,076<br>(2,538 / 2,538) | UKB<sup>38</sup> |
| Type 2 Diabetes | T2D | 5.00% | 158,261<br>(55,005 /<br>400,308) | Mahajan et<br>al. (2018b)<sup>37</sup> | 20,000<br>(10,000 /<br>10,000) | UKB<sup>38</sup> |

\* The effective testing sample size is reported for a testing sample case-control ratio of $P = 0.5$ . For analyses with testing sample case-control ratios of $P = 0.25$ and $0.75$ , cases and controls were down-sampled respectively.
\*\* The average effective sample size for leave-one-cohort-out GWASs is reported.
\*\*\* The total effective sample size across all cohorts is reported.

Standard quality control was applied: Ambiguous (i.e., A/T or C/G SNPs), duplicate, and mismatching alleles for SNPs across training, testing, and population reference sample were removed^1^; a minor allele frequency filter of 10%, and, when available, an imputation INFO filter of 0.9 was applied as described before^40^; The major histocompatibility complex (MHC) was removed (hg19 coordinates: 6:28000000:34000000).

Posterior mean betas of SNPs were computed with PRScs-auto10.1093/eurheartj/ehae649 (from here on simply referred to as PRScs; version June 4^th^, 2021) and SBayesR (version 2.03)^11^. PRScs uses a Linkage Disequilibrium (LD) reference panel based on HapMap3^41^ SNPs and Europeans from the 1000 Genomes Project^42^ (the default for PRScs). We use the default parameters listed on the software’s GitHub page (see **Web resources**). In the input of PRScs, we specified the sample size as N_eff_ to ensure posterior mean betas were on the standardized observed scale with 50% case ascertainment (see **Methods**: *Step 1 Compute posterior mean betas with a Bayesian PGS method*). SBayesR uses an LD reference panel that is based on HapMap3^41^ SNPs and 50,000 European UK Biobank subjects (the default for SBayesR version 2.03). In the input for SBayesR, we transformed the effect sizes to the standardized observed scale with 50% case ascertainment 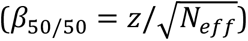 and set the sample size to N_eff_ (see **Methods**: *Step 1 Compute posterior mean betas with a Bayesian PGS method*).

To estimate *R*^2^_liability_ we use an ancestry-matched population reference sample, namely the European sample of 1000 Genomes^42^ (see **Methods**: *Derive R^2^_liability_ and the expected distribution of the PGS in cases and controls*), which we downloaded from the MAGMA website (see **Web resources**).

The posterior mean betas were used to compute the PGS in 1000 Genomes and in the testing sample with Plink1.9 (version Linux 64-bit 6^th^ June, 2021; command “--score <variant ID column> <effect allele column> <posterior mean beta> sum center”; see **Data and Code Availability**).

The BPC approach requires a valid estimate of the prior disorder probability, which we set to the case-control ratio in the testing sample (see **Discussion** for approaches to estimate the prior disorder probability). We ascertained cases in the testing sample such that the case-control ratio was equal to 25%, 50%, or 75%.

## Results

### Simulation analysis

We evaluated the BPC approach and Pain et al. (2022) across different values of *R*^2^_*liability*_ (1%, 5%, 10%, and 15%) and population lifetime prevalences (1% and 15%) in 100 simulation runs (see Figure 2). We used a simplified simulation setup with a relatively small number of causal SNPs in Linkage Equilibrium to limit computational costs (see **Methods: Simulation analyses**). Across all parameter combinations, the BPC approach consistently achieves mean ICI values close to 0 (ranging from mean 0.014 (± SE 0.0004) to 0.017 (± 0.0006) across 4×2=8 parameter settings), meaning the predicted and observed probabilities agree closely.

**Figure 2.**
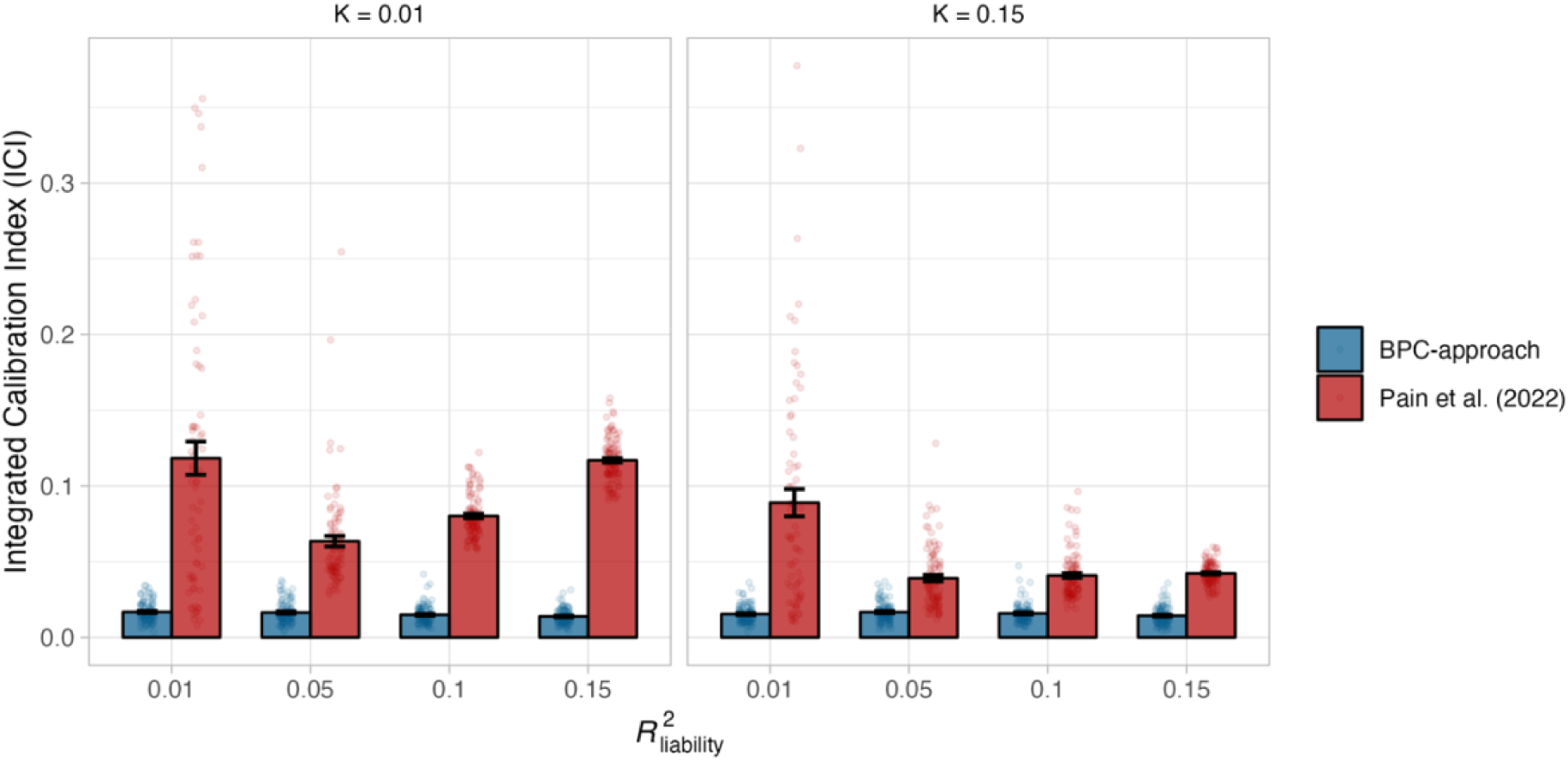
Calibration in simulations. Calibration of the BPC and the Pain et al. (2022) approach was evaluated using the Integrated Calibration Index (ICI) in 100 simulation runs and for combinations of two parameters, the population lifetime prevalence (K), and the explained variance of the PGS on the liability scale (*R*^2^_liability_). The BPC approach achieves low mean ICI values in every condition, while the mean ICI values of the Pain et al. (2022) approach are consistently larger. The difference between both approaches becomes larger for conditions with low population lifetime prevalences and large 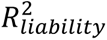 values. Error bars represent standard errors.

The Pain et al. (2022) approach performs considerably less well (ranging from 0.039 (± 0.002) to 0.118 (± 0.009) across all parameter settings; see Figure 2) because it does not distinguish the prior disorder probability (in this case, the testing sample case-control ratio) from the lifetime prevalence in the full population, which overestimates the predicted probabilities and negatively impacts calibration (see **Methods: Alternative approaches to obtain disorder probabilities from PGS** for details and Supplementary Figure 1 for a schematic representation). Indeed, the distinction between the BPC and Pain et al. (2022) approach is more pronounced when the disorder population lifetime prevalence is low because this increases the difference between the population lifetime prevalence and the prior disorder probability (which is set to 50%). Similarly, larger values of *R*^2^_*liability*_ exacerbate the overestimates of the Pain et al. (2022) approach because it leads to more power to detect the bias (except for *R*^2^_*liability*_ = 1%; see below). A simple adaptation of the Pain et al. (2022) approach to take both the population lifetime prevalence and prior disorder probability into account strongly improves its calibration and removes the negative impact of the low population lifetime prevalence and increasing *R*^2^_*liability*_ values; nevertheless, the BPC approach continues to achieve lower ICI values (see Supplementary Figure 2). For low simulated values of *R*^2^_*liability*_, when the GWAS has little power, the *R*^2^_*liability*_ values estimated with lassosum in the Pain et al. (2022) approach become unstable (see below), leading to an increased ICI. When we adjust the Pain et al. (2022) approach to take both the population lifetime prevalence and prior disorder probability into account and compute the variance of a well-calibrated PGS in a population reference sample to estimate *R*^2^_*liability*_ (instead of lassosum), the difference between both approaches becomes very small (see Supplementary Figure 3). Nonetheless, the BPC approach achieves slightly better calibration in nearly every condition, because the Pain et al. (2022) approach assumes that the variance of the PGS is the same in cases and controls while they are different. The difference becomes larger for higher *R*^2^_*liability*_ values and lower population lifetime prevalences (see Supplementary Figure 4 and **Methods**: **Alternative approaches to obtain disorder probabilities from PGS**).

We verified that doubling the number of causal SNPs does not affect these results, and the ICI of the BPC approach remains low (0.016 ± 0.008; *R*^2^_*liability*_= 0.05 and *K* = 0.01).

In addition to the ICI, we used the calibration slope and intercept to evaluate calibration. Again, the BPC approach consistently achieves good calibration (see Supplementary Figures 5 and 6) and performs better than the Pain et al. (2022) approach. Furthermore, the Pain et al. (2021) approach consistently overestimates the disorder probabilities, with slopes smaller than one and/or intercepts smaller than zero (see **Methods: Alternative approaches to obtain disorder probabilities from PGS**). In line with observations made in ^15^, we show that the ICI is a more stable metric of calibration, especially at small values of 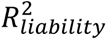 (see Supplementary Figure 7).

We also evaluated our linear rescaling approach (see **Methods**: **Alternative approaches to obtain disorder probabilities from PGS**). We found that the linear rescaling approach performs reasonably well but worse than the BPC approach because it can result in probabilities larger than 1 and lower than 0. This mostly occurs in conditions where the population lifetime prevalence is low and 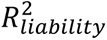 is large. Setting these outlying values to 1 and 0, respectively, negatively impacts calibration (see Supplementary Figure 8). Therefore, our primary recommendation is to use the BPC approach.

We found that the calibration slopes of untransformed Bayesian PGSs for binary disorder traits deviate from 1 in ascertained samples, even when the case-control ratios in the training and testing sample are both 50% and the PGSs are on the standardized observed scale with 50% case ascertainment. Similarly, the calibration intercepts deviate from 0 (see Supplementary Figures 9 and 10; the bias is most apparent when the population lifetime prevalence is low and 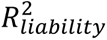 is large). This is because the transformation from the liability to the observed scale in ascertained samples is linear for the GWAS results (i.e., betas) used to compute the PGS^18^ but non-linear for the coefficient of determination (*R*^2^) of the PGS^7^ (see Supplementary Figure 11). As a result, *var*(*PGS*_*observed*_) and 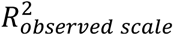 are not proportional, and the PGSs can thus not be well-calibrated (see equation 2) without a probability conversion approach. Untransformed PGS do attain accurate calibration when neither the training nor the testing sample case-control ratios differ from the population lifetime prevalence (i.e., random ascertainment), even when the population lifetime prevalence is low (K = 0.01) and 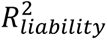 is large (0.15). The PGS’s mean calibration slope over 100 simulation runs does not significantly differ from 1 (mean calibration slope = 1.02, s.e.m. = 0.02). We note that the untransformed Bayesian PGSs are centered around zero and cannot be evaluated with the ICI^15^.

The BPC approach assumes that the PGSs are normally distributed in cases and controls. We verified that this assumption holds for all parameters in our simulations and that significant deviations are only observed at current unrealistically large values of 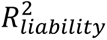 (≥ 0.6; see Supplementary Figures 12). A second assumption is that the liability conversion of the PGS is successful. We verified that regressing the liability scores on the PGSs (based on Bpred, a version of LDPred that assumes linkage equilibrium^13^) in a population reference sample leads to slopes and intercepts that are, on average, 1 and 0, respectively (see Supplementary Figure 13).

Lastly, we investigated the distribution of 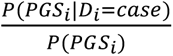 (see equation 3) to test how strongly the posterior predicted disorder probabilities depend on the prior (*P*(*D*_*i*_ = *case*)). If the probabilities are determined mainly by the prior, the distribution is expected to vary closely around 1. We find that the distributions vary markedly around 1 for most realistic simulation conditions (e.g., S.D. = 0.3 for K = 0.01, *R*^2^_liability_ = 0.05, and prior = 0.50), except when the 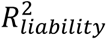 is very low, the population prevalence is high, and the prior is very high (i.e., S.D. = 0.05 for *R*^2^_liability_ = 0.01, K = 0.15, and prior = 0.75) (Supplementary Figure 14).

### Empirical analysis

To further evaluate the performance of the BPC approach, we applied it to nine phenotypes across nine training samples (SCZ^29^, MD^30^, BC^31^, CAD^32^, IBD^33^, MS^34^, PC^35^, RA^36^, and T2D^37^) and three testing samples (i.e., UK Biobank^38^, PGC-SCZ^29^, PGC-MD^30^; see **Methods: Empirical analysis** and Table 1 for a summary). We ascertained cases and controls for each phenotype such that the testing sample case-control ratios were 0.25, 0.5, and 0.75, thus testing the calibration of the BPC approach across a range of prior disorder probabilities. We performed similar comparisons as in the simulations with the addition of two applications of the BPC approach, one using PRScs^12^ (BPC-PRScs) and one using SBayesR^11^ (BPC-SBayesR) to compute posterior mean betas (see Figure 1 and **Methods: Bayesian polygenic score Probability Conversion (BPC) approach**). We note that for SBayesR, the results did not converge for prostate cancer and therefore depict one fewer data point. Results are reported in Figure 3 and Supplementary Table 2. Averaged across all prior disorder probabilities, BPC-PRScs achieves the lowest mean ICI value of 0.024 (± 0.002), followed by BPC-SBayesR with 0.034 (± 0.004). The Pain et al. (2022) approach has the largest mean ICI value of 0.053 (± 0.007). The BPC-PRScs approach consistently achieves the lowest mean ICI values across all prior disorder probabilities. We note the Pain et al. (2022) approach can be used with both PRScs and SBayesR. While the presented results are based on PRScs, using SBayesR yields comparable results (see Supplementary Figure 15 and Supplementary Table 2). The observation that the BPC approach produces well-calibrated predicted disorder probabilities suggests that the PGSs are also well-calibrated on the unobserved liability scale.

**Figure 3.**
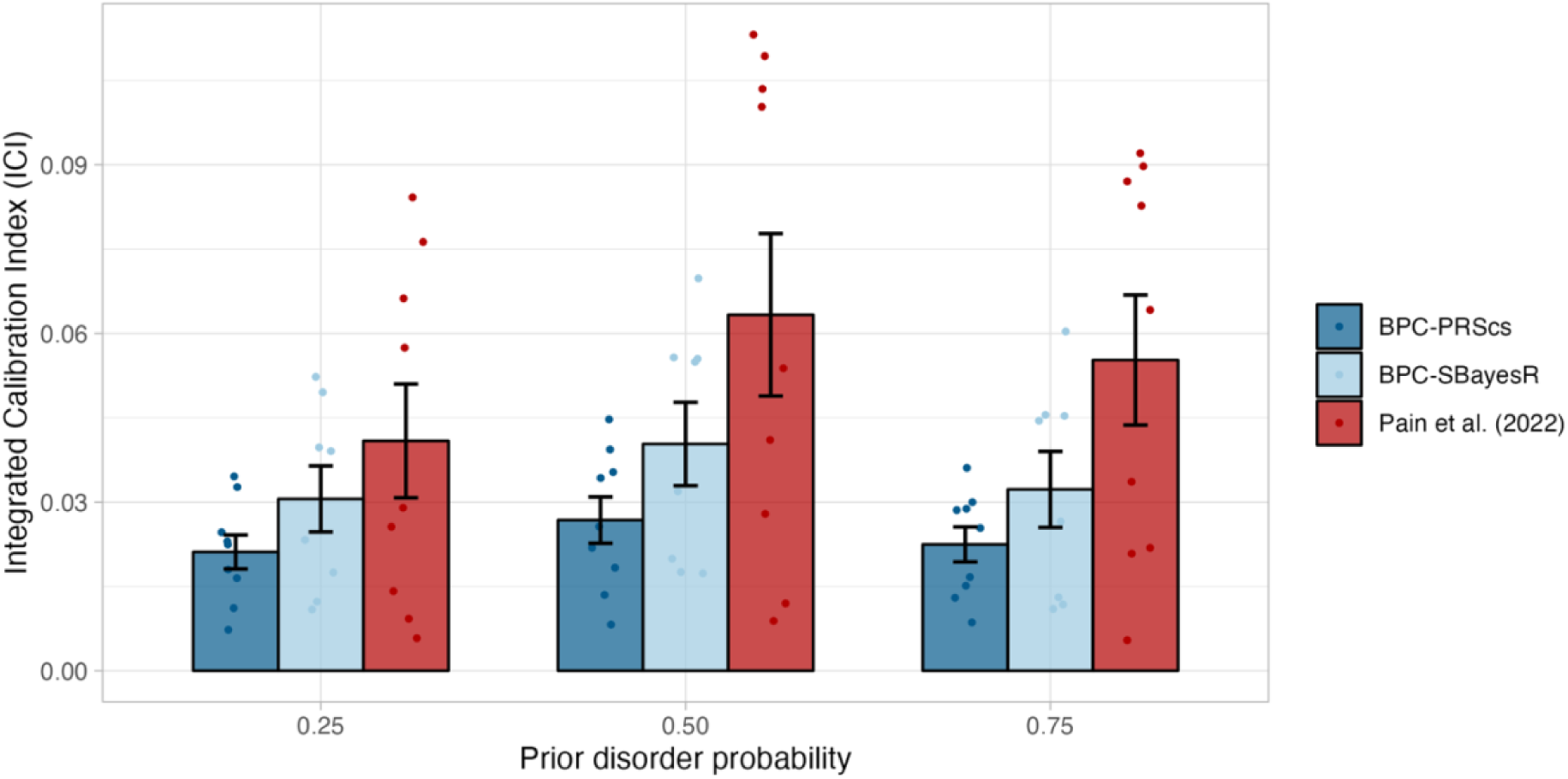
Calibration in empirical analyses of nine disorders. Calibration of the BPC and the Pain et al. (2022) approach was evaluated using the Integrated Calibration Index (ICI) for nine disorders, while varying the prior disorder probability. The BPC approach was applied using two Bayesian PGS methods, PRScs (BPC-PRScs) and SBayesR (BPC-SBayesR). The BPC-PRScs approach achieves the lowest mean ICI values across all prior disorder probabilities. BPC-SBayesR shows one fewer data points, as it did not converge for prostate cancer. Numerical values are presented in Table S2. Error bars represent standard errors.

When focusing in detail on the calibration plots with a prior disorder probability of 50%, BPC-PRScs shows better calibration than the Pain et al. (2022) approach for every trait, except Type 2 Diabetes (see Figure 4 and Supplementary Table 2). The Pain et al. (2022) approach tends to overestimate the probabilities for many traits, as can be seen by the right shift of the histograms and calibration lines. This is particularly true for traits with low population lifetime prevalence and large 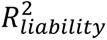 values, such as rare auto-immune disorders (i.e., Inflammatory Bowel Disorder, Multiple Sclerosis, and Rheumatoid Arthritis) and Prostate Cancer, which is in line with our theoretical expectations (see **Methods: Alternative approaches to obtain disorder probabilities from PGS** and Supplementary Figure 1 for a schematic representation).

**Figure 4.**
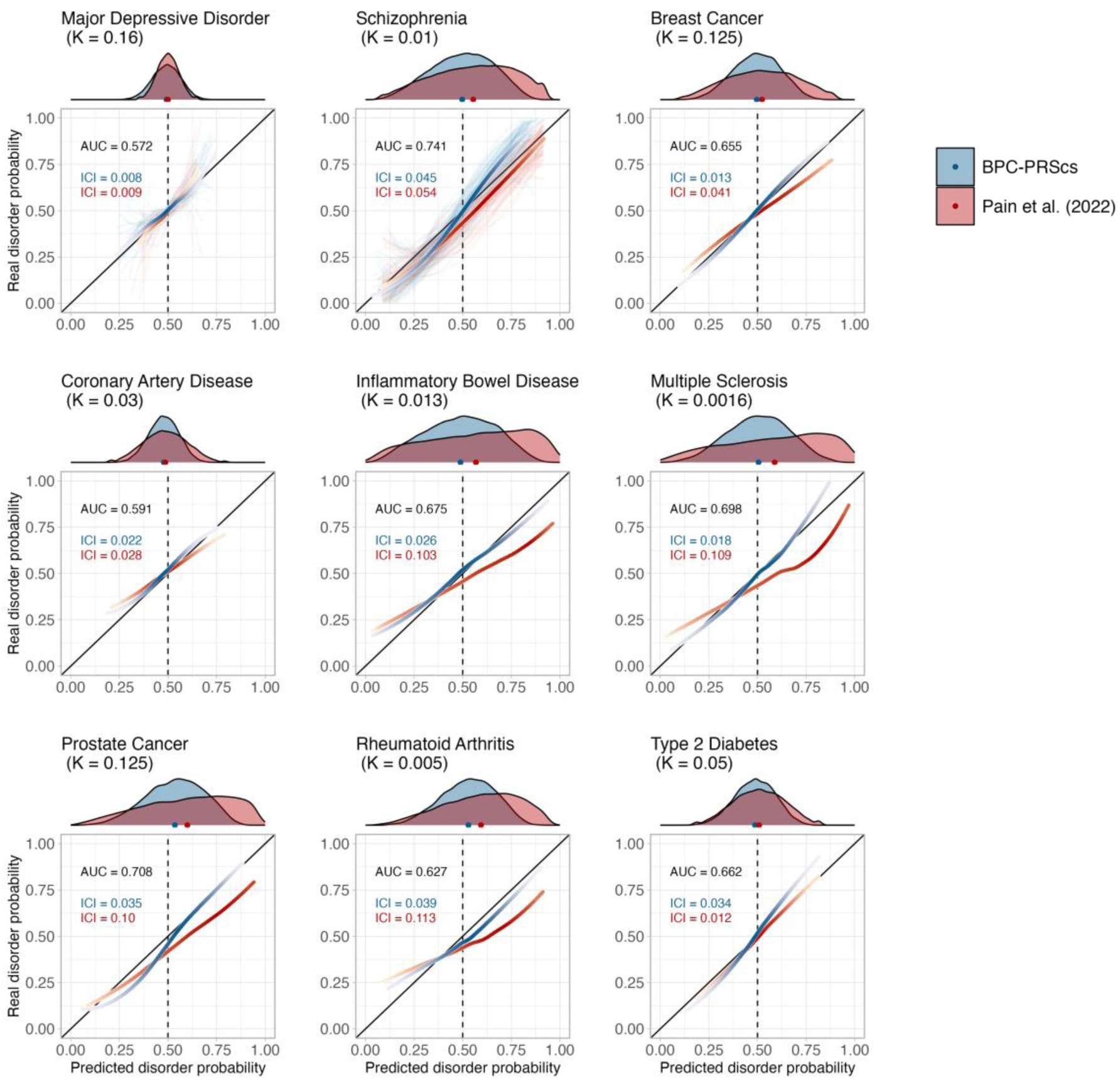
Disorder-specific calibration curves in empirical analyses of nine disorders. Calibration of the BPC and the Pain et al. (2022) approach was evaluated using the Integrated Calibration Index (ICI) for nine disorders, each with a prior disorder probability of 0.5 (see Table 1 for an overview of the case/control testing sample sizes). The prior disorder probability was set to 0.5, as opposed to the lifetime prevalence in the general population (K), to emulate the higher risk of help-seeking individuals in clinical settings. Histograms at the top of the plots depict the distribution of the predicted disorder probabilities, and the dots at the base of the histograms depict the mean predicted probability. The lines were drawn with a loess smoothing function, and their transparency follows the density of the histogram to show which parts of the distribution carry the most weight in the calculation of the ICI. For major depression and schizophrenia, 62 and 22 cohorts, respectively, were available for analysis and therefore depict thin, light-colored, and transparent lines for individual cohorts. In contrast, the thicker and darker lines depict results when data from all cohorts are concatenated. The disorder population lifetime prevalence (K) is reported. The Area Under the receiver operator Curve (AUC) is the same for both approaches because the transformations do not change the ranking of individual PGSs, and both approaches use the same PGS inputs. The BPC-PRScs approach achieves lower ICI values for eight out of nine disorders. The Pain et al. (2022) approach tends to overestimate the predicted disorder probabilities, as seen by the right shift of the histograms and the dots. Numerical values are presented in Supplementary Table 2. Calibration curves for BPC-SBayesR are presented in Supplementary Figure 16.

We performed secondary analyses yielding the following eight conclusions. First, comparing the calibration plots of BPC-PRScs with BPC-SBayesR, the latter makes correct predictions on average but is less well-calibrated for low and high values of the predicted disorder probabilities (see Supplementary Figure 16 and Supplementary Table 2). Second, misspecification of the effective sample size by a factor of 0.5 and 2 negatively impacts calibration for BPC-PRScs, while it does not affect the calibration of the Pain et al. (2022) approach (see Supplementary Figure 17 and Supplementary Table 3) as it involves a scaling step after the posterior mean betas have been computed. We note the BPC approach still has lower median ICI values than the Pain et al. (2022) approach. BPC-SBayesR seems generally more robust to misspecification of the effective sample size, except for Coronary Artery Disease, which suffers extreme miscalibration when N_eff_ is multiplied by 2. Third, misspecification of the prior impacts calibration because it shifts the mean of the predicted disorder probabilities. A mismatch of 0.25 between the true and assumed prior leads to an average increase of 0.21 (s.e.m. 0.02) in the ICI (see Supplementary Figure 18). However, given that the BPC approach is well-calibrated under a range of correctly specified priors, the change of the posterior predicted disorder probability relative to the prior remains informative as it makes the diagnosis less or more likely compared to the prior expectation. In practice, the prior can be estimated from small reference samples, literature, or prior elicitation (see **Discussion** for more information). Fourth, including the MHC region strongly and negatively impacts calibration for the autoimmune disorders Multiple Sclerosis and Rheumatoid Arthritis for BPC-PRScs and Pain et al. (2022) (but not BPC-SBayesR; This is because SBayesR’s reference sample excludes most of the MHC region; see Supplementary Figure 19 and Supplementary Table 4). Fifth, reducing the INFO filter from 0.9 to 0.3 and the minor allele frequency filter from 10% to 1% (as in ^40^) yields comparable average ICI values (except for Coronary Artery Disease and BPC-SBayesR; see Supplementary Figure 20 and Supplementary Table 6). Sixth, evaluating calibration with the slope and intercept from a linear regression of the phenotype on the predicted disorder probabilities also shows that BPC-PRScs is best calibrated overall (see Supplementary Figures 21 and 22, and Supplementary Table S6). Seventh, we tested and confirmed that the BPC’s assumption of normally distributed PGSs in cases and controls holds for all analyzed phenotypes (see Supplementary Figure 23). Eighth, we investigated the distribution of 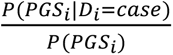 and found it to vary considerably around one (e.g., S.D. = 0.29 for schizophrenia when the prior = 0.50), showing that the predicted disorder probabilities are not solely determined by the prior (see Supplementary Figure 24).

In contrast to simulations (see Supplementary Figures 9 and 10), the untransformed Bayesian PGSs do not show strongly miscalibrated slopes and intercepts (see Supplementary Figures 25 and 26), likely due to the variance of estimates of the calibration slopes in combination with much fewer observations in empirical data (i.e., 9) than in simulations (100 simulation runs for 8 parametrizations). Our findings align with the previous observation that the calibration slope is very sensitive to miscalibration in small parts of the data and that the ICI is more robust and preferred as a metric for calibration^15^. Because untransformed Bayesian PGSs are centered around 0 and do not range from 0 to 1, they cannot be evaluated with the ICI and cannot be interpreted as predicted disorder probabilities.

#### Calibration of tuning approaches

The BPC approach does not require tuning samples to estimate predicted disorder probabilities. However, to benchmark the BPC approach, we compared it to other approaches that utilize such tuning samples that include genotype and phenotype data (see **Methods: Approaches using tuning samples**). The calibration of the BPC approach is similar to the tuning approaches when the tuning samples are smaller than 200 cases and 200 controls (see Figure 5), while the area under the ROC curve (AUC) does not differ between these approaches (see Supplementary Figure 27). For larger tuning sample sizes, the tuning approaches have an ICI that is approximately 0.015 smaller. However, we consider BPC’s calibration (ICI < 0.03) satisfactory, such that the benefit of not requiring a tuning sample outweighs the improved calibration of the tuning approaches.

**Figure 5.**
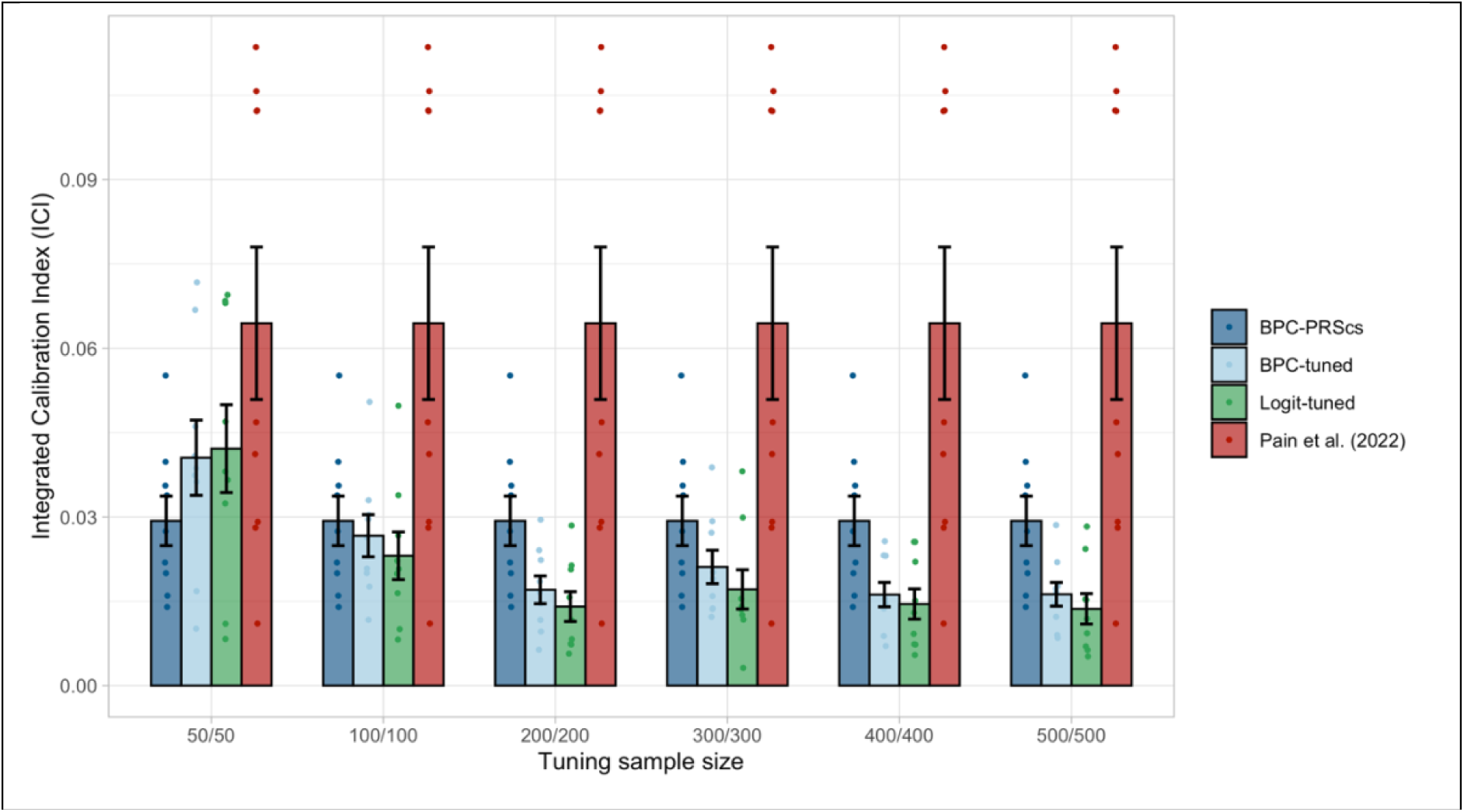
Calibration of tuning approaches in empirical analyses of nine disorders. Calibration of the BPC-PRScs, BPC-tuned, Logit-tuned and the Pain et al. (2022) approach was evaluated using the Integrated Calibration Index (ICI) for nine disorders. BPC-tuned and Logit-tuned use a tuning sample that includes genotype and phenotype data, whereas BPC-PRScs and Pain et al. (2022) do not require an additional independent tuning sample. Tuning sample sizes are presented as (N_case_/N_control_). Error bars represent standard errors.

#### Estimation of variance explained (*R*^2^_*liability*_)

The BPC approach depends on a valid estimate of *R*^2^_*liability*_. Our approach of computing the variance of a well-calibrated PGS in a population reference sample without the need for phenotype data (see **Methods**: *Step 3 Derive R^2^_liability_ and the expected distribution of the PGS in cases and controls*) leads to estimates that are very close to the observed values from linear regression^7^ in a sample with both pheno- and genotype data in simulations (mean absolute difference ranges from 0.009 to 0.011; see Figure 6A) and in empirical data (mean absolute difference = 0.02; see Figure 6B). **R1C4 & R2C4.i** This suggests that the PGSs are well-calibrated on the unobserved liability scale. The Pain et al. (2022) approach uses lassosum^26^, which leads to estimates that are slightly misspecified in simulations (mean absolute difference ranges from 0.058 to 0.088) and in empirical data (mean absolute difference = 0.05).

**Figure 6.**
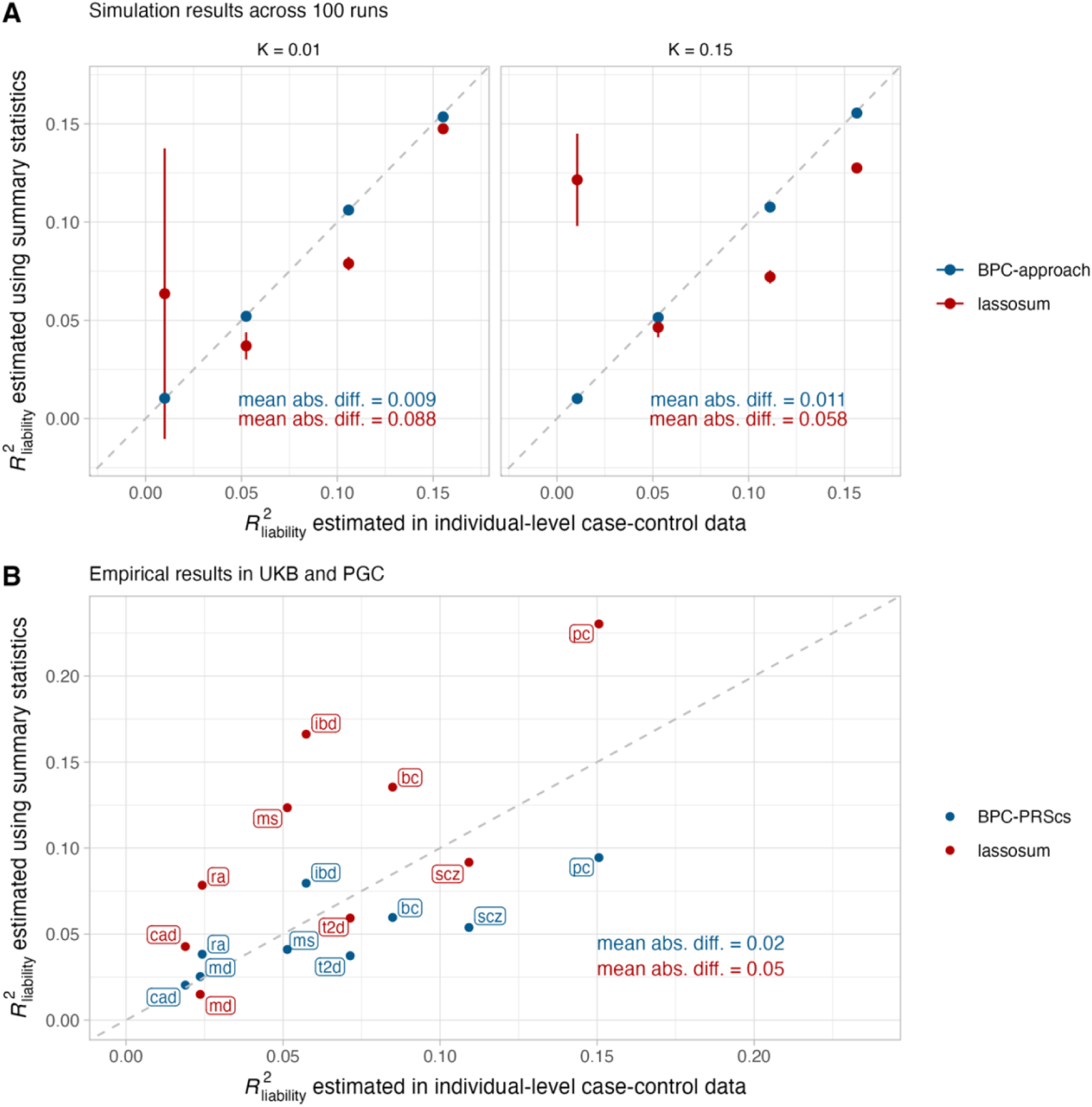
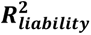 estimates in simulations and empirical analyses of nine disorders. (A) Simulation results of estimating 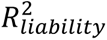 using the BPC approach and lassosum (as used by Pain et al. (2022)), both of which do not require disorder-specific individual-level genotype and phenotype data. The x-axis depicts 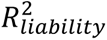 estimated by regressing disorder status on the Bayesian PGS in individual-level data in the testing sample^7^. Error bars depict standard errors for 100 simulation runs. The grey dashed line depicts the identity line when y = x. The BPC approach achieves mean estimates that are closer to the regression results in the testing sample in every simulation condition. mean abs. diff. = mean absolute difference of 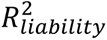 estimates using summary statistics and individual-level case-control data. (B) Empirical results in the UKB and PGC of estimating 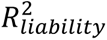 using the BPC-PRScs approach and lassosum. The BPC-PRScs approach achieves estimates that are closer to the regression results in the testing sample on average (mean absolute difference of 0.02 vs. 0.05).

## Discussion

We developed the BPC approach to transform PGSs to absolute risk values, which yields predicted disorder probabilities that may be clinically useful for single individuals. Based on Bayesian PGS methods, it requires only minimal input, namely GWAS summary statistics, a single individual’s genome-wide genotype data and prior disorder probability, and an estimate of the disorder’s population lifetime prevalence. We verified in simulations and empirical analyses of nine disorders that the BPC approach achieves good calibration across a range of prior disorder probabilities, meaning the predicted and real disorder probabilities closely align. The BPC approach depends on a valid estimate of *R*^2^_liability_, which we compute by estimating the variance of a well-calibrated PGS in a population reference sample without the need for phenotype data, and verify that the estimates are close to empirically calculated values in case-control data.

We compared the BPC approach to a recently published approach in Pain et al. (2022)^14^, and showed that it achieves lower ICI values in every simulation condition and for eight out of nine tested disorders in empirical analyses. This is partly because the Pain et al. (2022) approach overestimates the predicted disorder probabilities whenever the prior disorder probability exceeds the population lifetime prevalence. We also compared the BPC approach to methods requiring tuning data^10^. We found that for larger tuning sample sizes of more than 200 cases and controls, the tuning approaches have an ICI that is approximately 0.015 smaller. However, we consider BPC’s calibration (ICI < 0.03) satisfactory, such that the benefit of not requiring a tuning sample outweighs the improved calibration of the tuning approaches.

In clinical settings where a single individual may be considered, the prior disorder probability, which can be interpreted as the case-control ratio in a hypothetical testing sample to which that individual belongs, can be approximated in several ways. It may be estimated using a small external reference sample to obtain a data-informed prior, such as context-specific prevalence estimates of individuals seeking health care for a specific disorder in a given hospital. Such a reference sample does not require genotype data and may be smaller than those required for the tuning approaches. Alternatively, such context-specific prevalence estimates may also be obtained from the literature^19^. The context may refer to any variable that modifies a disorder’s prevalence, such as age or sex ^43^. When no data is available to estimate the prior, *prior elicitation*^44^ may be used, where a clinician (or a panel of clinicians) provides a subjective estimate of the prior. Generally, the lifetime risk for help-seeking individuals is expected to be higher than for individuals from the general population (where lifetime risk = K). As such, the prior will often be higher than K. When considerable uncertainty about the prior exists, a range of priors may be used to obtain a range of posterior disorder probabilities.

There are several limitations to this study. First, because most GWASs are based on individuals from European populations, the calibration of the BPC approach for individuals from non-European populations is unknown but may be negatively affected, as is the accuracy of risk predictions^45,46^. However, as long as the GWAS population matches that of the individual, the BPC approach is expected to be well-calibrated. Future studies are needed to develop methods to obtain well-calibrated predictions for individuals from non-European populations. Second, we performed simulations without LD, which may be perceived as a limitation. However, we note that the results from our simulation and empirical analyses were concordant, suggesting that our simplified simulation setup was appropriate. Third, the potential for clinical utility of polygenic prediction (and thereby the BPC approach) strongly depends on the magnitude of the PGS’s *R*^2^_liability_, which is currently prohibitively small for most traits. However, there are traits, such as coronary artery disease^47–49^, type 2 diabetes^50,51^, breast cancer^4,52,53^, chronic obstructive pulmonary disease^5^, and prostate cancer^6,54^, for which current PGSs may already be sufficiently powered to find clinical application and be economically effective. Moreover, as GWAS sample sizes grow, the PGS’s *R*^2^_liability_ is expected to approach the disorder’s *h*^2^_SNP_, and therefore, their clinical applicability will become more likely. Fourth, the calibration of the predicted disorder probabilities depends on a correct estimate of the prior. While we showed that misspecification of the prior negatively impacts calibration, we also showed that the BPC approach is well-calibrated across a range of correctly specified priors and that, therefore, the change of the posterior predicted disorder probability relative to the prior remains informative. Irrespectively, striving for the best possible prior disorder probabilities in practice is important and provides an important direction for future research. Fifth, the BPC approach can only be applied to polygenic traits with normally distributed PGSs in cases and controls. While we show that this assumption holds in our simulation and empirical analyses (Supplementary Figure 12), its violation due to outlying common, very large-effect variants can negatively impact calibration, such as APOE for Alzheimer’s Disease^55^, and should be removed prior to the application of the BPC approach. Integrating prediction based on rare variants with large effects with polygenic prediction is an important direction for future research. Sixth, while variables that are not correlated to the PGS (e.g., sex, age) can easily be used to adjust the prior, variables that are correlated to the PGS (e.g., family history^56–59)^ cannot currently be incorporated into the BPC approach because, in this case, the prior cannot be adjusted independently without modifying the 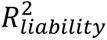. Extending the BPC approach to include variables correlated to the PGS is an important direction for future research. Seventh, the BPC outcome is presented as a fixed lifetime probability. Extending the BPC approach to model the decline in risk in the years following the assessment in which the disorder has not manifested is an important direction for future research.

In conclusion, the BPC approach provides an effective tool to compute well-calibrated predicted disorder probabilities based on polygenic scores.

## Data Availability

Individual-level data from the Psychiatric Genomics Consortium (https://pgc.unc.edu/) and the UK Biobank (https://www.ukbiobank.ac.uk/enable-your-research/apply-for-access) cannot be shared freely, but an access application is required first. The GWAS summary statistics used in the UKB analyses can be requested or downloaded from the following web pages: Breast Cancer (https://bcac.ccge.medschl.cam.ac.uk/bcacdata/oncoarray/oncoarray-and-combined-summary-result/gwas-summary-associations-breast-cancer-risk-2020/); BMI (https://portals.broadinstitute.org/collaboration/giant/index.php/GIANT_consortium_data_files); Coronary Artery Disease (http://www.cardiogramplusc4d.org/data-downloads/#); Inflammatory Bowel Disease (https://www.ibdgenetics.org/); Multiple Sclerosis (https://imsgc.net/?page_id=31); Prostate Cancer (http://practical.icr.ac.uk/blog/?page_id=8164); Rheumatoid Arthritis (https://data.cyverse.org/dav-anon/iplant/home/kazuyoshiishigaki/ra_gwas/ra_gwas-10-28-2021.tar); Type 2 Diabetes (https://diagram-consortium.org/downloads.html). GWAS summary statistics for Major Depression and Schizophrenia can be downloaded from the PGC website (https://pgc.unc.edu/for-researchers/download-results/).
1000 Genomes reference files can be downloaded from https://ctg.cncr.nl/software/magma.

## Acknowledgments

We thank Naomi Wray, Peter Visscher, and Oliver Pain for their helpful discussions. D.P. is supported by the Netherlands Organization for Scientific Research—Gravitation project ‘BRAINSCAPES: A Roadmap from Neurogenetics to Neurobiology’ (024.004.012) and the European Research Council advanced grant ‘From GWAS to Function’ (ERC-2018-ADG 834057). The PGC has received major funding from the US National Institute of Mental Health (PGC4: R01MH124839, PGC3: U01 MH109528; PGC2: U01 MH094421; PGC1: U01 MH085520). A.L.P has received an R01 grant from the US National Institutes of Health (HG006399). We thank the participants who donated their time, life experiences, and DNA to this research and the clinical and scientific teams that worked with them. We are deeply indebted to the investigators who comprise the PGC. Statistical analyses were carried out on the NL Genetic Cluster Computer (http://www.geneticcluster.org) hosted by SURFsara. The content is solely the responsibility of the authors and does not necessarily represent the official views of the National Institutes of Health.

## Author contributions

**Emil Uffelmann**: Methodology, Software, Formal analysis, Investigation, Data Curation, Writing - Original Draft, Visualization

**Alkes Price:** Writing - Review & Editing

**Danielle Posthuma**: Writing - Review & Editing, Funding acquisition, Supervision

**Wouter J. Peyrot**: Conceptualization, Methodology, Software, Resources, Writing – Original Draft and Review & Editing, Supervision

## Web resources

PRScs https://github.com/getian107/PRScs

SBayesR https://cnsgenomics.com/software/gctb/#Overview

1000 Genomes files https://ctg.cncr.nl/software/magma

## Data and code availability

Scripts to apply the BPC approach can be downloaded from https://github.com/euffelmann/bpc. Individual-level data from the Psychiatric Genomics Consortium (https://pgc.unc.edu/) and the UK Biobank (https://www.ukbiobank.ac.uk/enable-your-research/apply-for-access) cannot be shared freely, but an access application is required first. The GWAS summary statistics used in the UKB analyses can be requested or downloaded from the following web pages: Breast Cancer (https://bcac.ccge.medschl.cam.ac.uk/bcacdata/oncoarray/oncoarray-and-combined-summary-result/gwas-summary-associations-breast-cancer-risk-2020/); BMI (https://portals.broadinstitute.org/collaboration/giant/index.php/GIANT_consortium_data_files); Coronary Artery Disease (http://www.cardiogramplusc4d.org/data-downloads/#); Inflammatory Bowel Disease (https://www.ibdgenetics.org/); Multiple Sclerosis (https://imsgc.net/?page_id=31); Prostate Cancer (http://practical.icr.ac.uk/blog/?page_id=8164); Rheumatoid Arthritis (https://data.cyverse.org/dav-anon/iplant/home/kazuyoshiishigaki/ra_gwas/ra_gwas-10-28-2021.tar); Type 2 Diabetes (https://diagram-consortium.org/downloads.html). GWAS summary statistics for Major Depression and Schizophrenia can be downloaded from the PGC website (https://pgc.unc.edu/for-researchers/download-results/). 1000 Genomes reference files can be downloaded from https://ctg.cncr.nl/software/magma.

