## Supplementary Note for "Estimating Disorder Probability Based on Polygenic Prediction Using the BPC Approach"

Supplementary Note 1. Effective sample size

The effective sample size, N_eff_, of the training GWAS results is an important input value to attain well-calibrated results with the BPC approach. The effective sample size of a study is the sample size with a case-control ratio of 0.5, which yields the same power as the study of interest^1^. For example, a study with total N of 10,000 with 1,000 cases and 9,000 controls (case-control ratio = 0.1) yields the same power as a sample with 1,800 cases and 1,800 controls (case-control ratio = 0.5), and this has a N_eff_ of 3,600 (i.e., much smaller than the total N of 10,000).

We describe three ways of obtaining N_eff_. First, N_eff_ is sometimes provided by the meta-analysis software in the GWAS results, in which case we advise using this value of N_eff_.

Second, N_eff_ can be computed using the number of cases and controls. In this case, it is important to compute N_eff_ per cohort contributing to the meta-analyses and use the sum of the N_eff_’s of the contributing cohorts as overall N_eff_ ^1^. For example, a meta-analysis of a cohort with 1,000 cases and 9,000 controls and a cohort of 9,000 cases and 1,000 controls yields a N_eff_ of 3,600+3,600=7,200 (which is much less than 20,000, which would have been obtained by summing the cases, summing the controls and then computing N_eff_). Given a sample with $N_{case}$ cases and $N_{control}$ controls, N_eff_ can be computed with the formula

$$N_{eff}=\frac{4}{(\frac{1}{N_{case}}+ \frac{1}{N_{control}})}$$

This formular follows from the noncentrality parameter (NCP). The NCP is computed as

$NCP= \frac{{(p_{case}- p_{control})}^{2}}{\bar{p}(1-\bar{p})(\frac{1}{2N_{case}}+ \frac{1}{2N_{control}})}$ , where $p_{case}$ and $p_{control}$ are the allele frequencies in cases and controls, respectively, and $\bar{p}$ is the average allele frequency across both cases and controls.

Now, the formula for $N_{eff}$follows as:

$$NCP_{case-control ratio=0.5}=NCP_{case-control ratio=N_{case}/(N_{case}+N_{control})}$$

$$\Longrightarrow$$

$$\frac{{(p_{case}- p_{control})}^{2}}{\bar{p}(1-\bar{p})(\frac{1}{N_{eff}}+ \frac{1}{N_{eff}})}= \frac{{(p_{case}- p_{control})}^{2}}{\bar{p}(1-\bar{p})(\frac{1}{2N_{case}}+ \frac{1}{2N_{control}})}$$

$$\Longrightarrow$$

$$N_{eff}=\frac{4}{(\frac{1}{N_{case}}+ \frac{1}{N_{control}})}$$

Third, when N_eff_ is not provided in the GWAS results and when the number of cases and controls of the cohorts contributing to the meta-analysis are not known, $N_{eff}$ can also be derived analytically (outlined in detail by Grotzinger et al.^1^), based on the genotype variance, $2 \times AF \times\left( 1-AF \right)$, and the sampling variance, ${SE}^{2}$:

$N_{eff}=\frac{4}{2 \times AF \times\left( 1-AF \right) \times{SE}^{2}}$ , where AF is the allele frequency of the SNP of interest, and SE is the standard error of the SNP’s beta estimate on the log-odds scale.

For continuous phenotypes, N_eff_ as defined here has no natural counterpart. That is, the effective sample size for continuous phenotypes is simply the total sample size (as long as the study sample is representative of the population; the effective sample-size of non-random samples of a continuous phenotype falls out of the scope of this paper).

Supplementary Note 2. Standardized observed scale with 50% case ascertainment

As described in Peyrot et al.^2^, converting GWAS results to the standardized observed scale (both genotype and phenotype standardized to a mean of 0 and variance of 1) with 50% case ascertainment is a convenient scale to use, because of two main properties:

1. $SE= \frac{1}{\sqrt{N_{eff}}}$ , where SE is the standard error^2^.
2. $\beta^{2}=R^{2}$ , where $\beta$ is the beta for a SNP from a GWAS, and $R^{2}$ is the explained variance in the phenotype of interest by the genotype.

‘Standardized’ means that the phenotype and genotype are scaled to mean=0 and variance=1. ‘Observed’ means the betas are based on a linear regression of standardized case-control status coded as 1-0 (i.e., lm(scale(0-1) ~ SNP)). Because z-values are scale independent (i.e., $z= \frac{\beta}{SE}$), filling in the above formula of SE (1) in $\beta= z \times SE$ gives:

$$\beta_{50/50}= \frac{z}{\sqrt{N_{eff}}}$$

As such, we can easily change the scale of a set of SNP betas to the standardized observed scale with 50% case ascertainment.

Supplementary Note 3. Liability scale conversion

We transform the beta’s from the observed scale ($\beta_{50/50}$) to the betas on the liability scale ($\beta_{l}$) based on the equation from Lee et al.^3^ to transform observed scale heritability estimates ($h_{o}^{2}$) to the liability scale ($h_{l}^{2}$):

$h_{l}^{2}= h_{o}^{2}\times\frac{K(1-K)}{z^{2}}\times\frac{K(1-K)}{P(1-P)}$,

where *K* is the population prevalence, *P* is the case proportion, and $z$ is the height of the standard normal probability density function at a threshold corresponding to $K$. This same formula can be used to transform the scale of individual GWAS betas because $\beta^{2}=R^{2}$ (see Supplementary Note 2). This is achieved as follows:

$\beta_{l}=\beta_{o}\times\sqrt{\frac{K(1-K)}{z^{2}}\times\frac{K(1-K)}{P(1-P)}}$

Substituting *P* = 0.5 when the betas are on the standardized observed scale with 50% case ascertainment:

$\beta_{l}=\beta_{50/50}\times\sqrt{\frac{K(1-K)}{z^{2}}\times\frac{K(1-K)}{{0.5}^{2}}}$

$\beta_{l}=\beta_{50/50}\times\frac{K(1-K)}{z^{2} \times0.5}$

Supplementary Note 4. Mean and variance of PGS in cases and controls

Based on the population prevalence (K) of a disorder of interest and the explained variance of the PGS on the liability scale ($R_{l}^{2}$), we can estimate the means and variances of the PGS in cases and controls using normal theory.

In short, as described previously^4–6^, for any *K*, $T$ and $z$ can be defined such that $K$ equals $P\left( l>T|l\sim N\left( 0,1 \right) \right)$, $z$ is the height of the standard normal distribution at threshold ($T$). The mean liability ($l$) in cases follows from normal theory as $\mu_{l_{cases}}= \frac{z}{K}$ ^4^. Given that the PGS describes $R_{l}^{2}$ variance in the liability, the mean PGS in cases follows as $\mu_{PGS_{cases}}=\mu_{l_{cases}}\times R_{l}^{2}$. Again, following normal theory, the variance of the PGS in cases can be estimated using Tallis’ rule^5^ as $\sigma_{PGS_{cases}}^{2}=R_{l}^{2}-i_{cases}\times\left( i_{cases}-T \right)\times R_{l}^{2}\times R_{l}^{2}$. Means and variances for controls can be derived by substituting $1-K$ for $K$, and multiplying z with -1.

Supplementary Note 5. Simulation Set-up.

We simulated individual liabilities and 1,000 genotypes (500 causal and 500 non-causal) in linkage equilibrium based on the liability threshold model^7^, such that an individual was designated a case if the liability exceeded the disorder lifetime prevalence (K)-dependent threshold. The liability is defined as $Y \sim X\beta+ e$, where *Y* is the vector of liability values and distributed as $Y \sim N(0,1)$, *X* is a vector of genotype values, *β* is the vector of corresponding fixed effect sizes, and *e* is a vector of residual effects. The disorder lifetime prevalence (K)-dependent threshold is computed in R as -qnorm(K,0,1). If *Y >* -qnorm(K,0,1), an individual is designated a case and a control otherwise. The disorder’s SNP-based heritability was set to 0.2. We simulated three independent samples: A training sample used to estimate SNP effects, individuals in a testing sample for which we want to obtain predicted disorder probabilities, and a population reference sample emulating 1000 Genomes^8^ in our empirical analyses to estimate ${R^{2}}_{liability}$ without using phenotype data (see Methods). We repeated the simulations 100 times for eight different parameter settings where we varied the power of the training sample and thereby the coefficient of determination (*R*^2^) of the PGS (${R^{2}}_{liability}$ = {0.01, 0.05, 0.10, 0.15}), as well as the disorder population lifetime prevalence (K = {0.01, 0.15}). The training and testing case-control ratio was set to P = 0.5, while the population reference sample case-control ratio was set to K. To achieve the desired ${R^{2}}_{liability}$ in the testing sample, we estimated the required sample size of the training sample using the avengeme package in R^9^. As such the training sample size differed for every parameter combination and is reported in the table below. The testing sample size was set to 2,000, and the population reference sample to 500 for every simulation run. We computed posterior mean betas using Bpred, a version of LDPred that assumes linkage equilibrium^10^, with GWAS betas on the standardized observed scale with P = 0.5 and therefore used N_eff_ as input. The posterior mean betas were subsequently used to calculate PGSs for every individual in the testing and population reference sample. We applied the BPC approach to estimate predicted disorder probabilities and compared it to the existing approach introduced in Pain et al. (2022)^11^, as well as an alternative approach we developed, the linear rescaling approach (see Methods). We assessed the calibration of the predicted disorder probabilities using the ICI^12^ (see Methods).

**Training (case/control) sample sizes across all parameter combinations**

|  | | K | |
| --- | --- | --- | --- |
|  |  | **0.01** | **0.15** |
| ${\boldsymbol{R}^{\boldsymbol{2}}}_{\boldsymbol{liability}}$ | **0.01** | 145/145 | 315/315 |
|  | **0.05** | 920/920 | 1994/1994 |
|  | **0.10** | 2759/2759 | 5980/5980 |
|  | **0.15** | 7851/7851 | 17015/17015 |

### Supplementary Figures

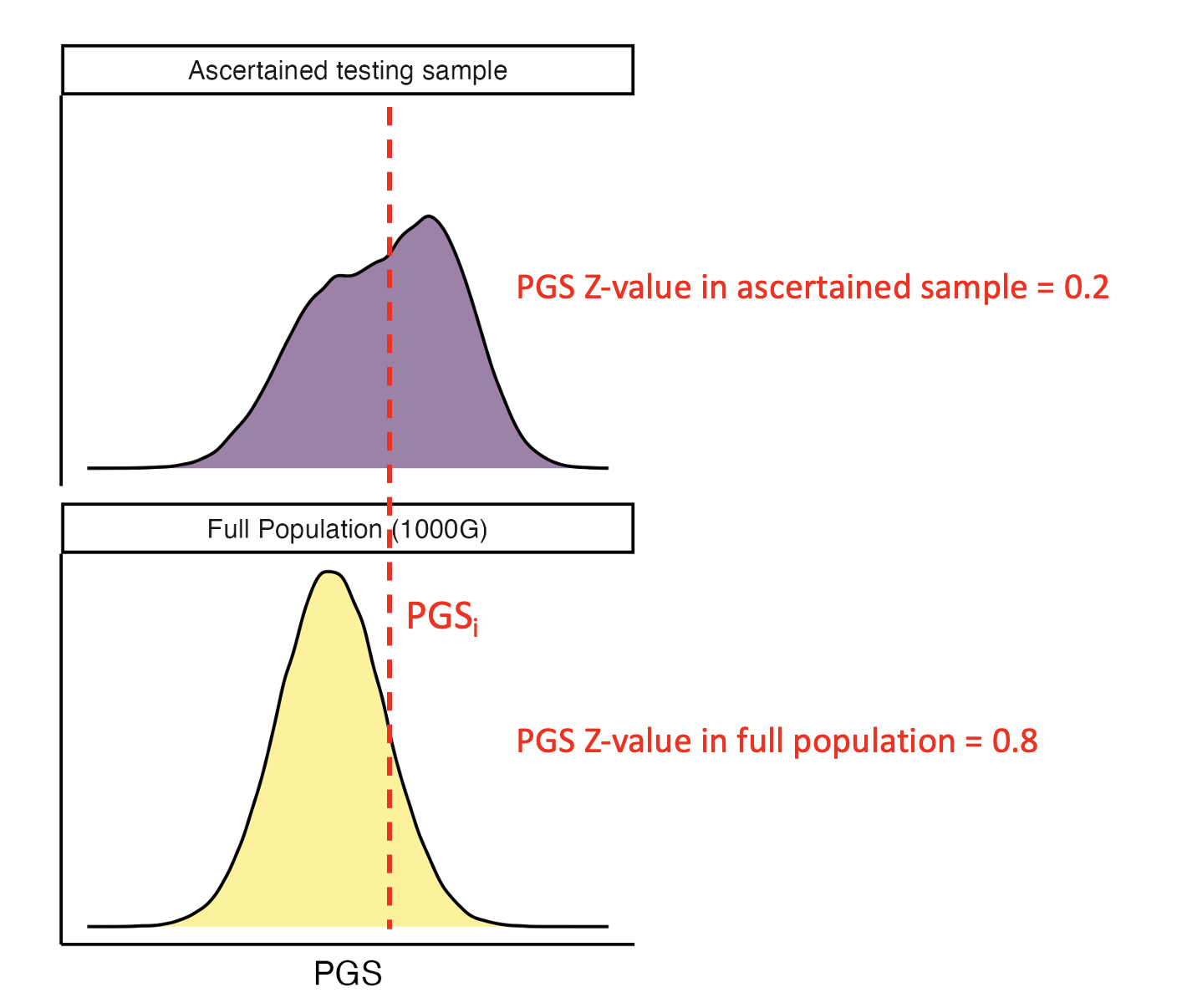

**Supplementary Figure 1** **Overestimation of PGS Z-values in Pain et al. (2022).**

Hypothetical density plots of a PGS distribution in the full population and a testing sample ascertained for the disorder. The variance and the mean of the PGS are larger in the ascertained sample, while the Pain et al. (2022) approach implicitly assumes that the variance and the mean of the PGS in the full population are the same as in the ascertained sample. As such, PGS Z-values based on the full population (e.g. 1000 Genomes) will overestimate the PGS Z-values in the ascertained sample, and consequently also the predicted disorder probabilities.

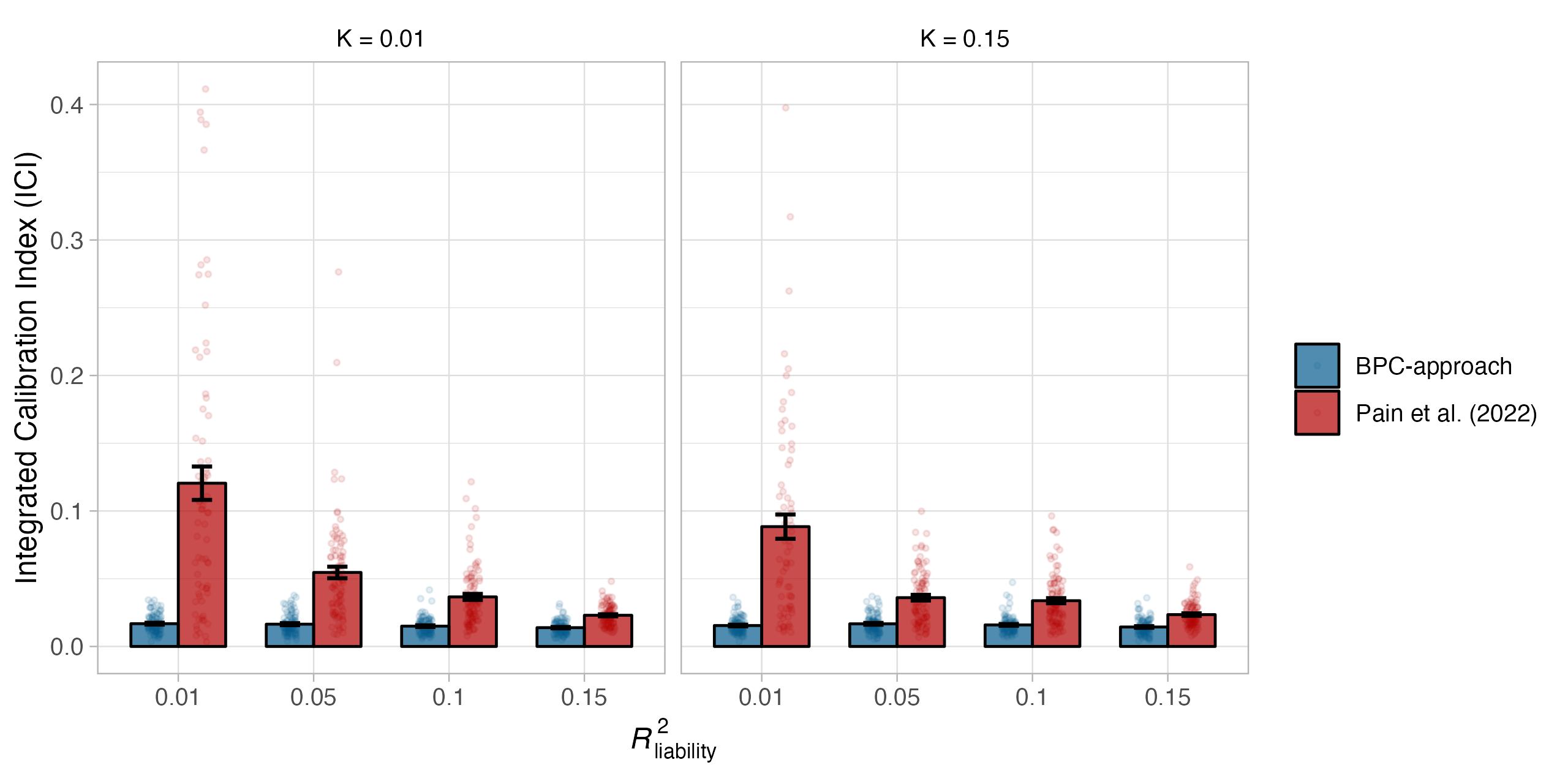

**Supplementary Figure 2 Calibration of adjusted Pain et al. (2022) approach in simulations.**

Calibration of the BPC and the Pain et al. (2022) approach was evaluated using the Integrated Calibration Index (ICI) in 100 simulation runs and for combinations of two parameters, the population prevalence (K), and the explained variance of the PGS on the liability scale (*R*^2^_liability_). The Pain et al. (2022) approach was adjusted to take both the population and testing sample prevalence (i.e. the prior) into account. The BPC approach achieves low mean ICI values in every condition, while the mean ICI values of the Pain et al. (2022) approach are consistently larger. Error bars denote the standard error across 100 simulation runs.

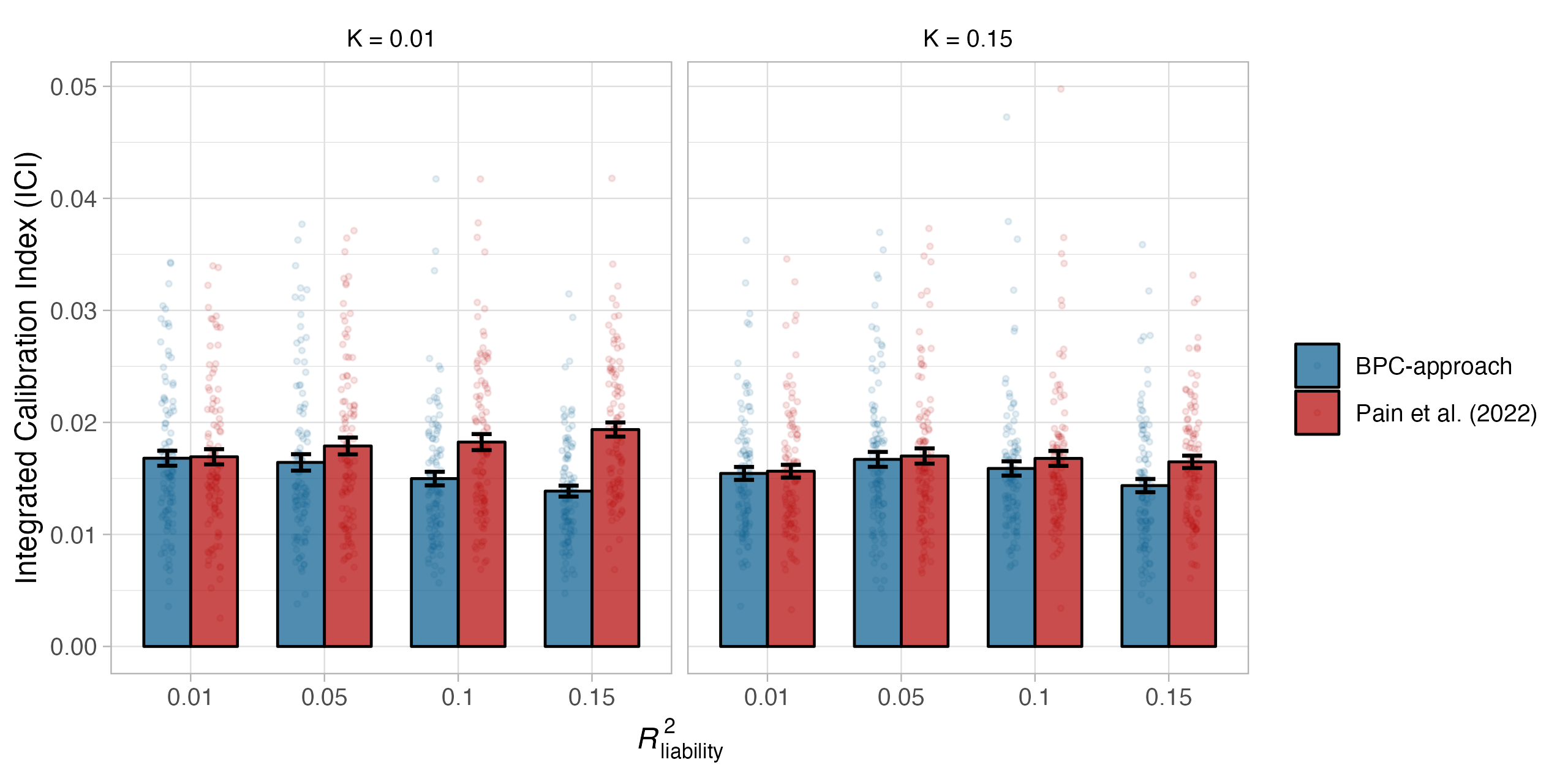

**Supplementary Figure 3 Calibration of adjusted Pain et al. (2022) approach in simulations.**

Calibration of the BPC and the Pain et al. (2022) approach was evaluated using the Integrated Calibration Index (ICI) in 100 simulation runs and for combinations of two parameters, the population prevalence (K), and the explained variance of the PGS on the liability scale (*R*^2^_liability_). The Pain et al. (2022) approach was adjusted to take both the population and testing sample prevalence (i.e. the prior) into account, and we computed the variance of a well-calibrated PGS in a population reference sample to estimate ${R^{2}}_{liability}$ instead of using lassosum. While the difference between both approaches becomes very small, the BPC approach achieves slightly better calibration in nearly every condition. The difference between both methods is largest for conditions with low population prevalence and large ${R^{2}}_{liability}$values. Error bars denote the standard error across 100 simulation runs.

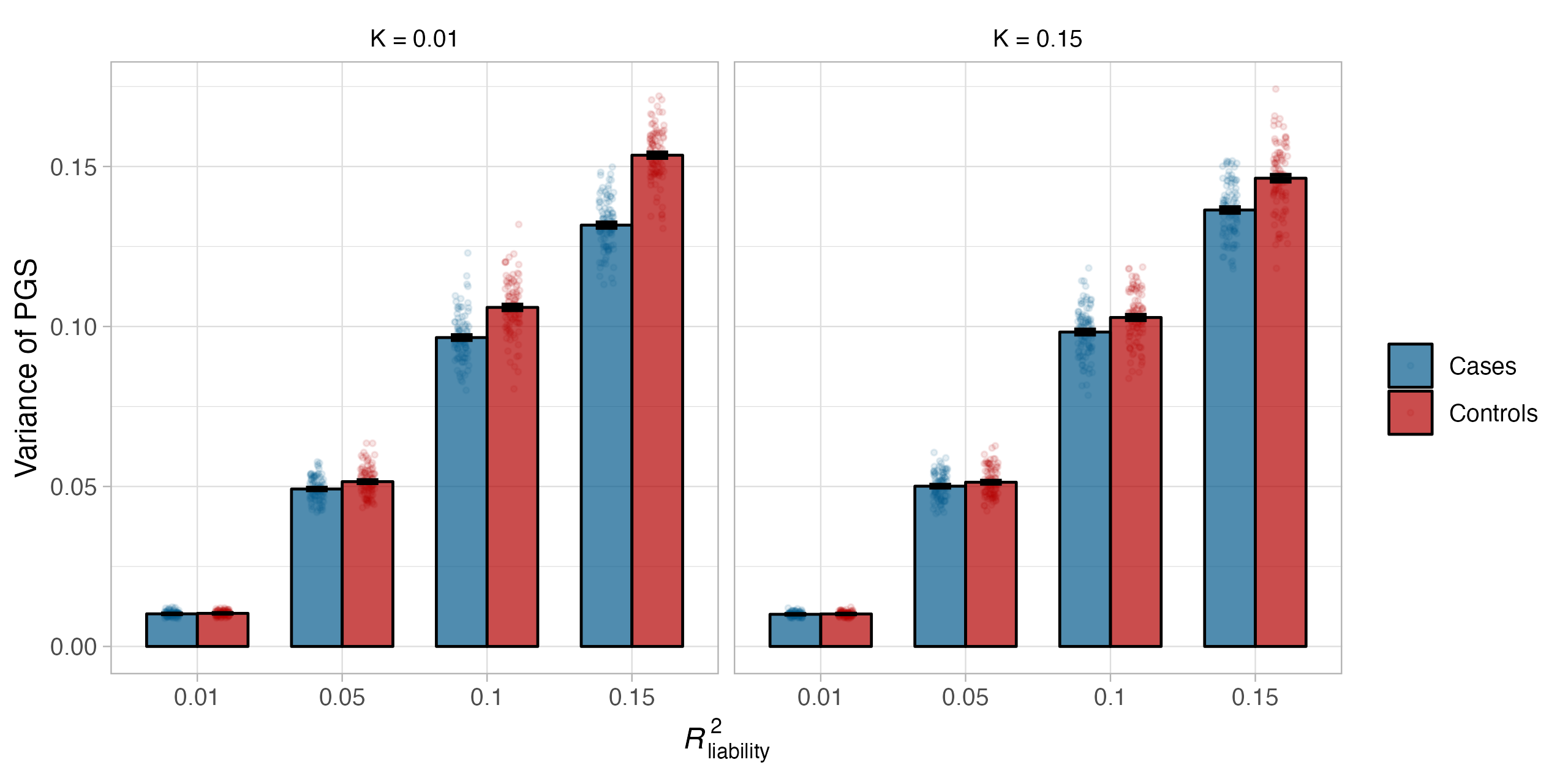

**Supplementary Figure 4 Variance of PGSs in cases and controls.**

Variances of PGSs in cases and controls. The variances in controls are larger than in cases, with the difference between cases and controls becoming larger for low population prevalence and large ${R^{2}}_{liability}$values. Note that the PGSs are well-calibrated on the liability scale and that the variance of the PGS has a clear relationship with the variance explained (not only in the full population, see equation 1 in the manuscript, but also in cases and controls). Error bars denote the standard error across 100 simulation runs.

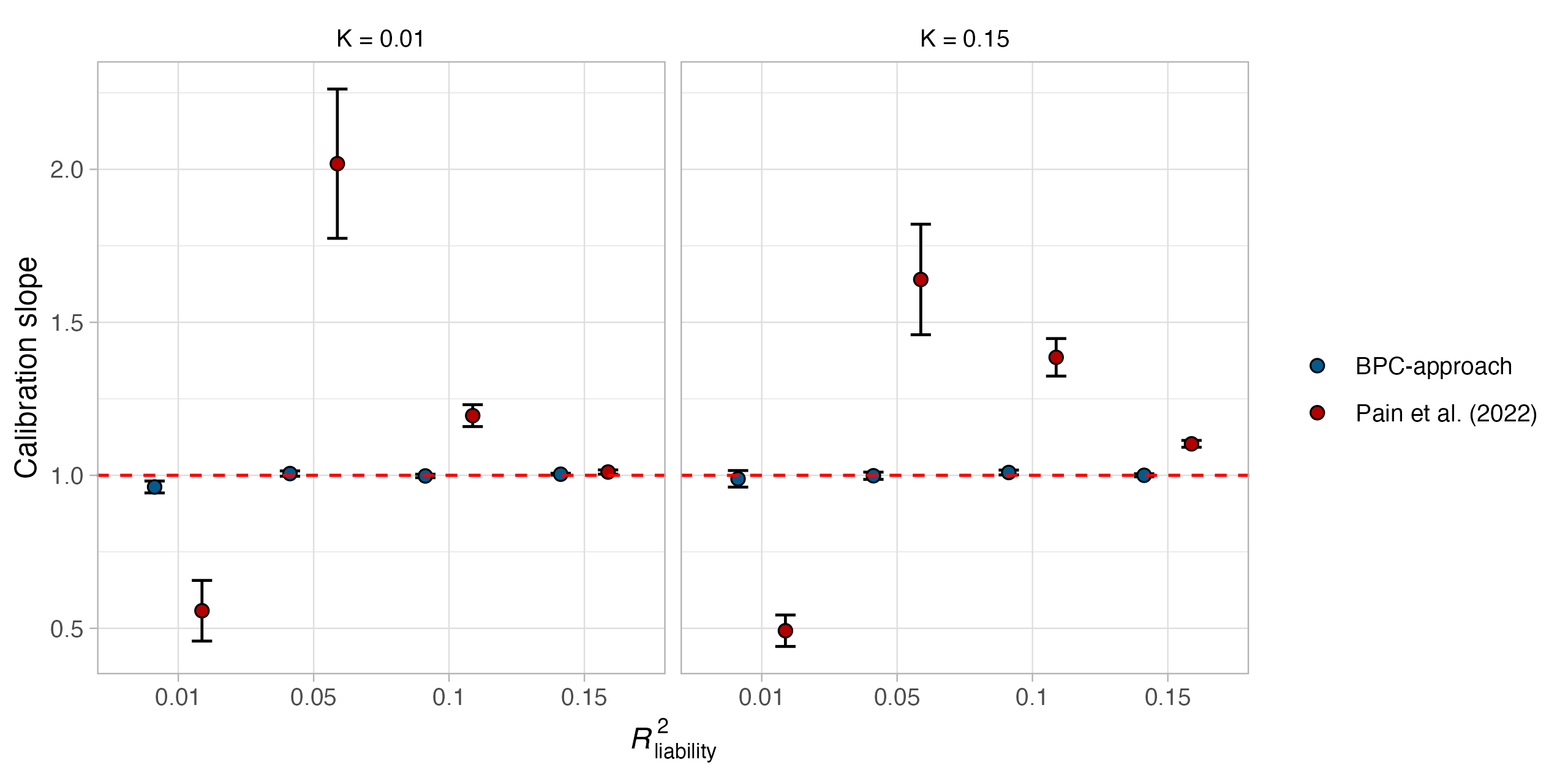

**Supplementary Figure 5 Calibration in simulations (regression slope).**

Evaluating the calibration of the BPC and Pain et al. (2022) approach using the slope from a regression of the standardized disorder status on the PGS. Deviation from 1 indicates miscalibration. The BPC approach consistently achieves good calibration and performs better than the Pain et al. (2022) approach. Error bars denote the standard error across 100 simulation runs.

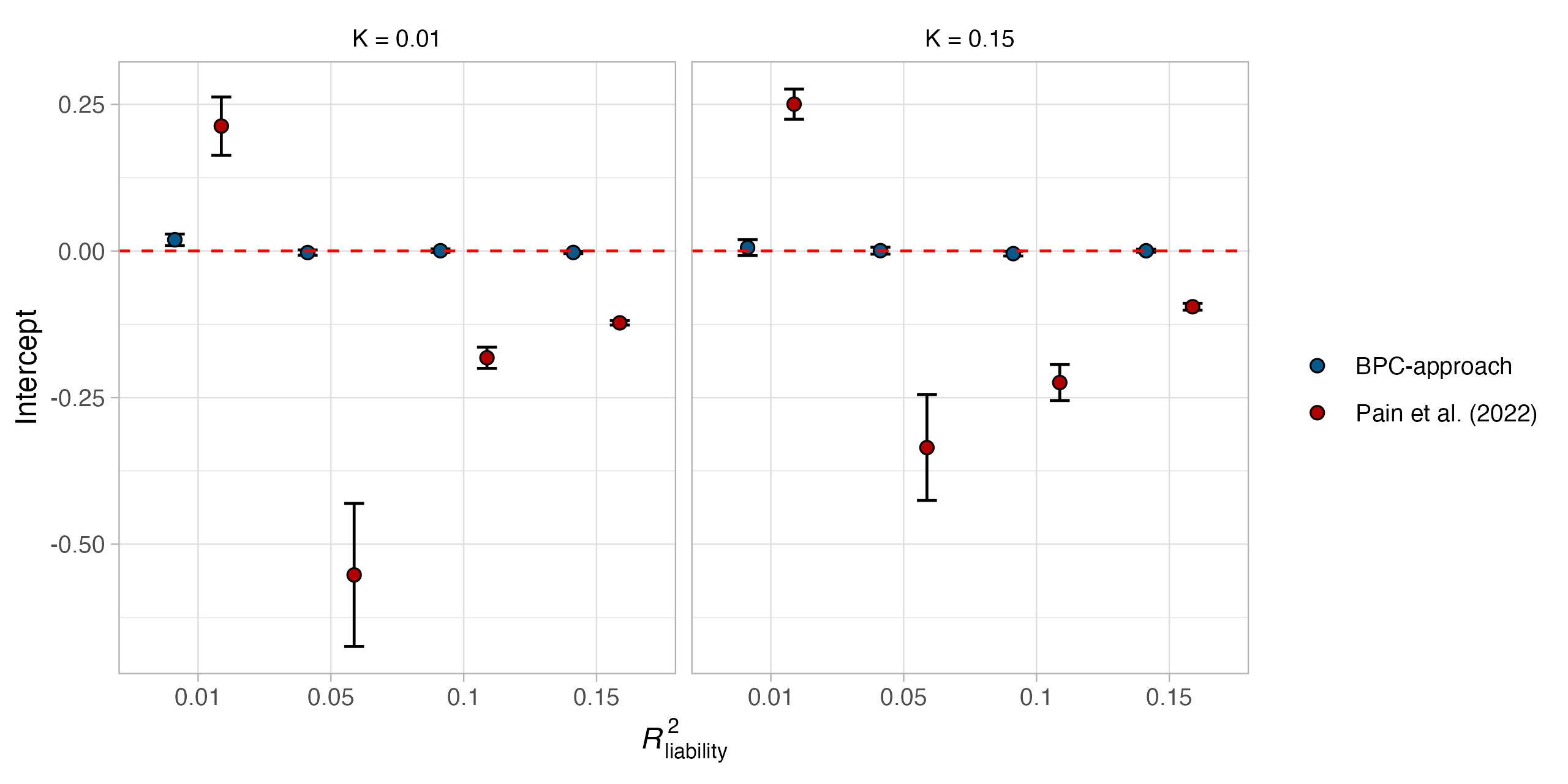

**Supplementary Figure 6 Calibration in simulations (intercept).**

Evaluating the calibration of the BPC and Pain et al. (2022) approach using the intercept from a regression of the standardized disorder status on the PGS. Deviation from 0 indicates miscalibration. The BPC approach consistently achieves good calibration and performs better than the Pain et al. (2022) approach. Error bars denote the standard error across 100 simulation runs.

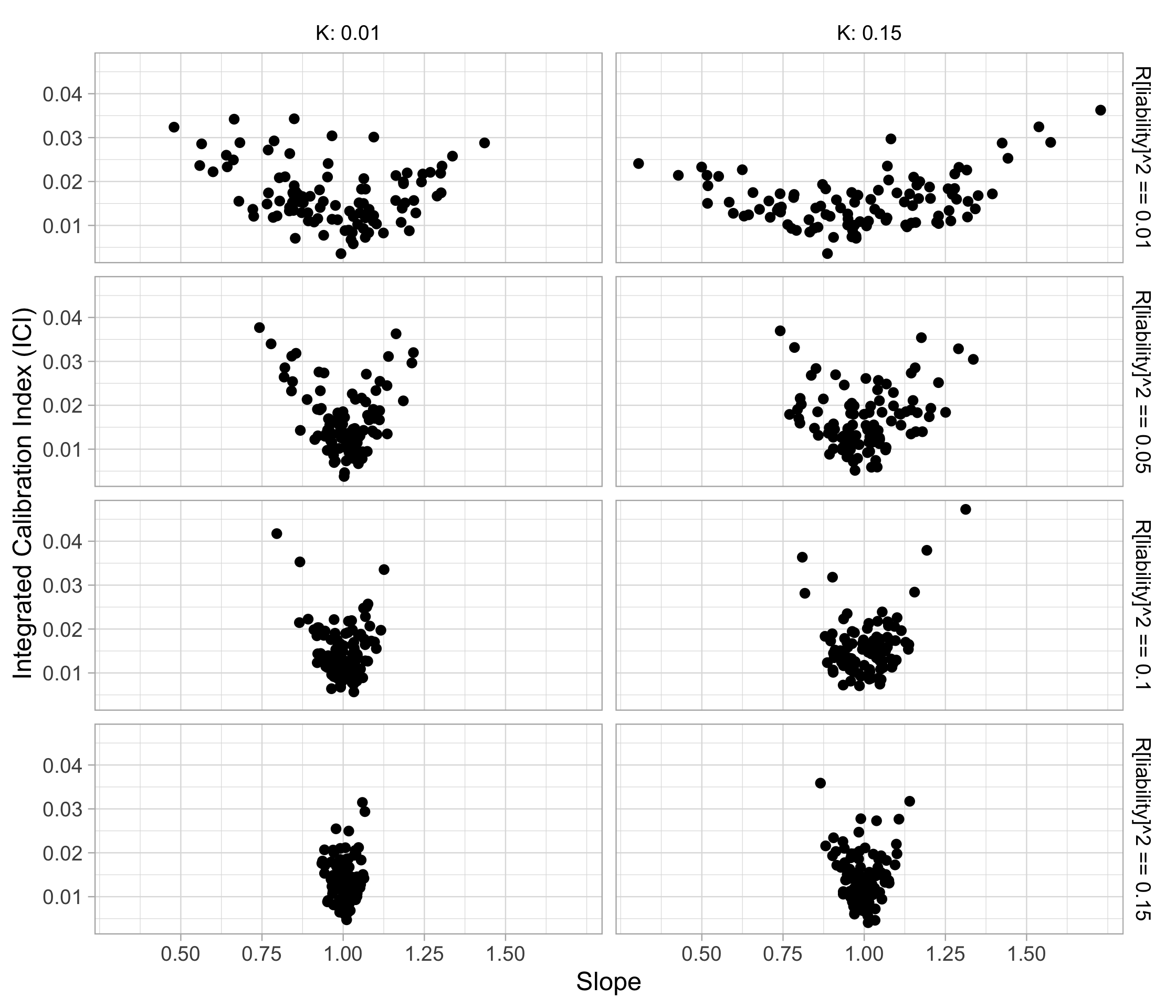

**Supplementary Figure 7 ICI vs. slope in simulations**

Evaluating the stability of the Integrated Calibration Index (ICI) and the Slope. Across different values of the disorder prevalence (K) and $R_{liability}^{2}$, the ICI appears to be very stable with values ranging between 0.04 and 0. The slope is strongly affected by both K and $R_{liability}^{2}$, with increasing variance for larger values of both parameters. Values are based on simulation results for the BPC approach presented in Main Figure 2.

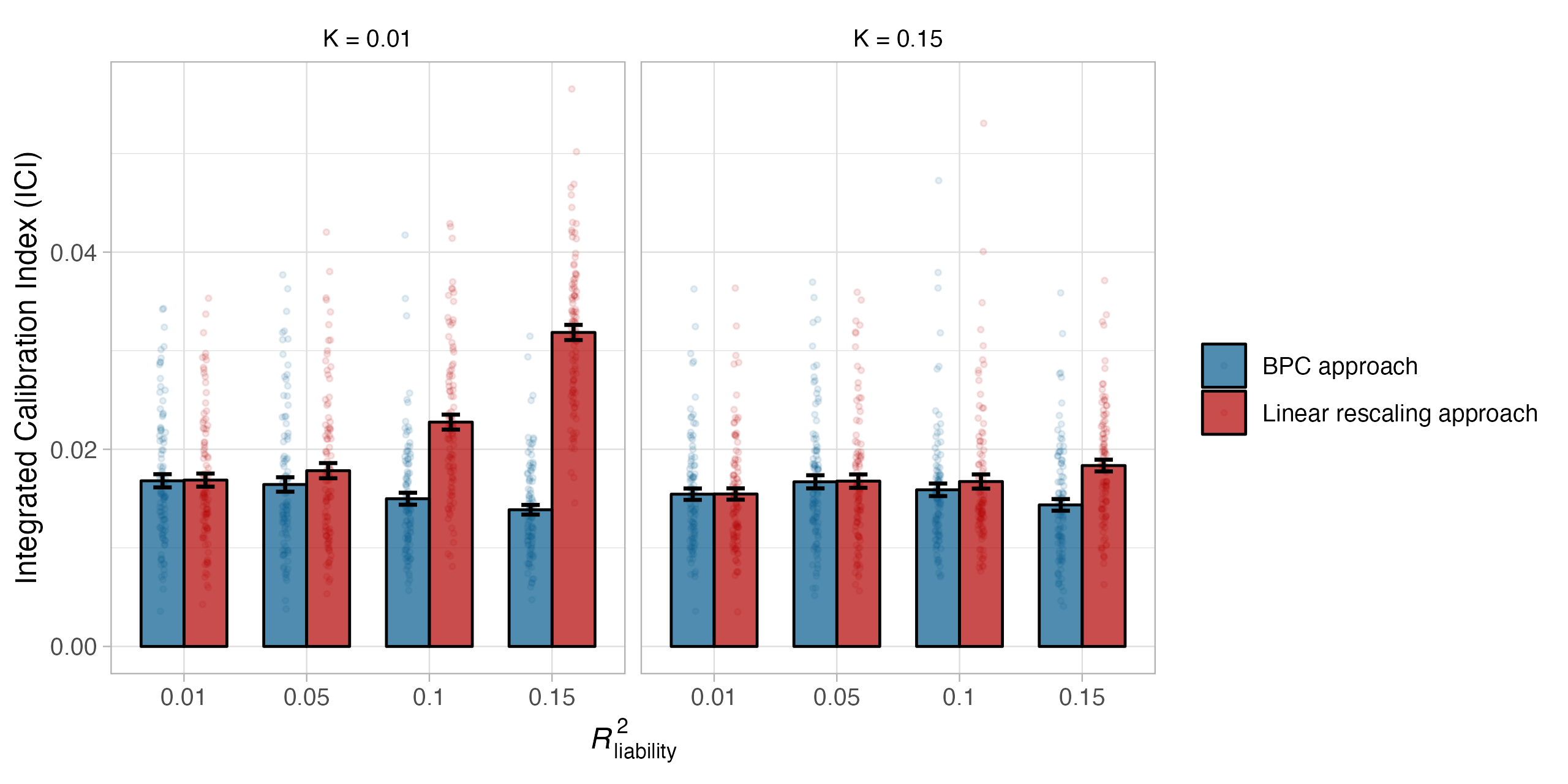

**Supplementary Figure 8 Calibration of the linear rescaling approach in simulations.**

Calibration of our linear rescaling approach was evaluated using the Integrated Calibration Index (ICI) in 100 simulation runs and for combinations of two parameters, the population prevalence (K), and the explained variance of the PGS on the liability scale (*R*^2^_liability_). The linear rescaling approach performs worse for conditions with low population prevalences and large $R_{liability}^{2}$ values. Error bars denote the standard error across 100 simulation runs.

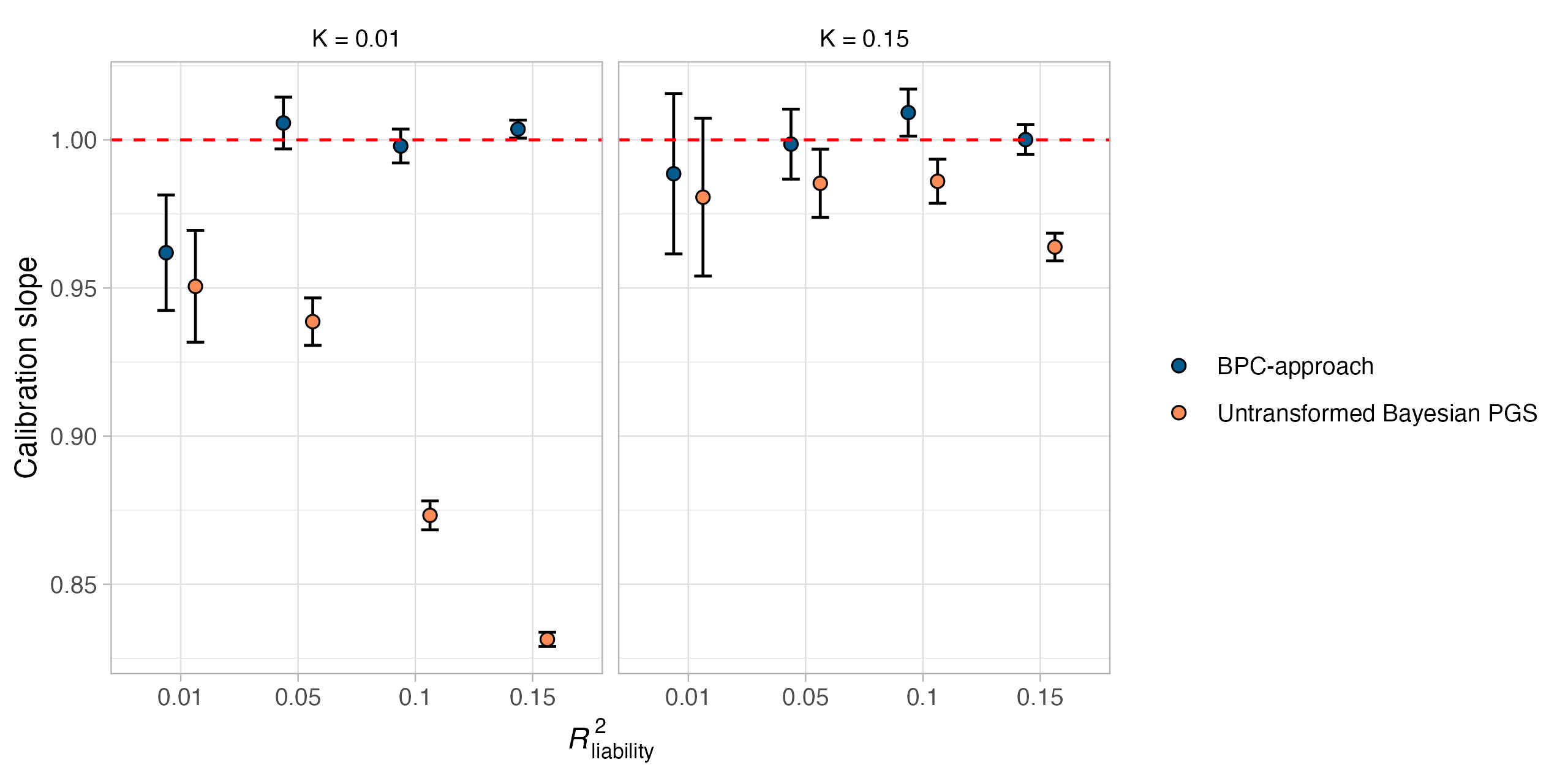

**Supplementary Figure 9 Miscalibration of untransformed Bayesian PGSs (slope).**

Evaluating the calibration of untransformed Bayesian PGSs for binary traits using the slope from a regression of the standardized disorder status on the PGS in 100 simulation runs and for combinations of two parameters, the population prevalence (K), and the explained variance of the PGS on the liability scale (*R*^2^_liability_). The case-control ratios in the training and testing samples were both simulated to be 50%, and the PGSs are on the standardized observed scale. Miscalibration is most apparent when the population prevalence is low and $R_{liability}^{2}$ is large. Error bars denote the standard error across 100 simulation runs.

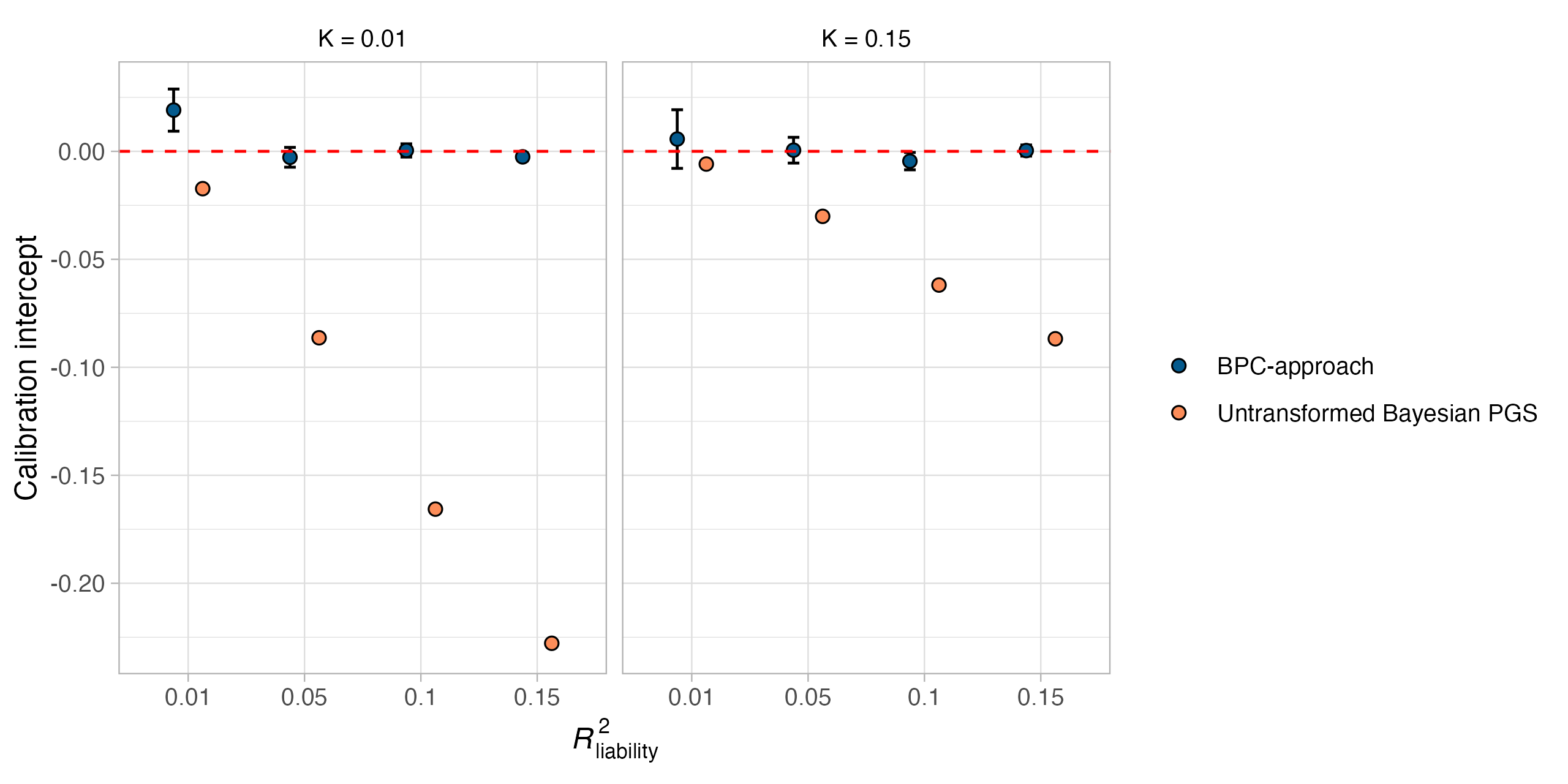

**Supplementary Figure 10 Miscalibration of untransformed Bayesian PGSs (intercept).**

Evaluating the calibration of untransformed Bayesian PGSs for binary traits using the intercept from a regression of the standardized disorder status on the PGS in 100 simulation runs and for combinations of two parameters, the population prevalence (K), and the explained variance of the PGS on the liability scale (*R*^2^_liability_). The case-control ratios in the training and testing samples were both simulated to be 50%, and the PGSs are on the standardized observed scale. Miscalibration is most apparent when the population prevalence is low and $R_{liability}^{2}$ is large. Error bars denote the standard error across 100 simulation runs. We note the scale of the PGS, which differs between the BPC approach (ranges from 0 to 1) and the untransformed Bayesian PGS (ranges from -1 to 1), affects the variance of the intercept but does not affect the interpretation of the presented results.

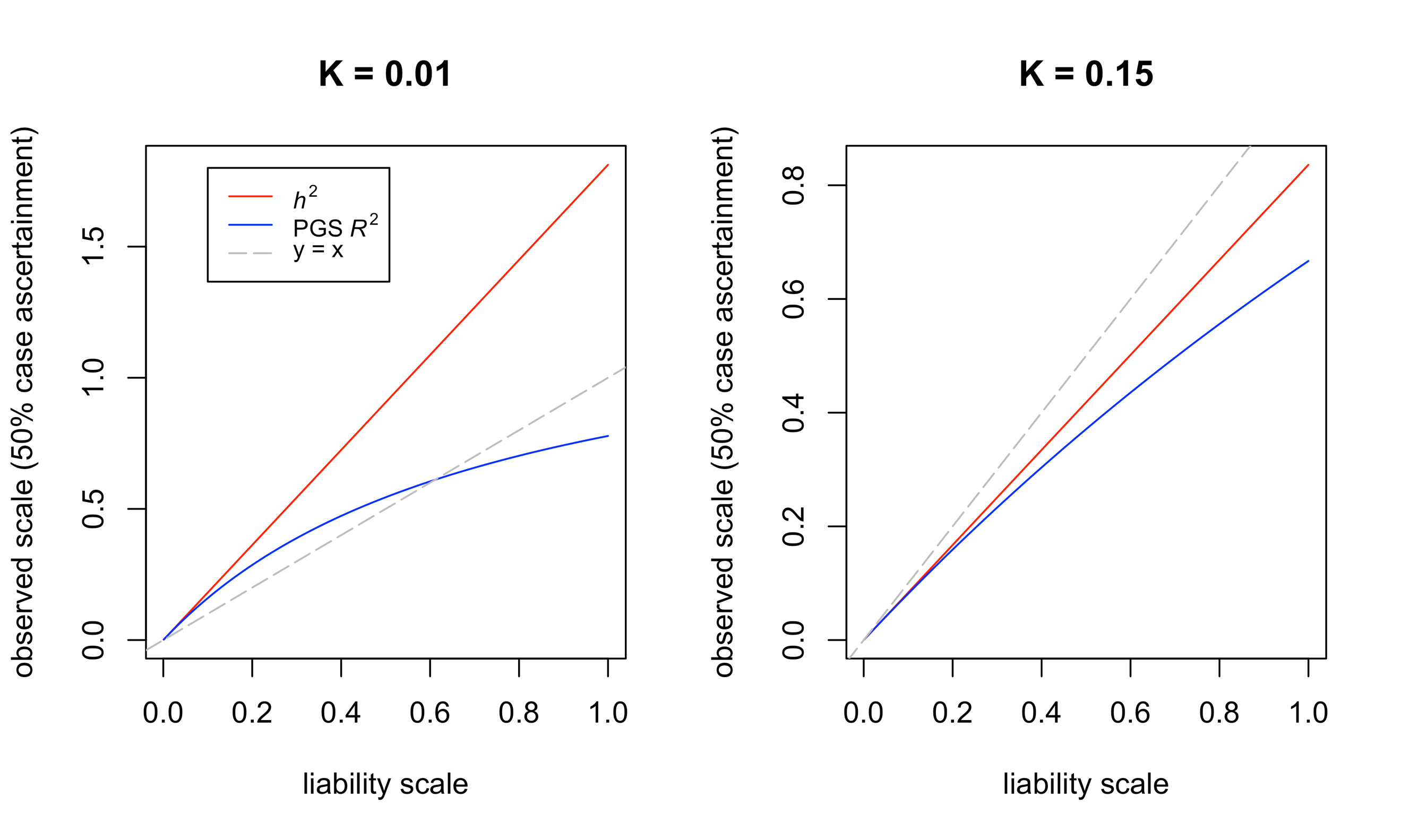

**Supplementary Figure 11 The transformation from the liability to the observed scale differs between *h*^2^ and *R*^2^.**

Depicted is the transformation of the heritability (*h*^2^; which is equivalent to the transformation of the betas)^13^ and the coefficient of determination (*R*^2^) from the liability to the observed scale with 50% case ascertainment. While the transformation from the liability to the observed scale in ascertained samples is linear for *h*^2^ and the GWAS results used to compute the PGS, it is non-linear for the *R*^2^ of the PGS. As a result, $var(PGS_{observed})$ and $R_{observed scale}^{2}$ are not proportional, and the PGS can thus not be well-calibrated without a probability conversion approach.

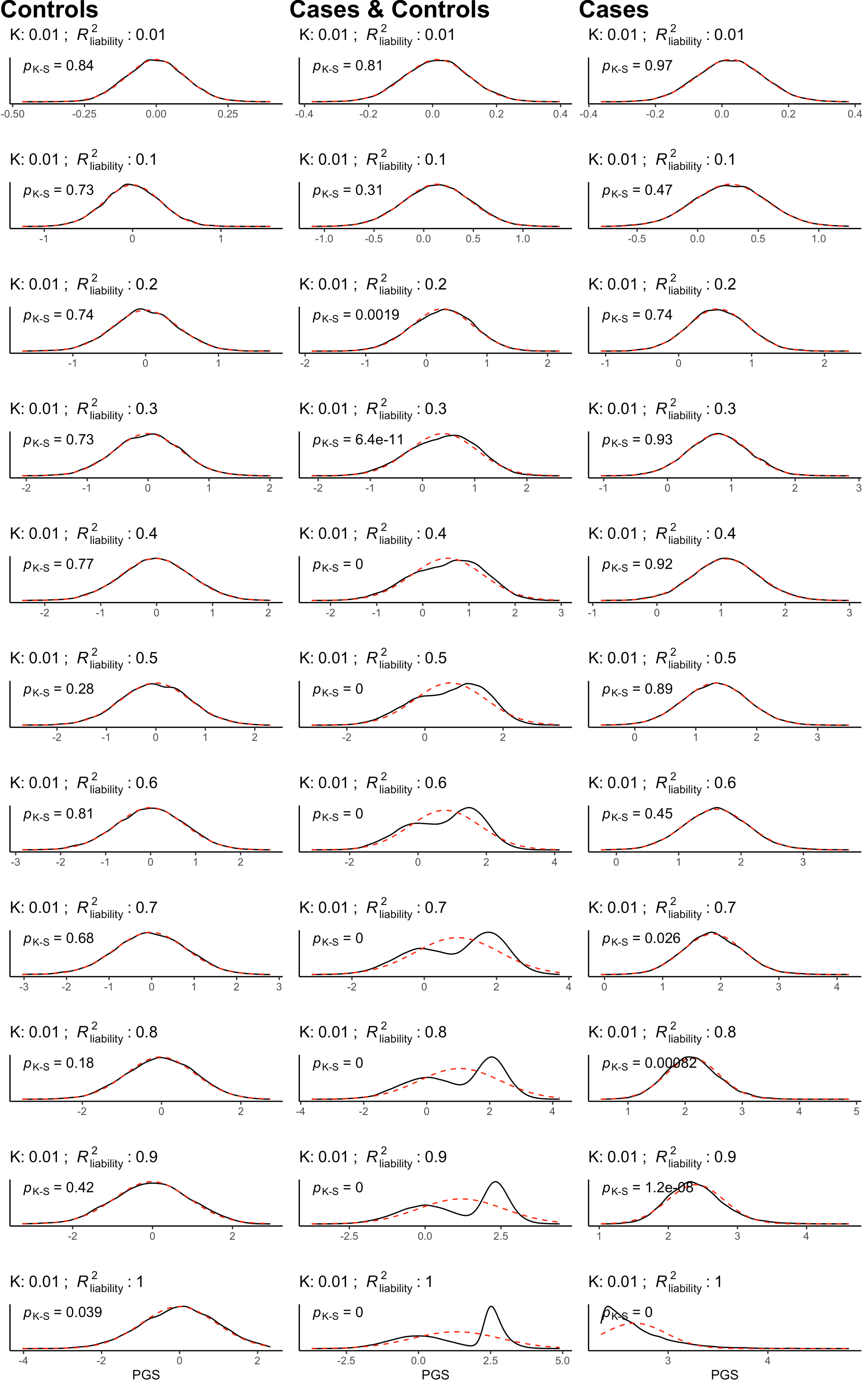

**Supplementary Figure 12 Normality of PGSs in simulations (K = 0.01). Panel A.**

Evaluating the normality of PGSs in only cases, only controls and in cases and controls. The BPC approach only assumes normality of PGSs in cases and controls separately; the combined distribution is shown for completeness, but we emphasize that BPC does not depend on the normality in this combined sample. Normality only starts to be violated at currently unrealistically large values of $R_{liability}^{2}$ (≥ 0.7). Normality is evaluated with the Kolmogorov-Smirnov test.

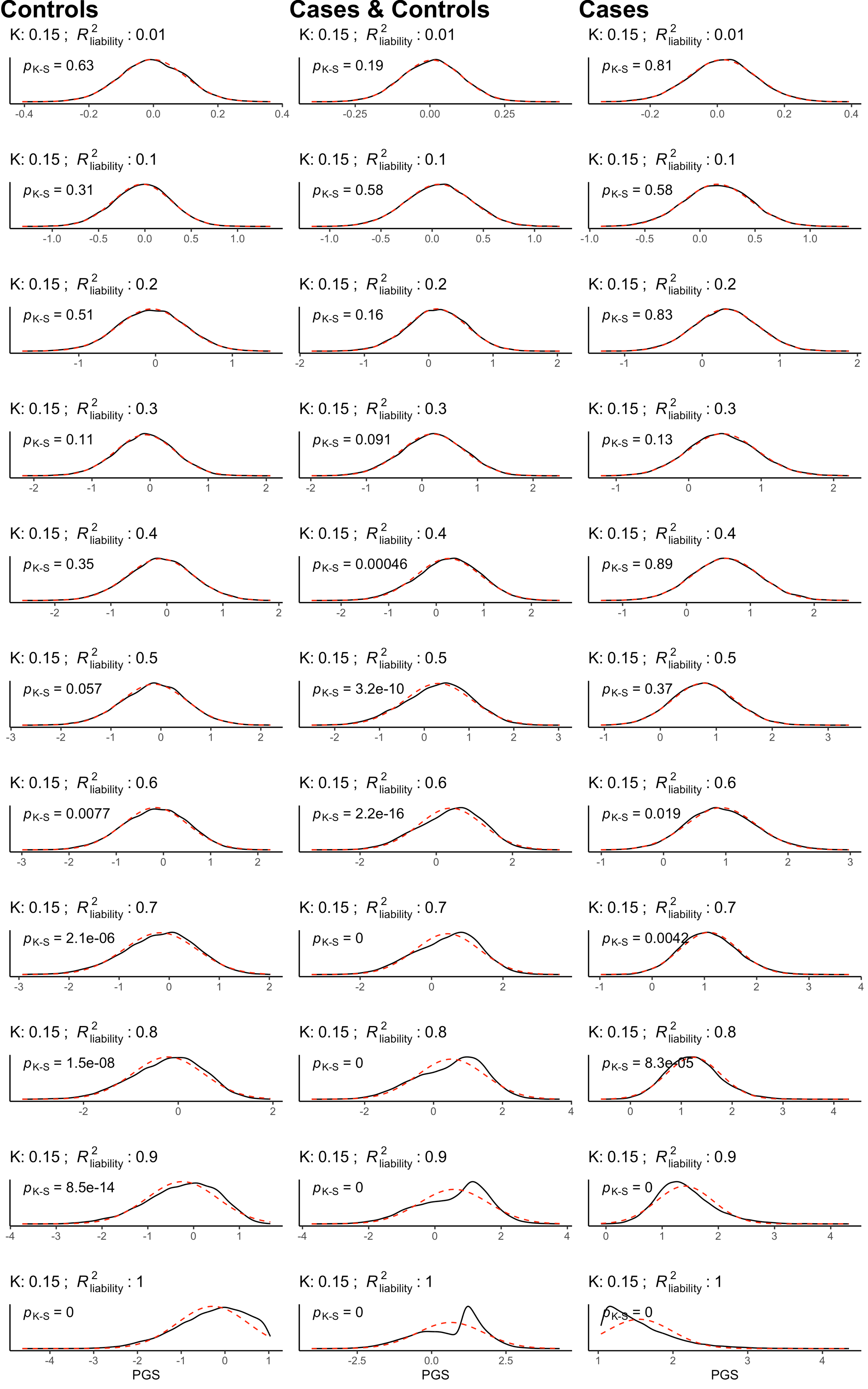

**Supplementary Figure 12 Normality of PGSs in simulations (K = 0.15). Panel A.**

Evaluating the normality of PGSs in only cases, only controls and in cases and controls. The BPC approach only assumes normality of PGSs in cases and controls separately; the combined distribution is shown for completeness, but we emphasize that BPC does not depend on the normality in this combined sample. Normality only starts to be violated at currently unrealistically large values of $R_{liability}^{2}$ (≥ 0.6). Normality is evaluated with the Kolmogorov-Smirnov test.

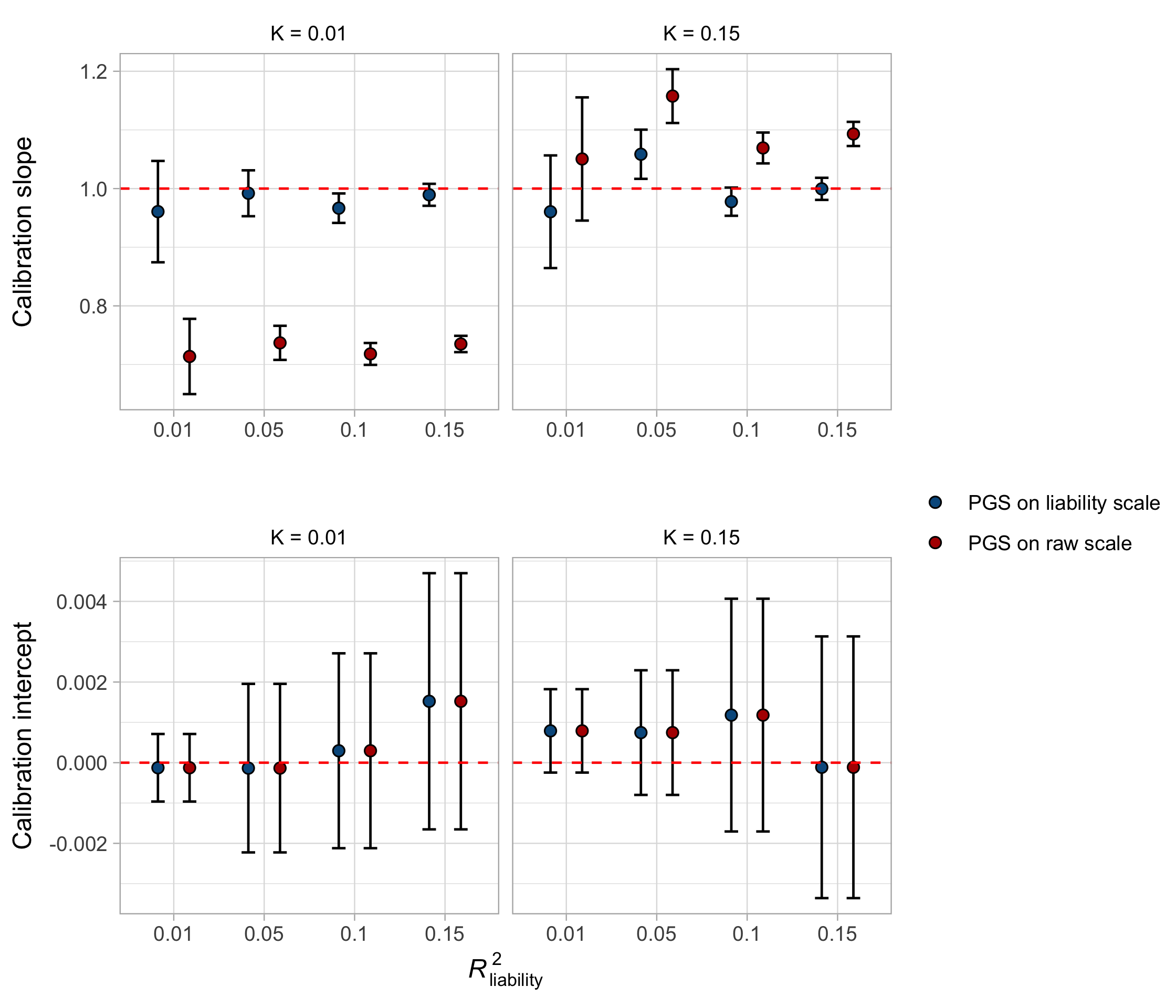

**Supplementary Figure 13 Calibration of PGSs on the liability scale in simulations using the calibration slope.**

Slopes and intercepts from regressing liability scores on PGSs on the liability and raw (i.e., untransformed) scale in a population reference sample. Error bars denote the 95% confidence intervals; The range of slopes expressed as 1.96 * S.D. is ten times larger than the error bars denoted here.

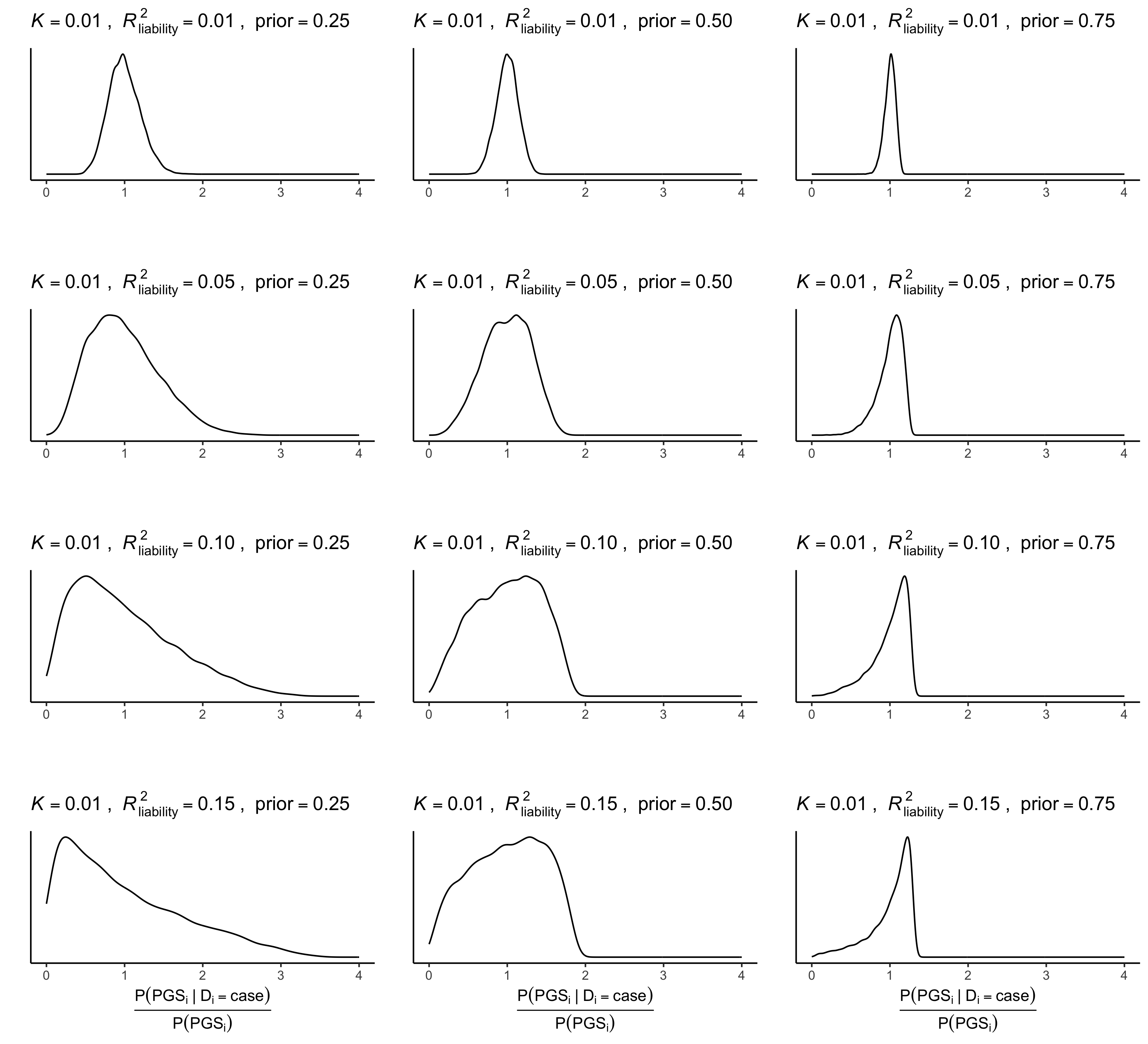

**Supplementary Figure 14 Panel A Distribution of** $\frac{\boldsymbol{P(}\boldsymbol{PGS}_{\boldsymbol{i}}\boldsymbol{|}\boldsymbol{D}_{\boldsymbol{i}}\boldsymbol{=case)}}{\boldsymbol{P(PG}\boldsymbol{S}_{\boldsymbol{i}}\boldsymbol{)}}$ **in simulations (K = 0.01). Panel A.**

To evaluate the extent to which the predicted disorder probabilities are dominated by the prior, we plotted the distribution of $\frac{P({PGS}_{i}|D_{i}=case)}{P(PGS_{i})}$ across all simulation parameters. The distribution varies markedly around one for most realistic simulation conditions, except when the $R_{liability}^{2}$ is very low (i.e., 1%) and the prior is very high (i.e., 75%).

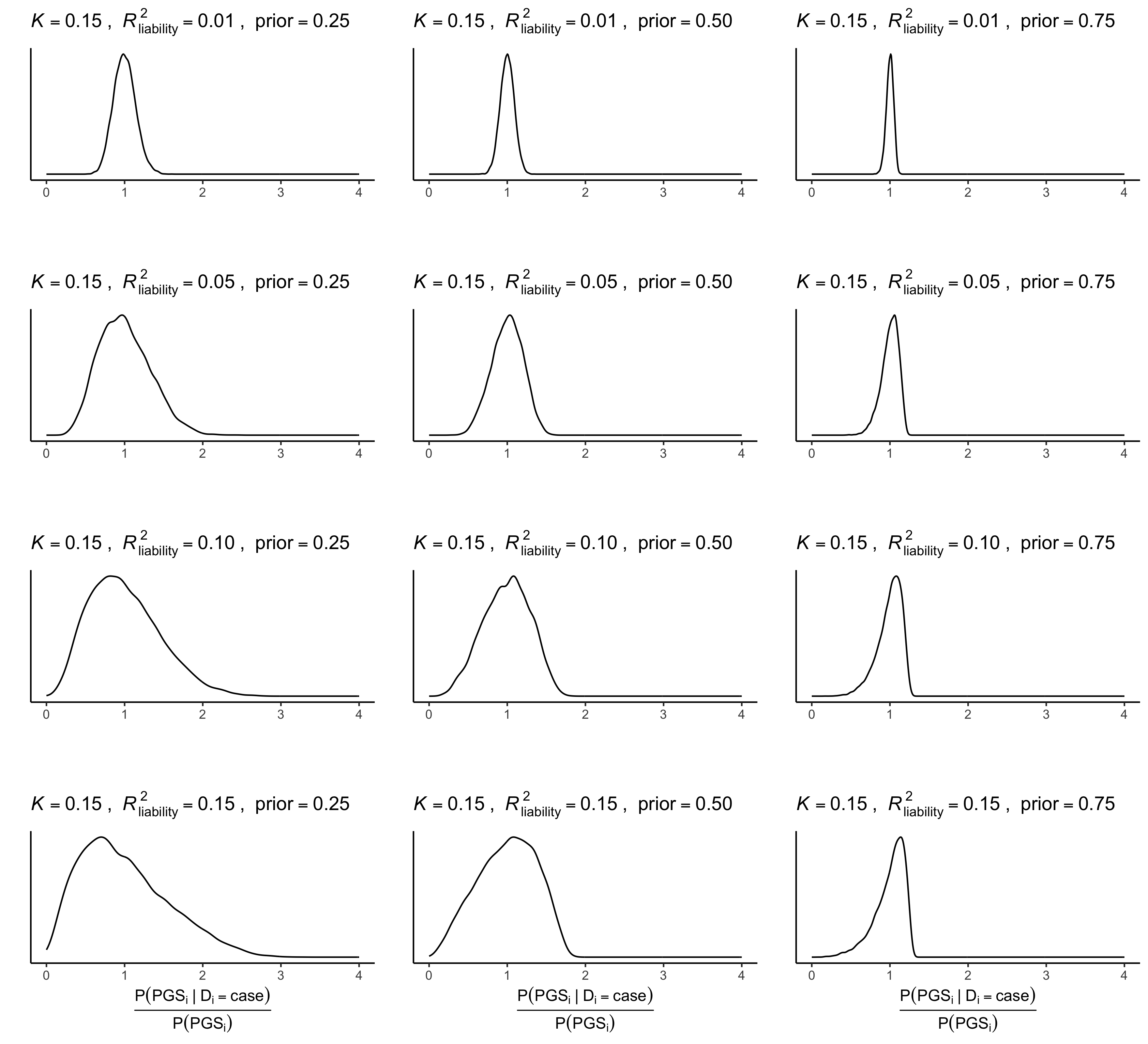

**Supplementary Figure 14 Distribution of** $\frac{\boldsymbol{P(}\boldsymbol{PGS}_{\boldsymbol{i}}\boldsymbol{|}\boldsymbol{D}_{\boldsymbol{i}}\boldsymbol{=case)}}{\boldsymbol{P(PG}\boldsymbol{S}_{\boldsymbol{i}}\boldsymbol{)}}$ **in simulations (K = 0.15). Panel B.**

To evaluate the extent to which the predicted disorder probabilities are dominated by the prior, we plotted the distribution of $\frac{P({PGS}_{i}|D_{i}=case)}{P(PGS_{i})}$ across all simulation parameters. The distribution varies markedly around one for most realistic simulation conditions, except when the $R_{liability}^{2}$ is very low (i.e., 1%) and the prior is very high (i.e., 75%).

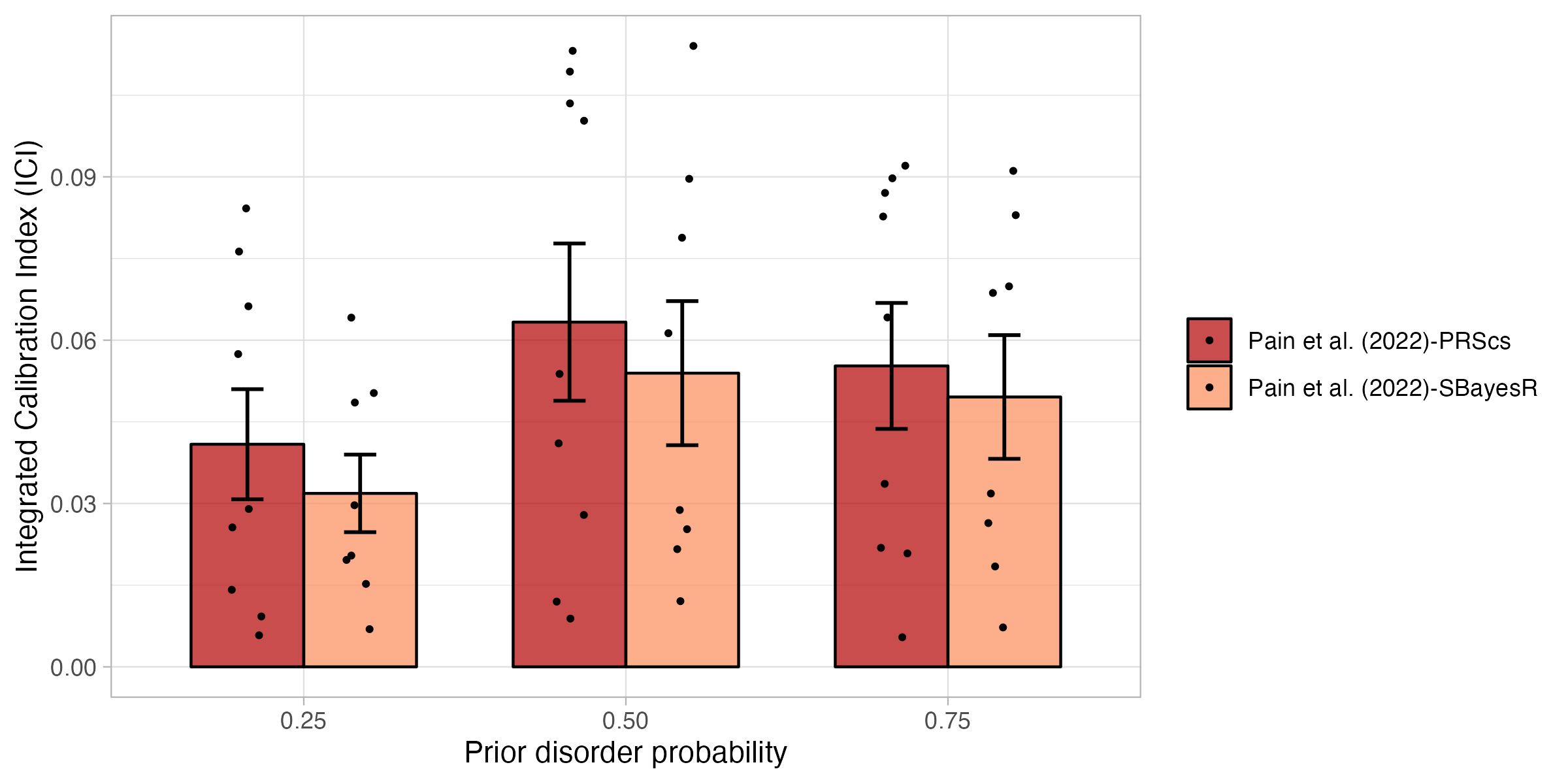

**Supplementary Figure 15 Calibration of Pain et al. (2022) using PRScs and SBayesR in empirical analyses of nine disorders.**

Calibration of the Pain et al. (2022) approach using PRScs and SBayesR was evaluated using the Integrated Calibration Index (ICI) for nine disorders while varying the prior disorder probability. The calibration of PRScs and SBayesR are very similar. Error bars denote the standard error.

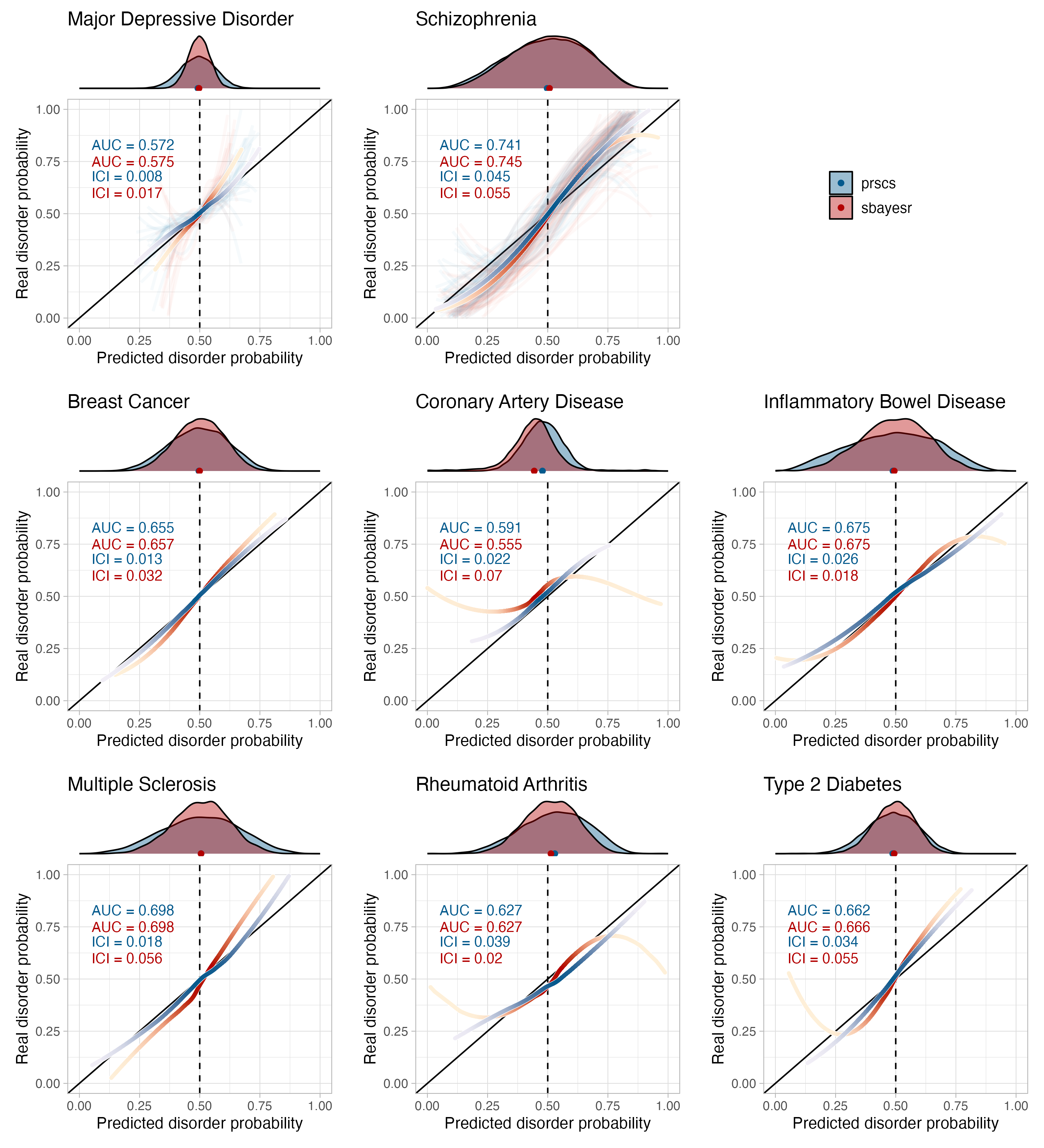

**Supplementary Figure 16 Disorder-specific calibration curves comparing BPC-PRScs and BPC-SBayesR in empirical analyses of nine disorders.**

Calibration of the BPC-PRScs and BPC-SBayesR approach was evaluated using the Integrated Calibration Index (ICI) for nine disorders, each with a prior disorder probability of 0.5. Histograms at the top of the plots depict the distribution of the predicted disorder probabilities, and the dots at the base of the histograms depict the mean predicted probability. The lines were drawn with a loess smoothing function, and their transparency follows the density of the histogram to show which parts of the distribution carry the most weight in the calculation of the ICI. For major depressive disorder and schizophrenia, several cohorts were available for analysis and therefore depict thin, light-colored, and transparent lines for individual cohorts. In contrast, the thicker and darker lines depict results when data from all cohorts was analyzed together. BPC-SBayesR makes correct predictions on average but is less well-calibrated for low and high values of the predicted disorder probabilities.

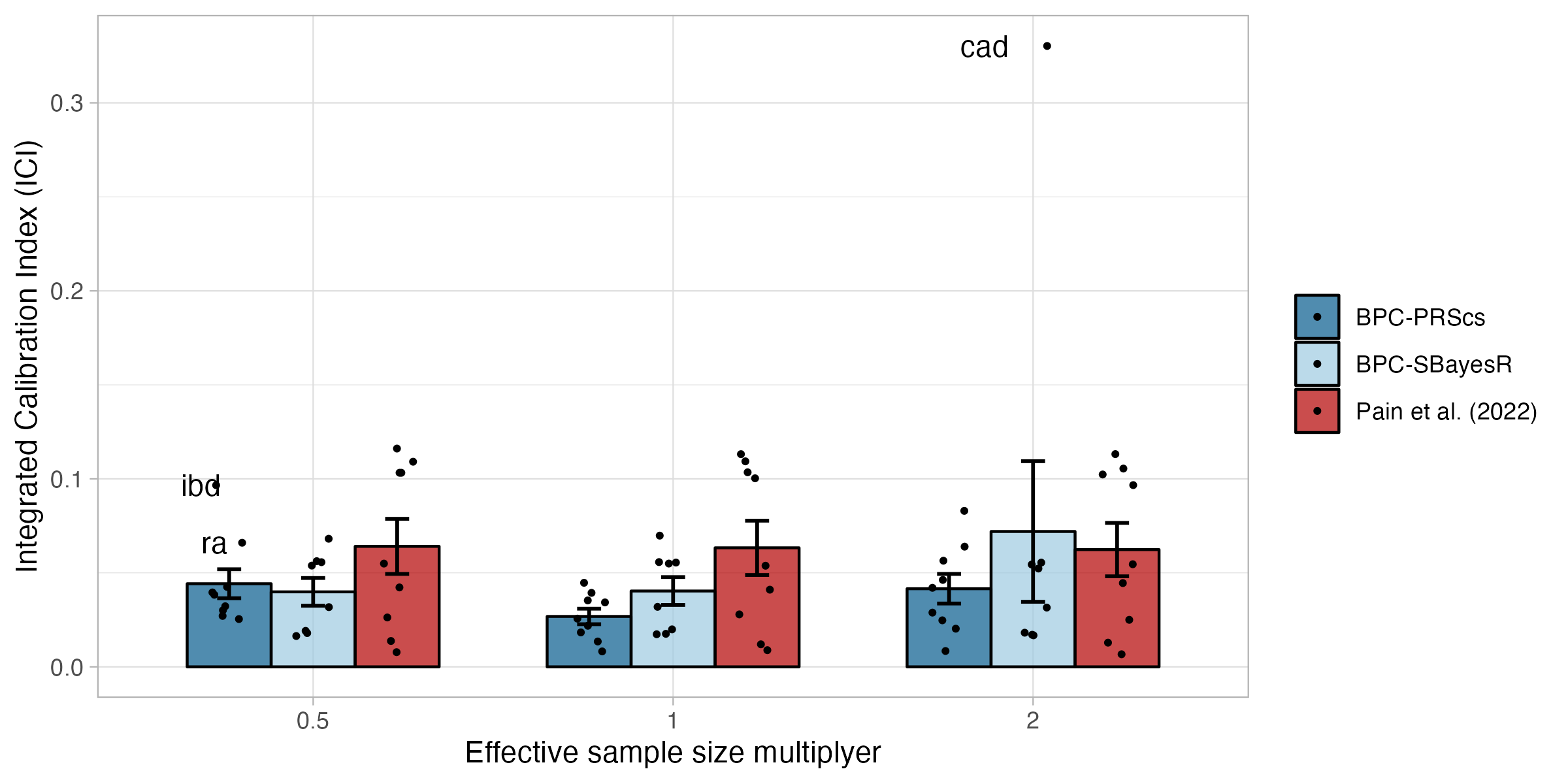

**Supplementary Figure 17 Calibration under misspecifications of the effective sample size in empirical analyses of nine disorders.**

Calibration of the BPC and the Pain et al. (2022) approach was evaluated using the Integrated Calibration Index (ICI) for nine disorders while misspecifying the effective sample size. The BPC approach was applied using two Bayesian PGS methods, PRScs (BPC-PRScs) and SBayesR (BPC-SBayesR). Misspecification of the effective sample size by a factor of 0.5 and 2 negatively impacts calibration for BPC-PRScs and for coronary artery disease for BPC-SBayesR, while it does not affect the calibration of the Pain et al. (2022) approach. The BPC approach still has lower mean ICI values than the Pain et al. (2022) approach in all conditions. Error bars denote the standard error.

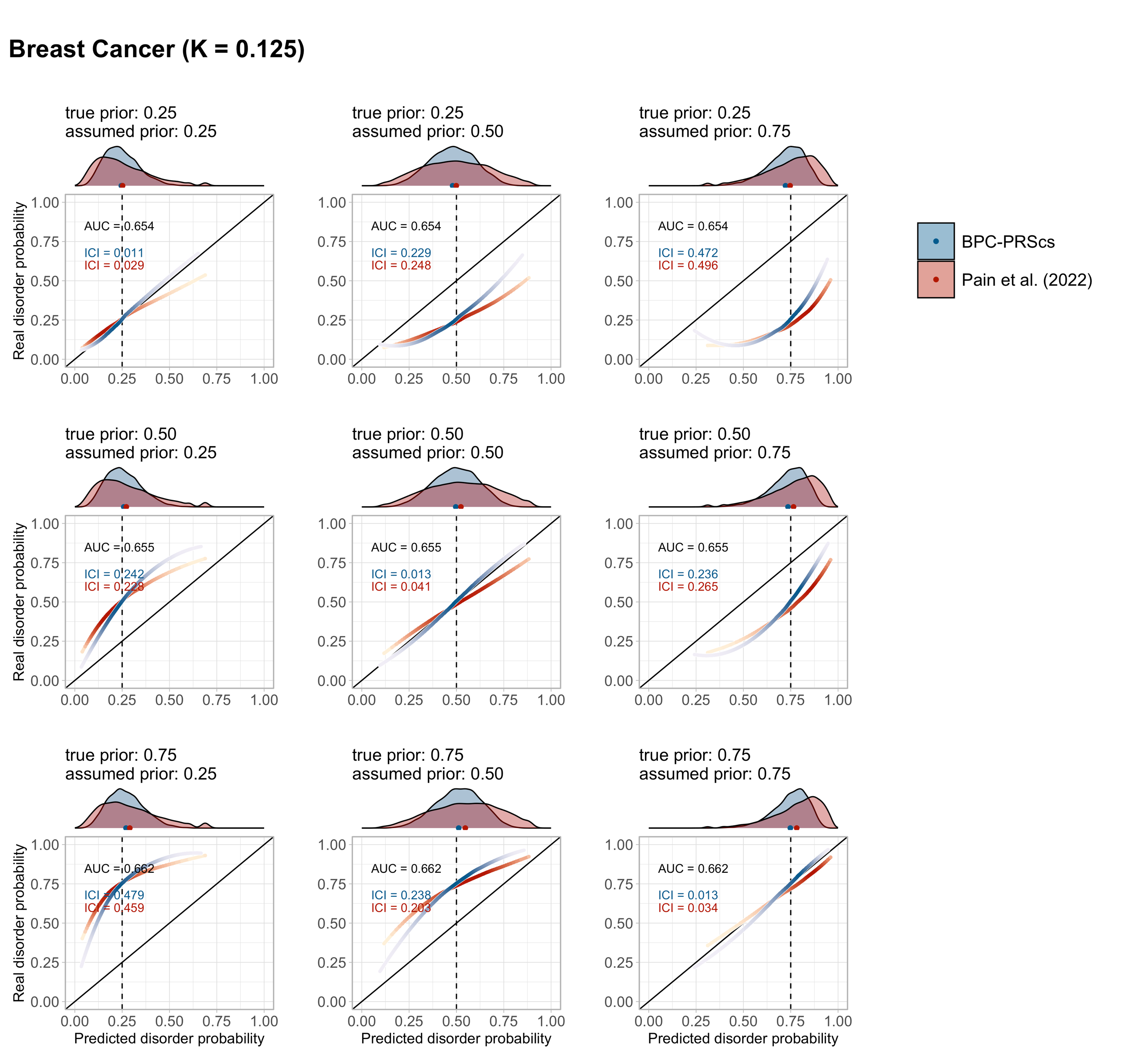

**Supplementary Figure 18 Misspecification of the prior in empirical analyses of Breast Cancer. Panel A.**

Calibration of the BPC and the Pain et al. (2022) approach was evaluated using the Integrated Calibration Index (ICI) for nine disorders. The true and assumed prior disorder probability varied between 0.25%, 0.50%, and 0.75%. Histograms at the top of the plots depict the distribution of the predicted disorder probabilities, and the dots at the base of the histograms depict the mean predicted probability. The lines were drawn with a loess smoothing function, and their transparency follows the density of the histogram to show which parts of the distribution carry the most weight in the calculation of the ICI. For major depression and schizophrenia, 62 and 22 cohorts, respectively, were available for analysis and therefore depict thin, light-colored, and transparent lines for individual cohorts. In contrast, the thicker and darker lines depict results when data from all cohorts are concatenated. The disorder population lifetime prevalence (K) is reported. The Area Under the receiver operator Curve (AUC) is the same for both approaches because the transformations do not change the ranking of individual PGSs, and both approaches use the same PGS inputs.

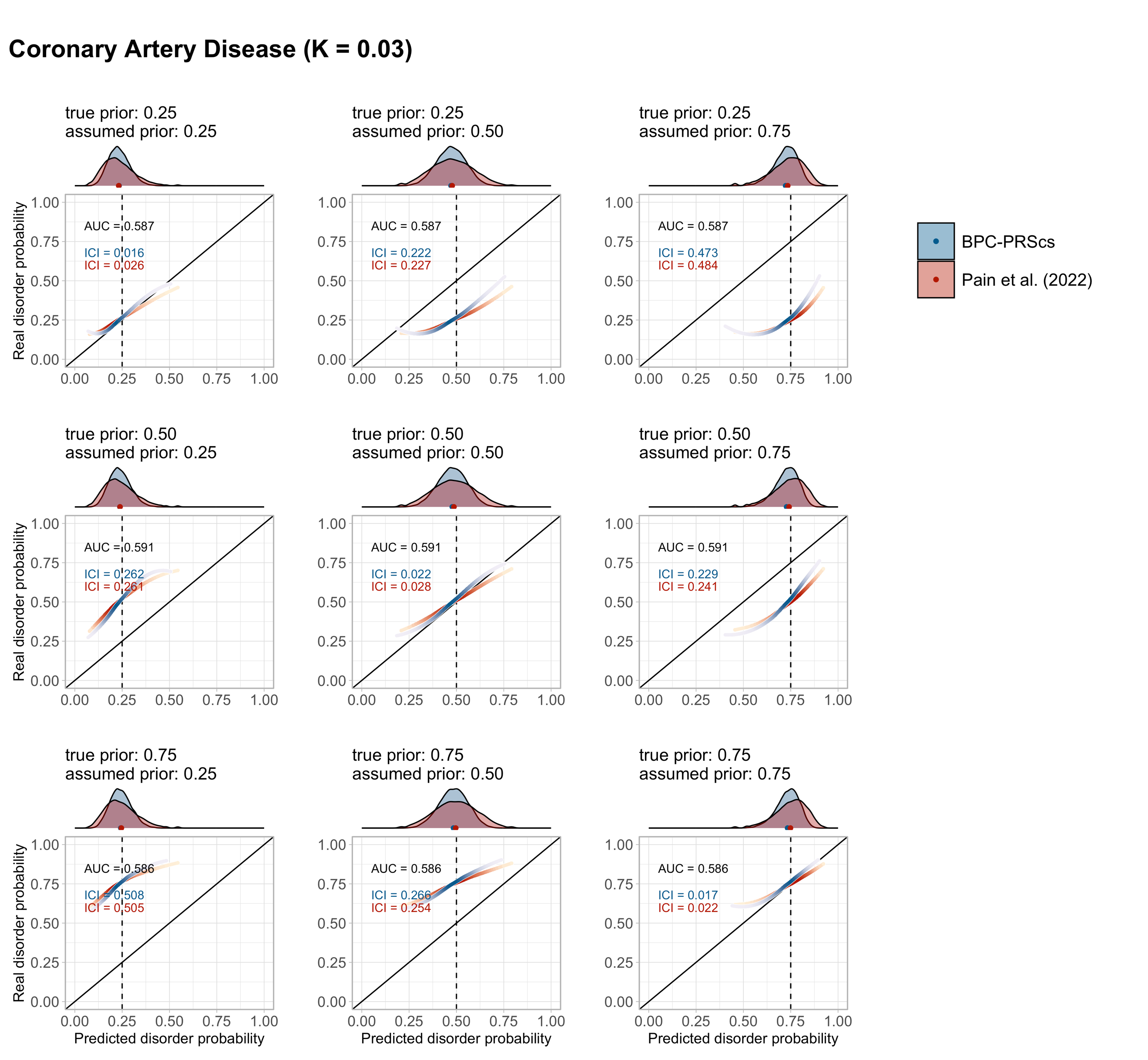

**Supplementary Figure 18 Misspecification of the prior in empirical analyses of Coronary Artery Disease. Panel B.**

Calibration of the BPC and the Pain et al. (2022) approach was evaluated using the Integrated Calibration Index (ICI) for nine disorders. The true and assumed prior disorder probability varied between 0.25%, 0.50%, and 0.75%. Histograms at the top of the plots depict the distribution of the predicted disorder probabilities, and the dots at the base of the histograms depict the mean predicted probability. The lines were drawn with a loess smoothing function, and their transparency follows the density of the histogram to show which parts of the distribution carry the most weight in the calculation of the ICI. For major depression and schizophrenia, 62 and 22 cohorts, respectively, were available for analysis and therefore depict thin, light-colored, and transparent lines for individual cohorts. In contrast, the thicker and darker lines depict results when data from all cohorts are concatenated. The disorder population lifetime prevalence (K) is reported. The Area Under the receiver operator Curve (AUC) is the same for both approaches because the transformations do not change the ranking of individual PGSs, and both approaches use the same PGS inputs.

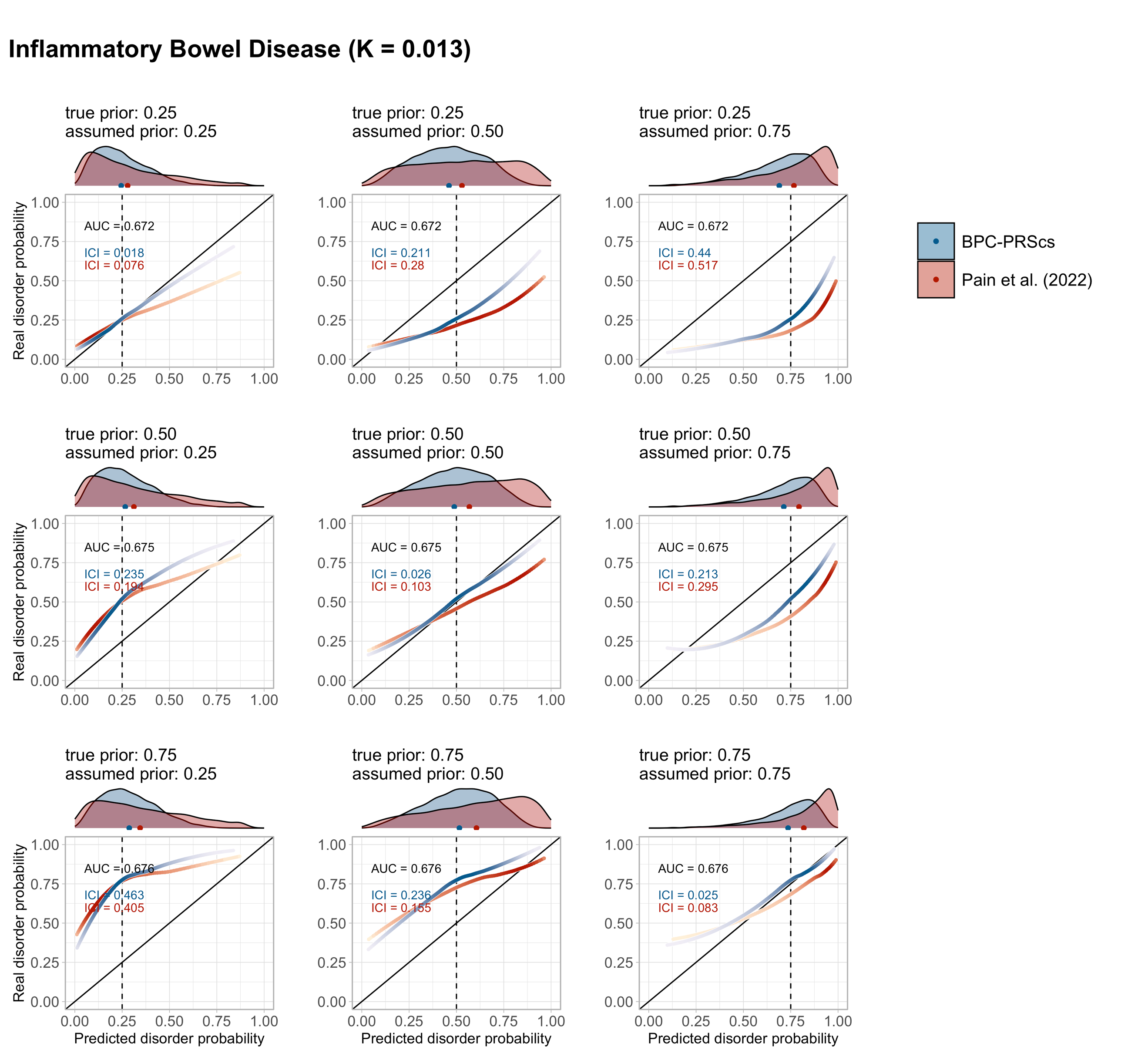

**Supplementary Figure 18 Misspecification of the prior in empirical analyses of Inflammatory Bowel Disease. Panel C.**

Calibration of the BPC and the Pain et al. (2022) approach was evaluated using the Integrated Calibration Index (ICI) for nine disorders. The true and assumed prior disorder probability varied between 0.25%, 0.50%, and 0.75%. Histograms at the top of the plots depict the distribution of the predicted disorder probabilities, and the dots at the base of the histograms depict the mean predicted probability. The lines were drawn with a loess smoothing function, and their transparency follows the density of the histogram to show which parts of the distribution carry the most weight in the calculation of the ICI. For major depression and schizophrenia, 62 and 22 cohorts, respectively, were available for analysis and therefore depict thin, light-colored, and transparent lines for individual cohorts. In contrast, the thicker and darker lines depict results when data from all cohorts are concatenated. The disorder population lifetime prevalence (K) is reported. The Area Under the receiver operator Curve (AUC) is the same for both approaches because the transformations do not change the ranking of individual PGSs, and both approaches use the same PGS inputs.

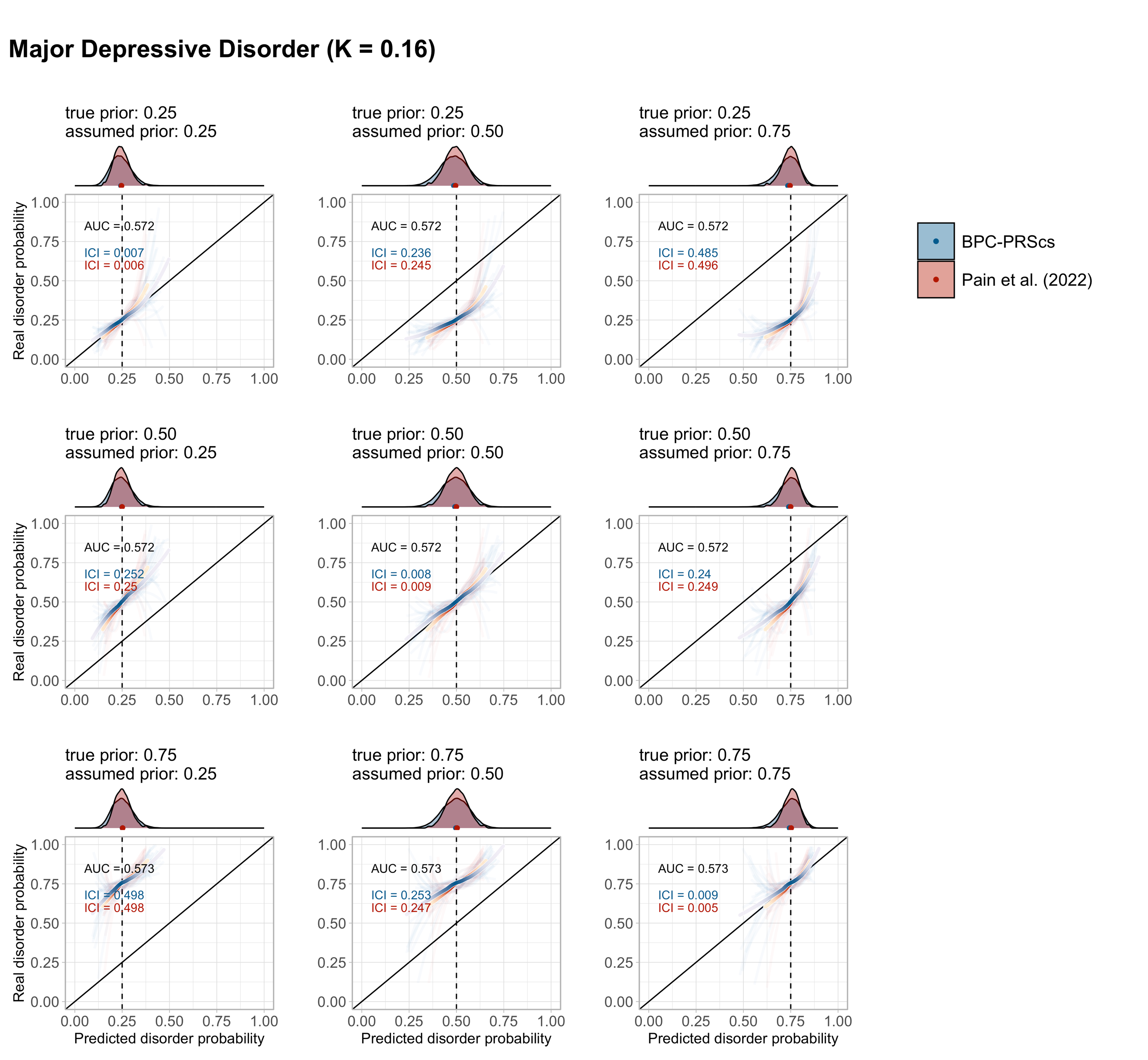

**Supplementary Figure 18 Misspecification of the prior in empirical analyses of Major Depressive Disorder. Panel D.**

Calibration of the BPC and the Pain et al. (2022) approach was evaluated using the Integrated Calibration Index (ICI) for nine disorders. The true and assumed prior disorder probability varied between 0.25%, 0.50%, and 0.75%. Histograms at the top of the plots depict the distribution of the predicted disorder probabilities, and the dots at the base of the histograms depict the mean predicted probability. The lines were drawn with a loess smoothing function, and their transparency follows the density of the histogram to show which parts of the distribution carry the most weight in the calculation of the ICI. For major depression and schizophrenia, 62 and 22 cohorts, respectively, were available for analysis and therefore depict thin, light-colored, and transparent lines for individual cohorts. In contrast, the thicker and darker lines depict results when data from all cohorts are concatenated. The disorder population lifetime prevalence (K) is reported. The Area Under the receiver operator Curve (AUC) is the same for both approaches because the transformations do not change the ranking of individual PGSs, and both approaches use the same PGS inputs.

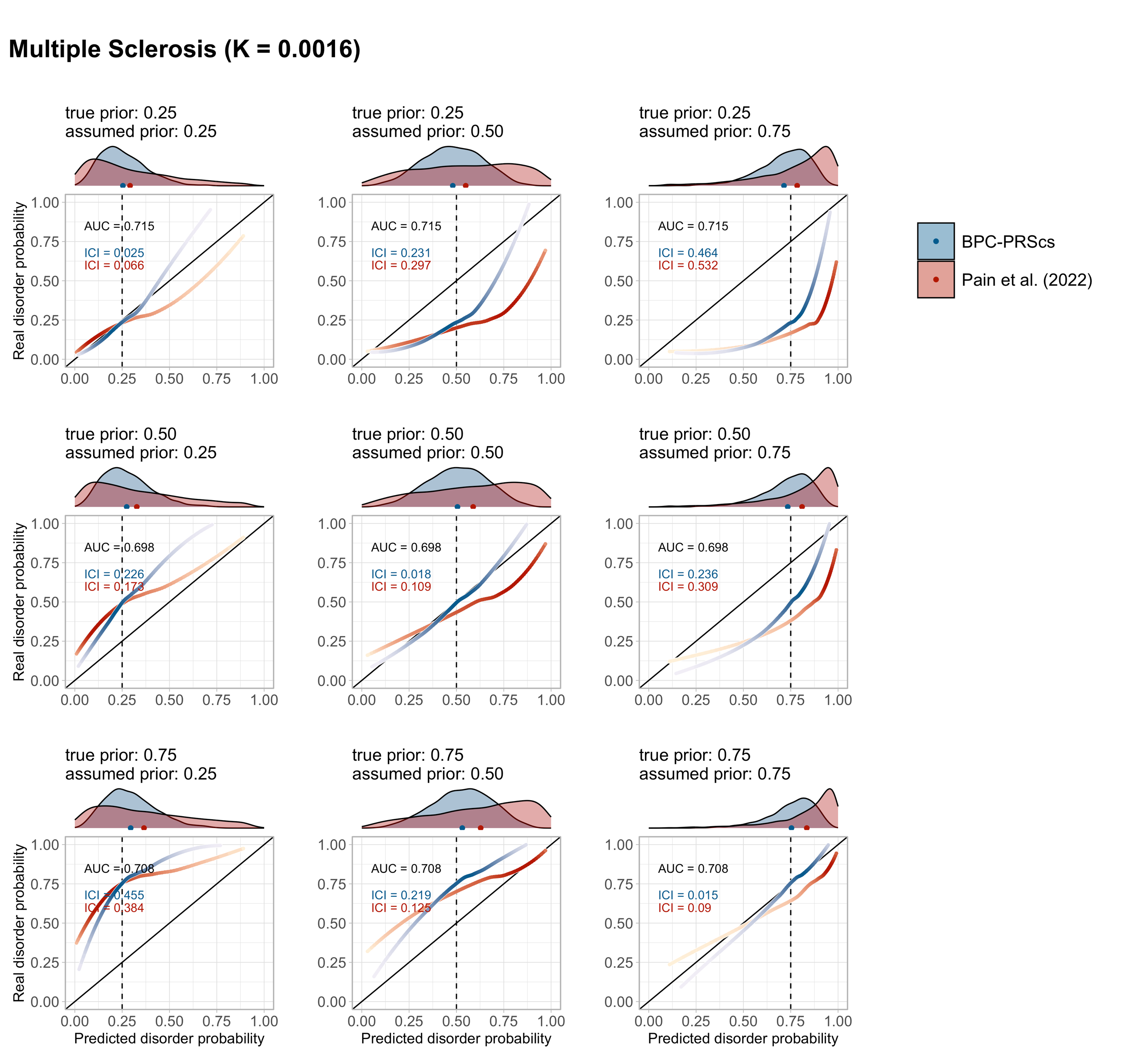

**Supplementary Figure 18 Misspecification of the prior in empirical analyses of Multiple Sclerosis. Panel E.**

Calibration of the BPC and the Pain et al. (2022) approach was evaluated using the Integrated Calibration Index (ICI) for nine disorders. The true and assumed prior disorder probability varied between 0.25%, 0.50%, and 0.75%. Histograms at the top of the plots depict the distribution of the predicted disorder probabilities, and the dots at the base of the histograms depict the mean predicted probability. The lines were drawn with a loess smoothing function, and their transparency follows the density of the histogram to show which parts of the distribution carry the most weight in the calculation of the ICI. For major depression and schizophrenia, 62 and 22 cohorts, respectively, were available for analysis and therefore depict thin, light-colored, and transparent lines for individual cohorts. In contrast, the thicker and darker lines depict results when data from all cohorts are concatenated. The disorder population lifetime prevalence (K) is reported. The Area Under the receiver operator Curve (AUC) is the same for both approaches because the transformations do not change the ranking of individual PGSs, and both approaches use the same PGS inputs.

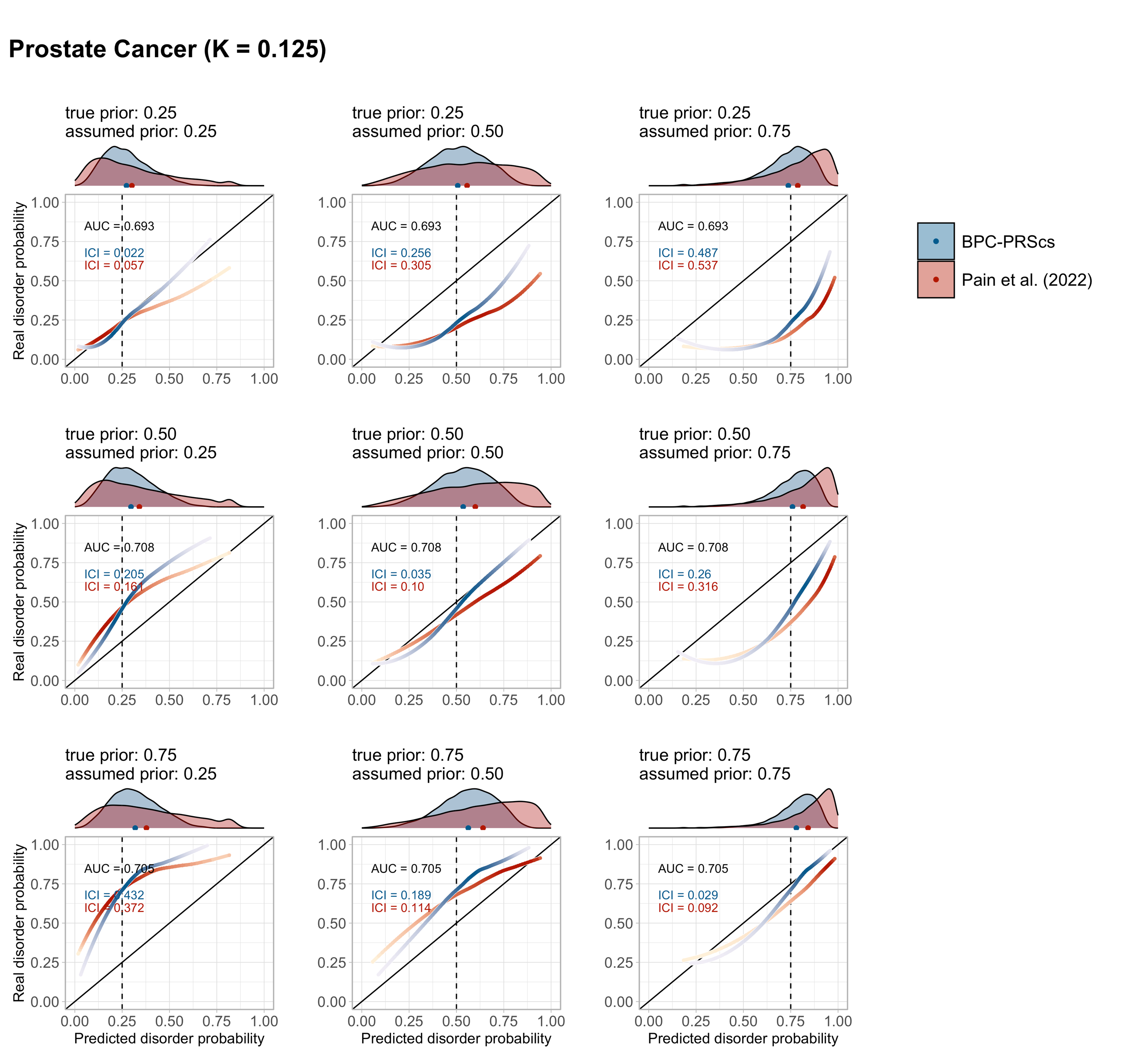

**Supplementary Figure 18 Misspecification of the prior in empirical analyses of Prostate Cancer. Panel F.**

Calibration of the BPC and the Pain et al. (2022) approach was evaluated using the Integrated Calibration Index (ICI) for nine disorders. The true and assumed prior disorder probability varied between 0.25%, 0.50%, and 0.75%. Histograms at the top of the plots depict the distribution of the predicted disorder probabilities, and the dots at the base of the histograms depict the mean predicted probability. The lines were drawn with a loess smoothing function, and their transparency follows the density of the histogram to show which parts of the distribution carry the most weight in the calculation of the ICI. For major depression and schizophrenia, 62 and 22 cohorts, respectively, were available for analysis and therefore depict thin, light-colored, and transparent lines for individual cohorts. In contrast, the thicker and darker lines depict results when data from all cohorts are concatenated. The disorder population lifetime prevalence (K) is reported. The Area Under the receiver operator Curve (AUC) is the same for both approaches because the transformations do not change the ranking of individual PGSs, and both approaches use the same PGS inputs.

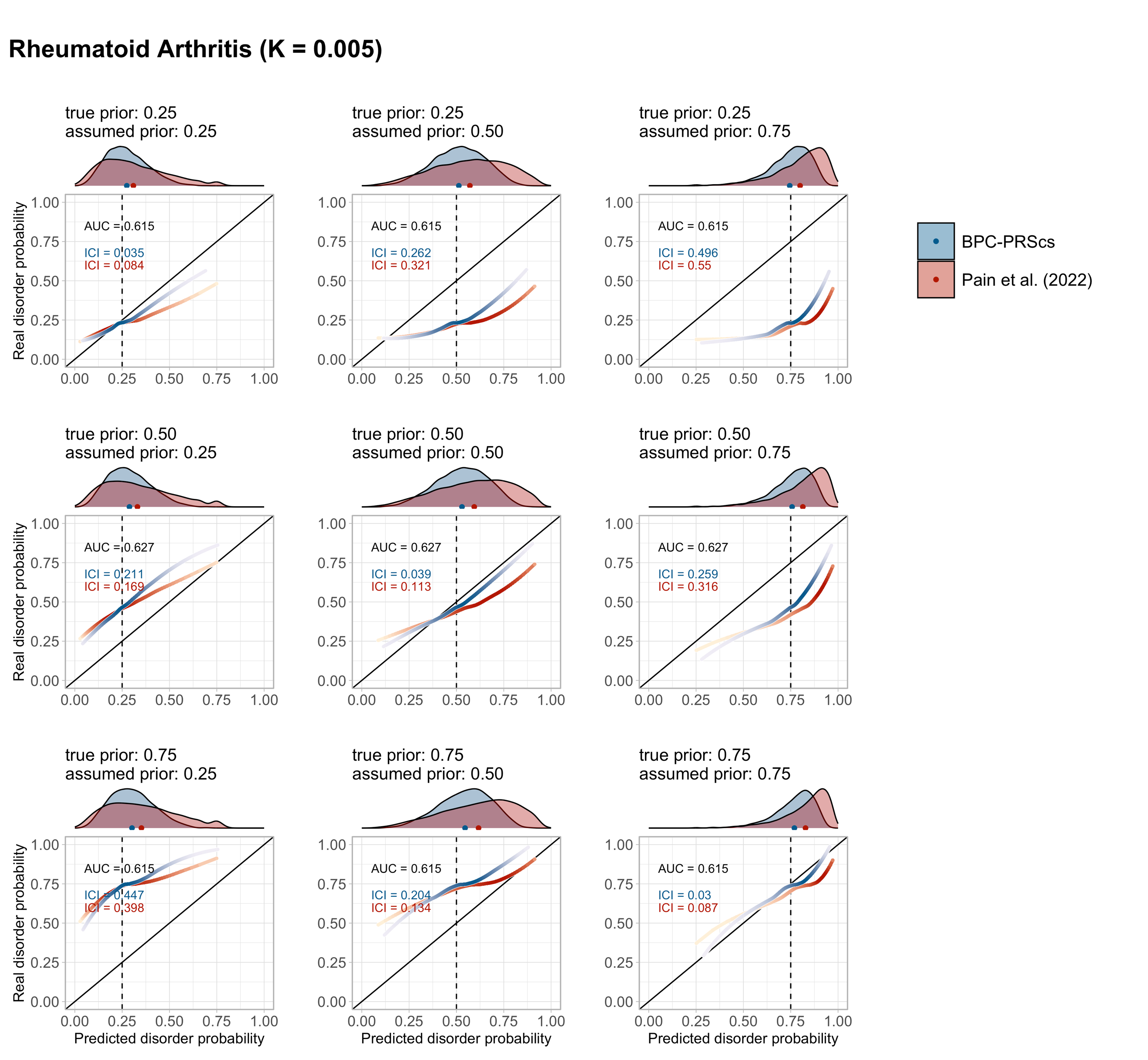

**Supplementary Figure 18 Misspecification of the prior in empirical analyses of Rheumatoid Arthritis. Panel G.**

Calibration of the BPC and the Pain et al. (2022) approach was evaluated using the Integrated Calibration Index (ICI) for nine disorders. The true and assumed prior disorder probability varied between 0.25%, 0.50%, and 0.75%. Histograms at the top of the plots depict the distribution of the predicted disorder probabilities, and the dots at the base of the histograms depict the mean predicted probability. The lines were drawn with a loess smoothing function, and their transparency follows the density of the histogram to show which parts of the distribution carry the most weight in the calculation of the ICI. For major depression and schizophrenia, 62 and 22 cohorts, respectively, were available for analysis and therefore depict thin, light-colored, and transparent lines for individual cohorts. In contrast, the thicker and darker lines depict results when data from all cohorts are concatenated. The disorder population lifetime prevalence (K) is reported. The Area Under the receiver operator Curve (AUC) is the same for both approaches because the transformations do not change the ranking of individual PGSs, and both approaches use the same PGS inputs.

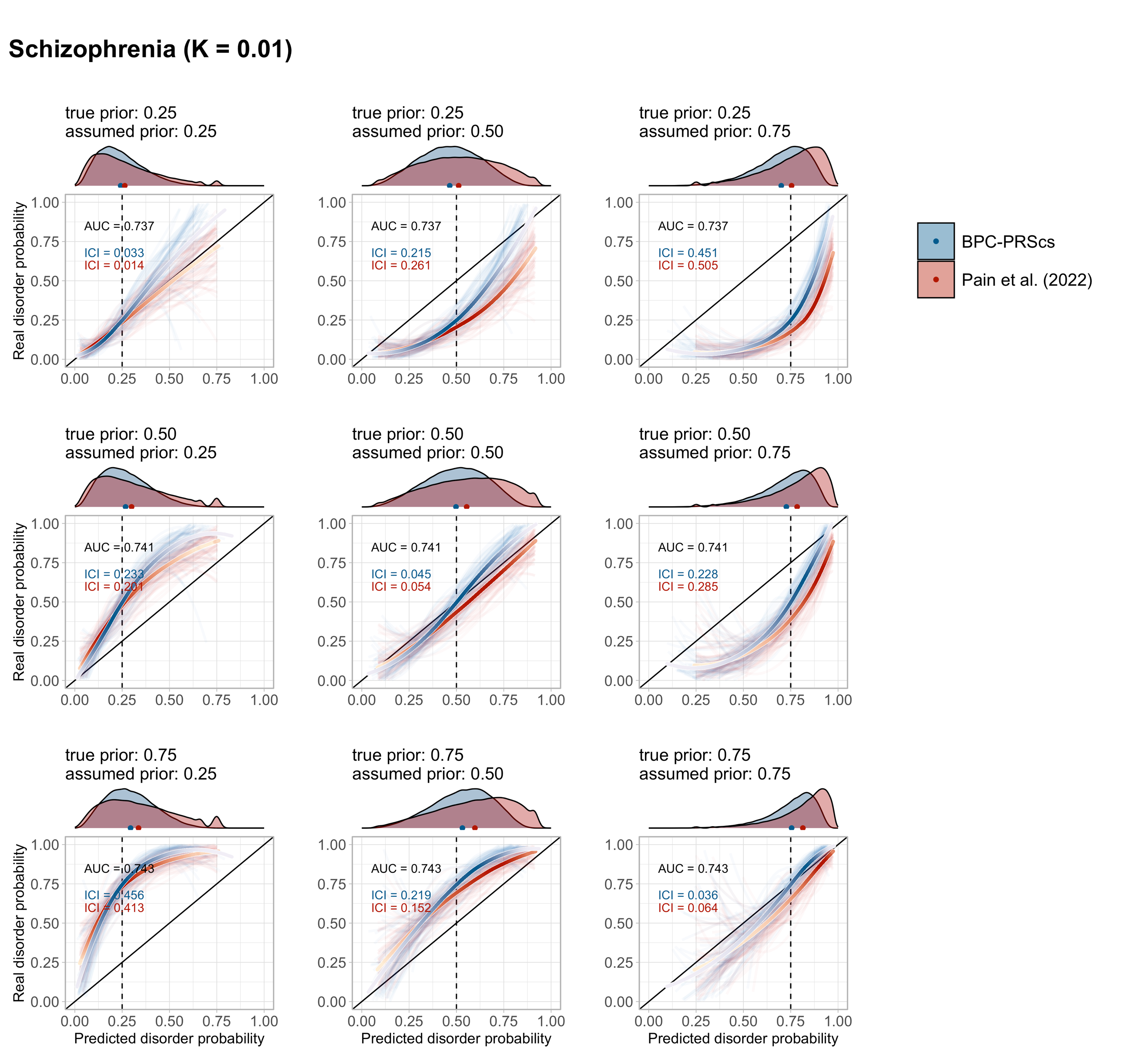

**Supplementary Figure 18 Misspecification of the prior in empirical analyses of Schizophrenia. Panel H.**

Calibration of the BPC and the Pain et al. (2022) approach was evaluated using the Integrated Calibration Index (ICI) for nine disorders. The true and assumed prior disorder probability varied between 0.25%, 0.50%, and 0.75%. Histograms at the top of the plots depict the distribution of the predicted disorder probabilities, and the dots at the base of the histograms depict the mean predicted probability. The lines were drawn with a loess smoothing function, and their transparency follows the density of the histogram to show which parts of the distribution carry the most weight in the calculation of the ICI. For major depression and schizophrenia, 62 and 22 cohorts, respectively, were available for analysis and therefore depict thin, light-colored, and transparent lines for individual cohorts. In contrast, the thicker and darker lines depict results when data from all cohorts are concatenated. The disorder population lifetime prevalence (K) is reported. The Area Under the receiver operator Curve (AUC) is the same for both approaches because the transformations do not change the ranking of individual PGSs, and both approaches use the same PGS inputs.

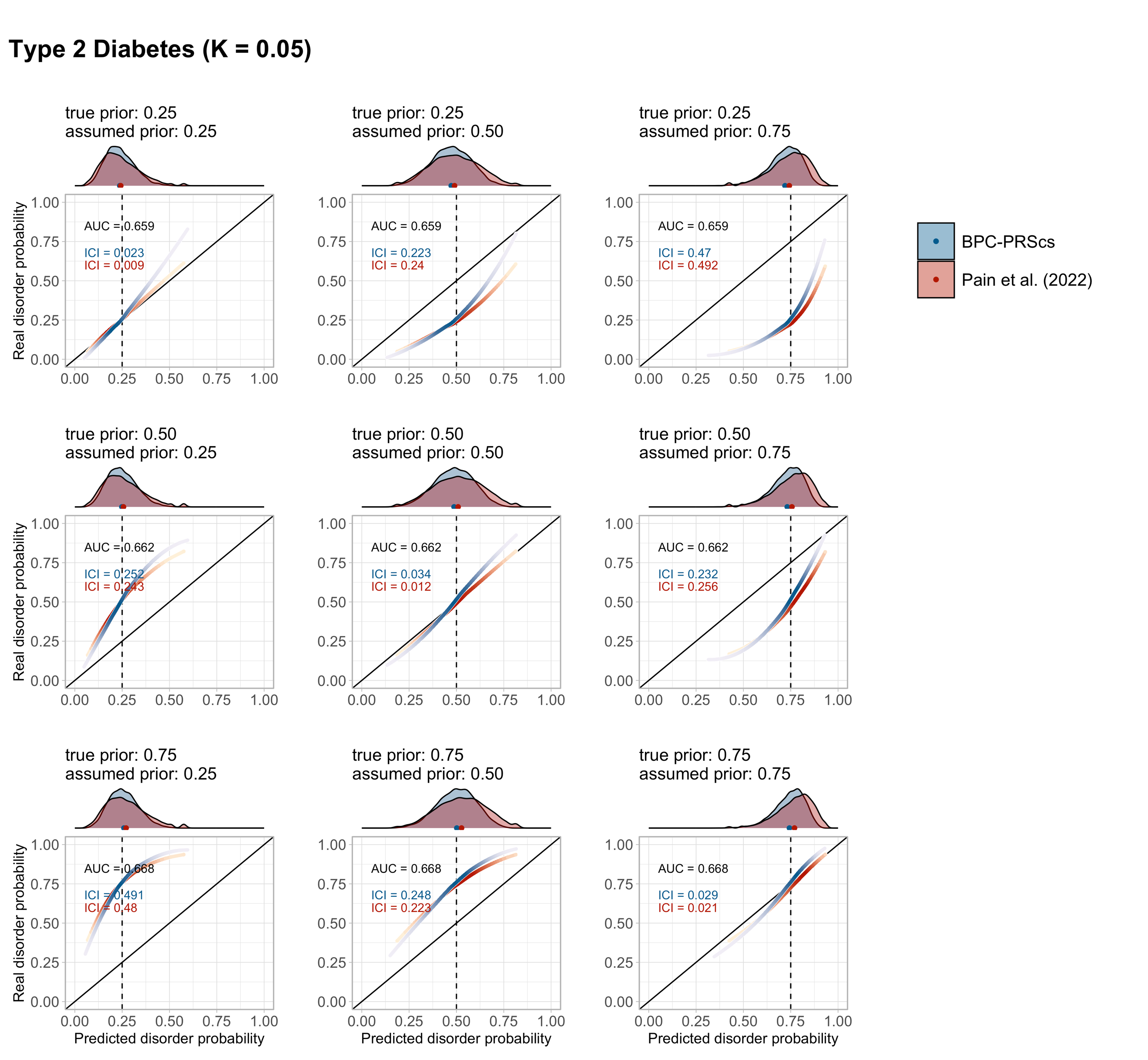

**Supplementary Figure 18 Misspecification of the prior in empirical analyses of Type 2 Diabetes. Panel I.**

Calibration of the BPC and the Pain et al. (2022) approach was evaluated using the Integrated Calibration Index (ICI) for nine disorders. The true and assumed prior disorder probability varied between 0.25%, 0.50%, and 0.75%. Histograms at the top of the plots depict the distribution of the predicted disorder probabilities, and the dots at the base of the histograms depict the mean predicted probability. The lines were drawn with a loess smoothing function, and their transparency follows the density of the histogram to show which parts of the distribution carry the most weight in the calculation of the ICI. For major depression and schizophrenia, 62 and 22 cohorts, respectively, were available for analysis and therefore depict thin, light-colored, and transparent lines for individual cohorts. In contrast, the thicker and darker lines depict results when data from all cohorts are concatenated. The disorder population lifetime prevalence (K) is reported. The Area Under the receiver operator Curve (AUC) is the same for both approaches because the transformations do not change the ranking of individual PGSs, and both approaches use the same PGS inputs.

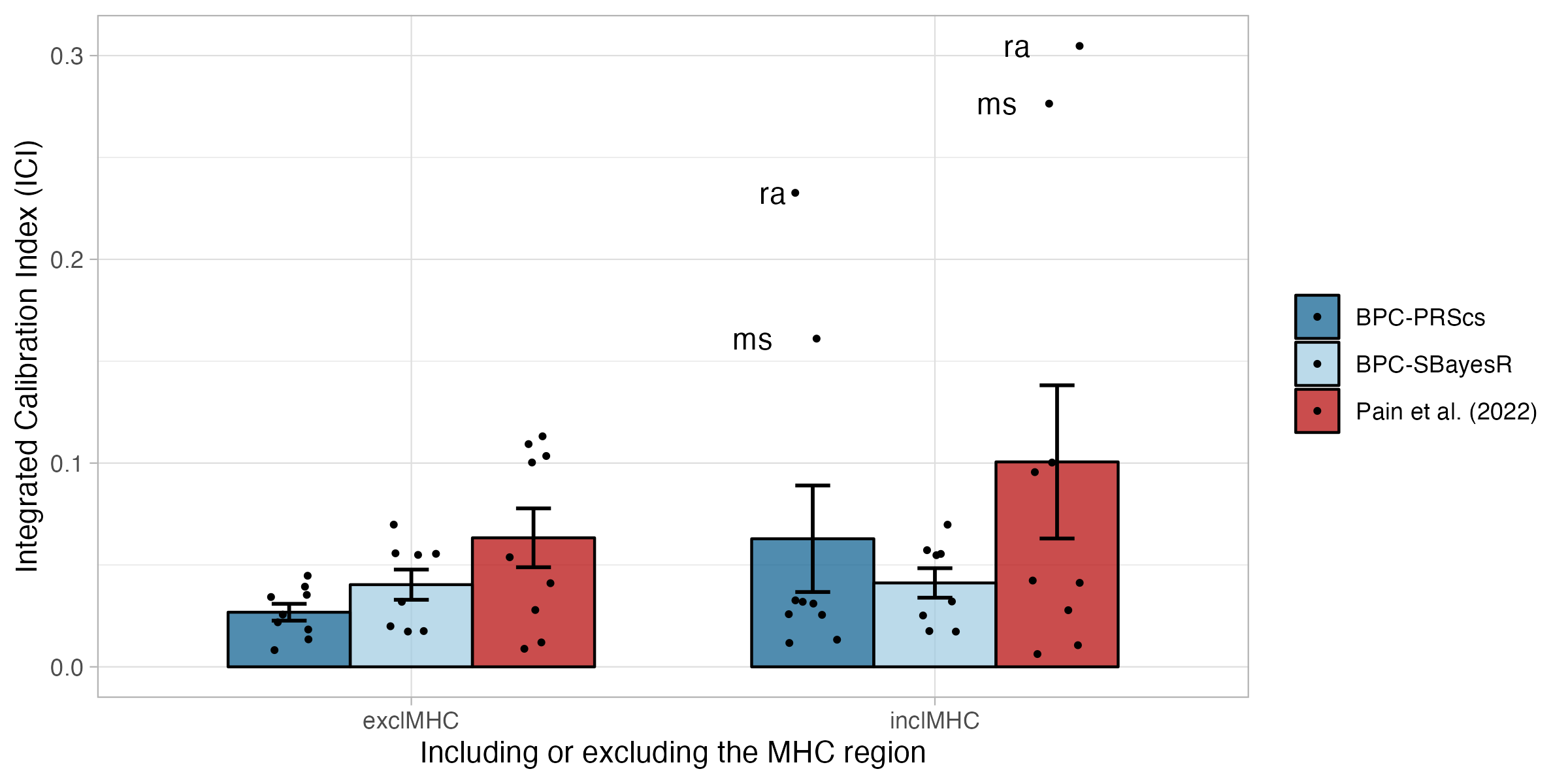

**Supplementary Figure 19 Effect of inclusion of the MHC region on calibration in empirical analyses of nine disorders.**

Calibration of the BPC and the Pain et al. (2022) approach was evaluated using the Integrated Calibration Index (ICI) for nine disorders while excluding and including the MHC region. The BPC approach was applied using two Bayesian PGS methods, PRScs (BPC-PRScs) and SBayesR (BPC-SBayesR). Including the MHC region (hg19 coordinates: 6:28000000:34000000) strongly and negatively impacts calibration for autoimmune disorders. Calibration of BPC-SBayesR is not affected, because SBayesR’s reference files exclude most of the MHC region by default. exclMHC = excluding the MHC region; inclMHC = including the MHC region. Error bars denote the standard error.

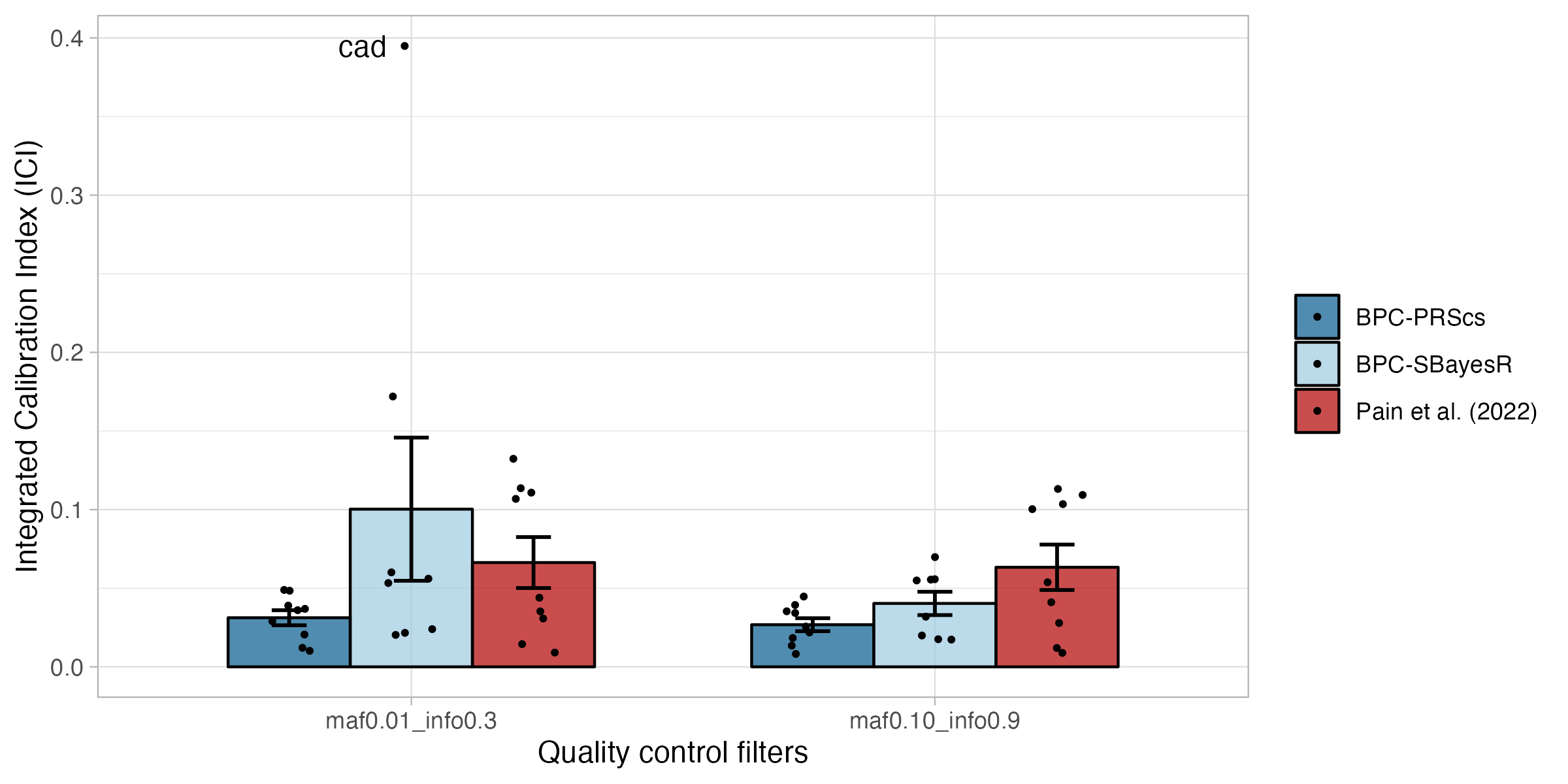

**Supplementary Figure 20 Effect of quality control thresholds on calibration in empirical analyses of nine disorders.**

Calibration of the BPC and the Pain et al. (2022) approach was evaluated using the Integrated Calibration Index (ICI) for nine disorders while varying the quality control filters. The BPC approach was applied using two Bayesian PGS methods, PRScs (BPC-PRScs) and SBayesR (BPC-SBayesR). Reducing the INFO filter from 0.9 to 0.3 and the minor allele frequency (MAF) filter from 10% to 1% slightly increases the average ICI for BPC-PRScs and strongly increases the ICI for Coronary artery disease for BPC-SBayesR. Error bars denote the standard error.

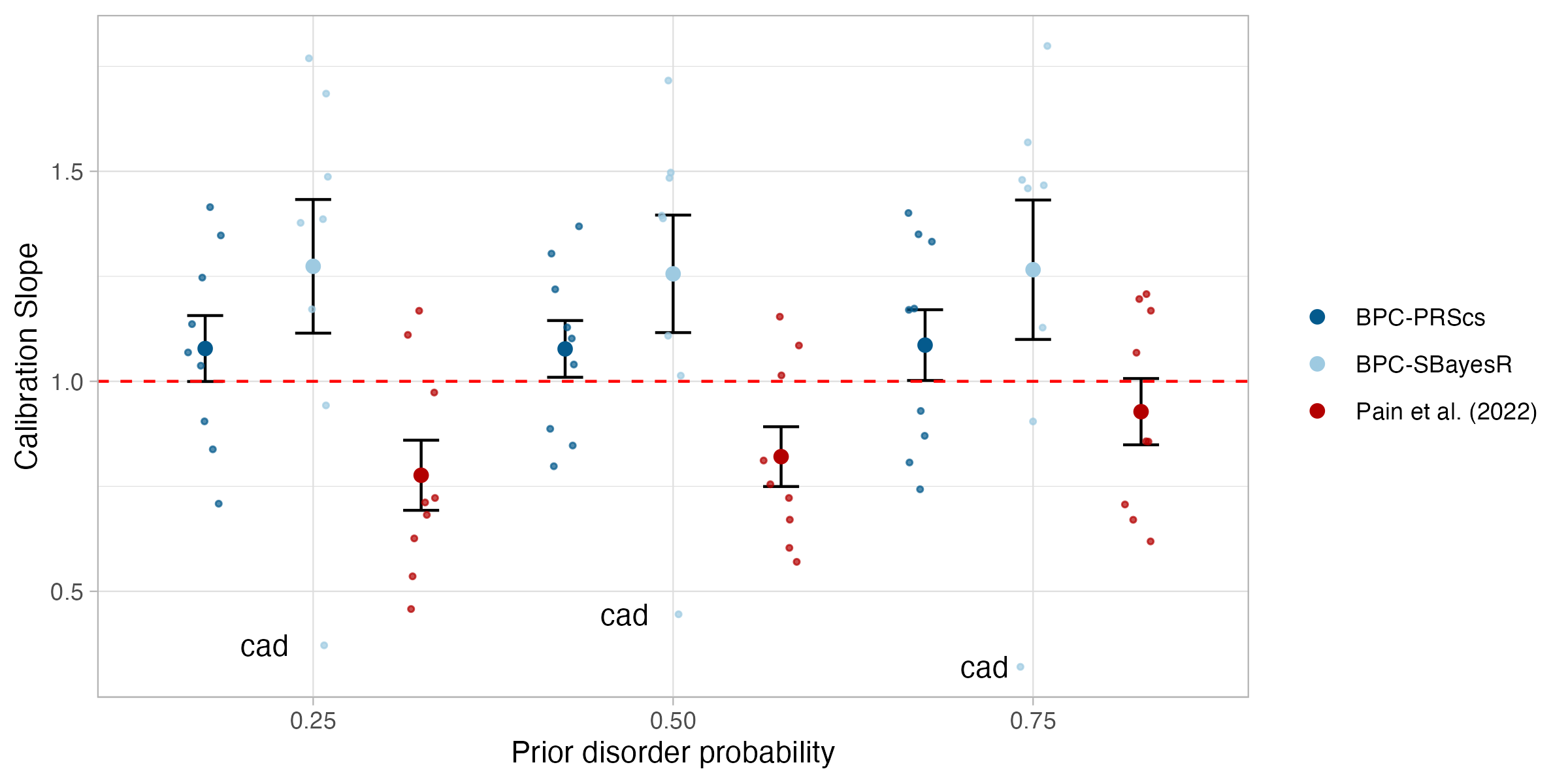

**Supplementary Figure 21 Calibration in empirical analyses of nine disorders (regression slope).**

Calibration of the BPC and the Pain et al. (2022) approach was evaluated using the slope from a regression of disorder status on the PGS for nine disorders while varying the prior disorder probability. The BPC approach was applied using two Bayesian PGS methods, PRScs (BPC-PRScs) and SBayesR (BPC-SBayesR). Error bars denote the standard error.

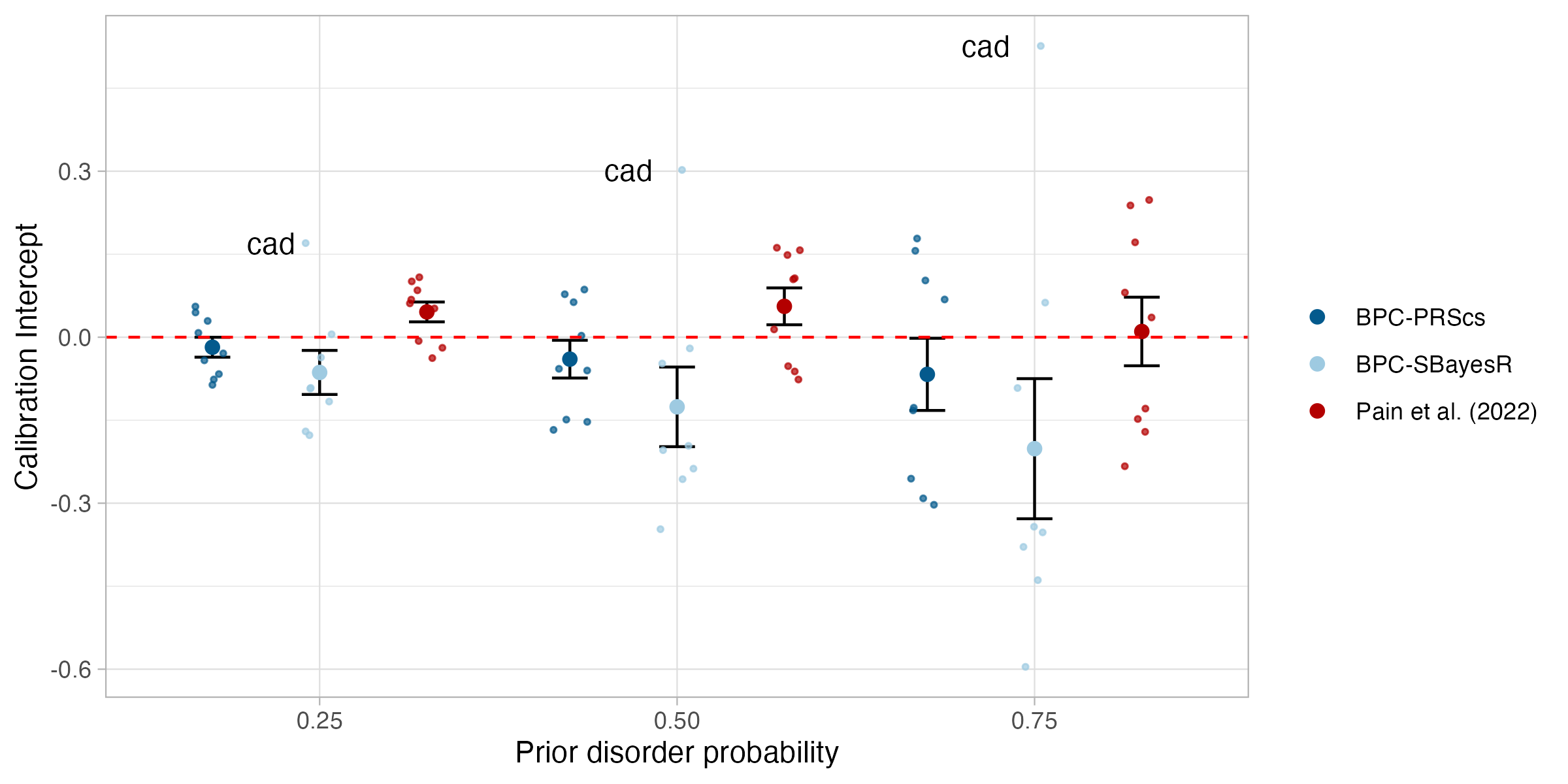

**Supplementary Figure 22 Calibration in empirical analyses of nine disorders (intercept).**

Calibration of the BPC and the Pain et al. (2022) approach was evaluated using the intercept from a regression of disorder status on the PGS for nine disorders while varying the prior disorder probability. The BPC approach was applied using two Bayesian PGS methods, PRScs (BPC-PRScs) and SBayesR (BPC-SBayesR). Error bars denote the standard error.

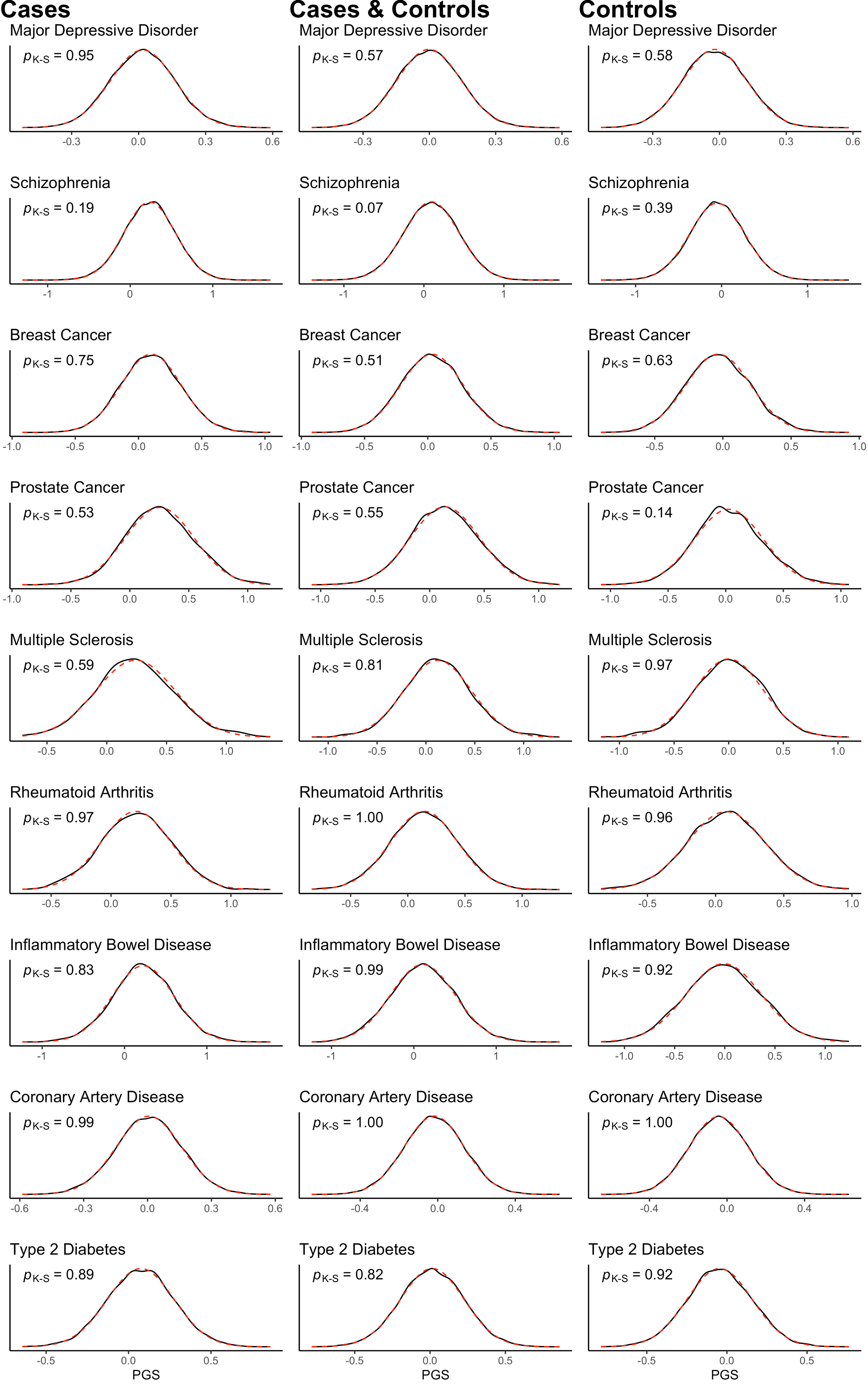

**Supplementary Figure 23 Normality of PGSs in empirical analyses (K = 0.15).**

Evaluating the normality of PGSs in only cases, only controls and in cases and controls. The BPC approach assumes normality of PGSs in cases and controls separately, but the combined distribution is shown for reference. The normality assumption is not violated for any phenotype. Normality is evaluated with the Kolmogorov-Smirnov test.

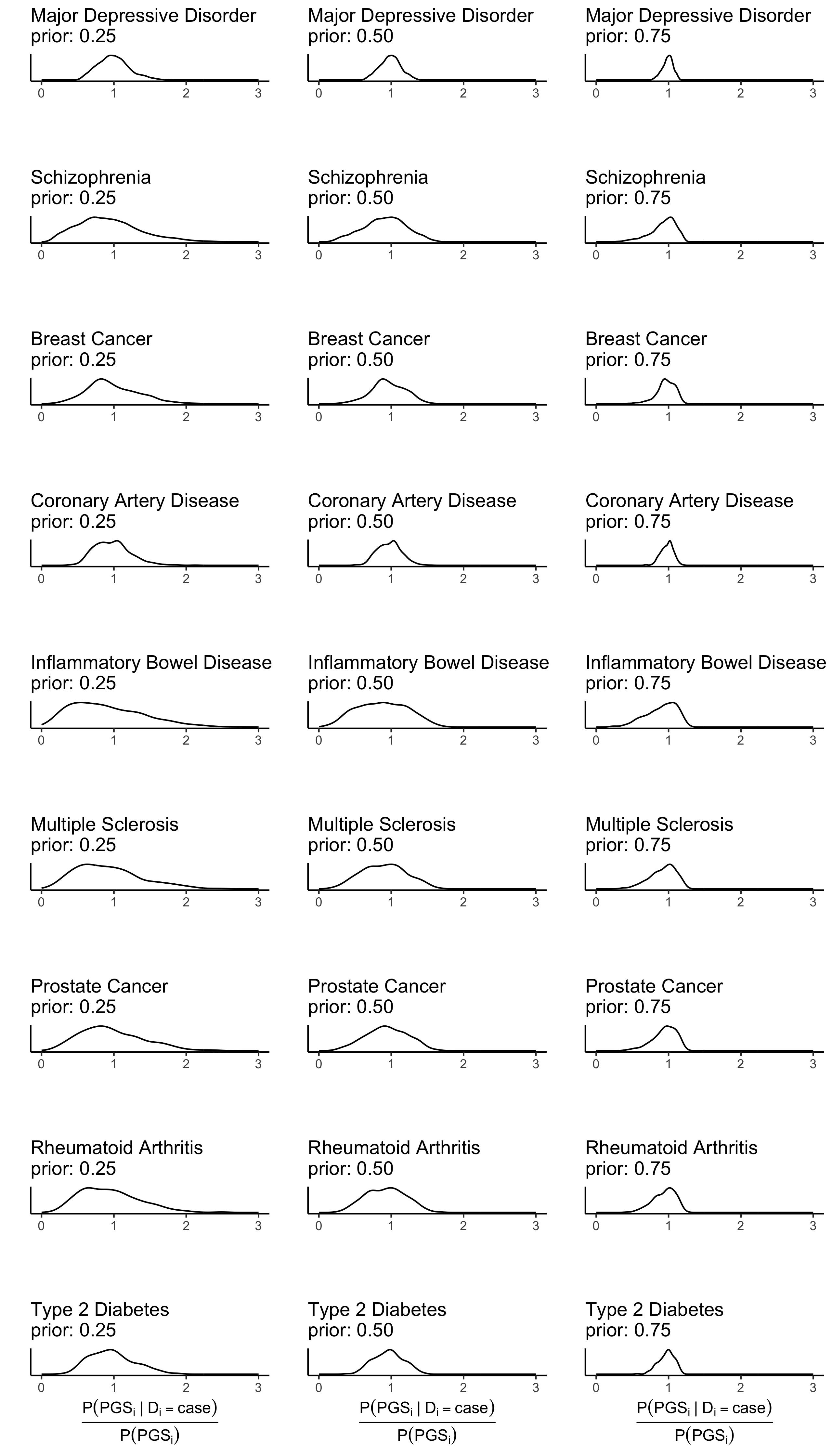

**Supplementary Figure 24 Distribution of** $\frac{\boldsymbol{P(}\boldsymbol{PGS}_{\boldsymbol{i}}\boldsymbol{|}\boldsymbol{D}_{\boldsymbol{i}}\boldsymbol{=case)}}{\boldsymbol{P(PG}\boldsymbol{S}_{\boldsymbol{i}}\boldsymbol{)}}$ **in empirical analyses.**

To evaluate the extent to which the predicted disorder probabilities are dominated by the prior, we plotted the distribution of $\frac{P({PGS}_{i}|D_{i}=case)}{P(PGS_{i})}$ for all phenotypes.

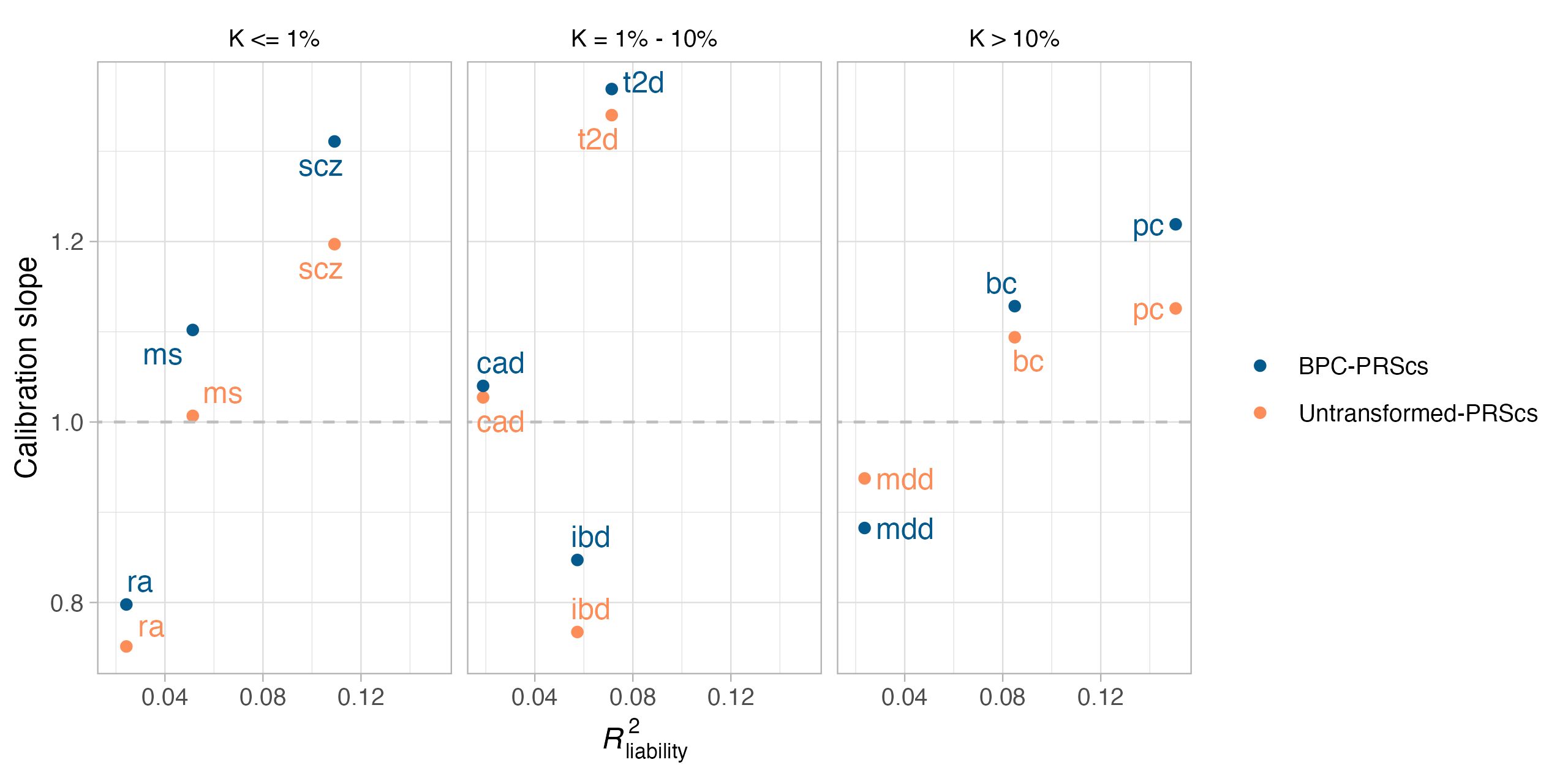

**Supplementary Figure 25 Calibration slope of untransformed Bayesian PGSs.**

Evaluating the calibration of untransformed PRScs PGSs for binary traits using the slope from a regression of standardized disorder status on the PGS in empirical analyses of nine disorders.

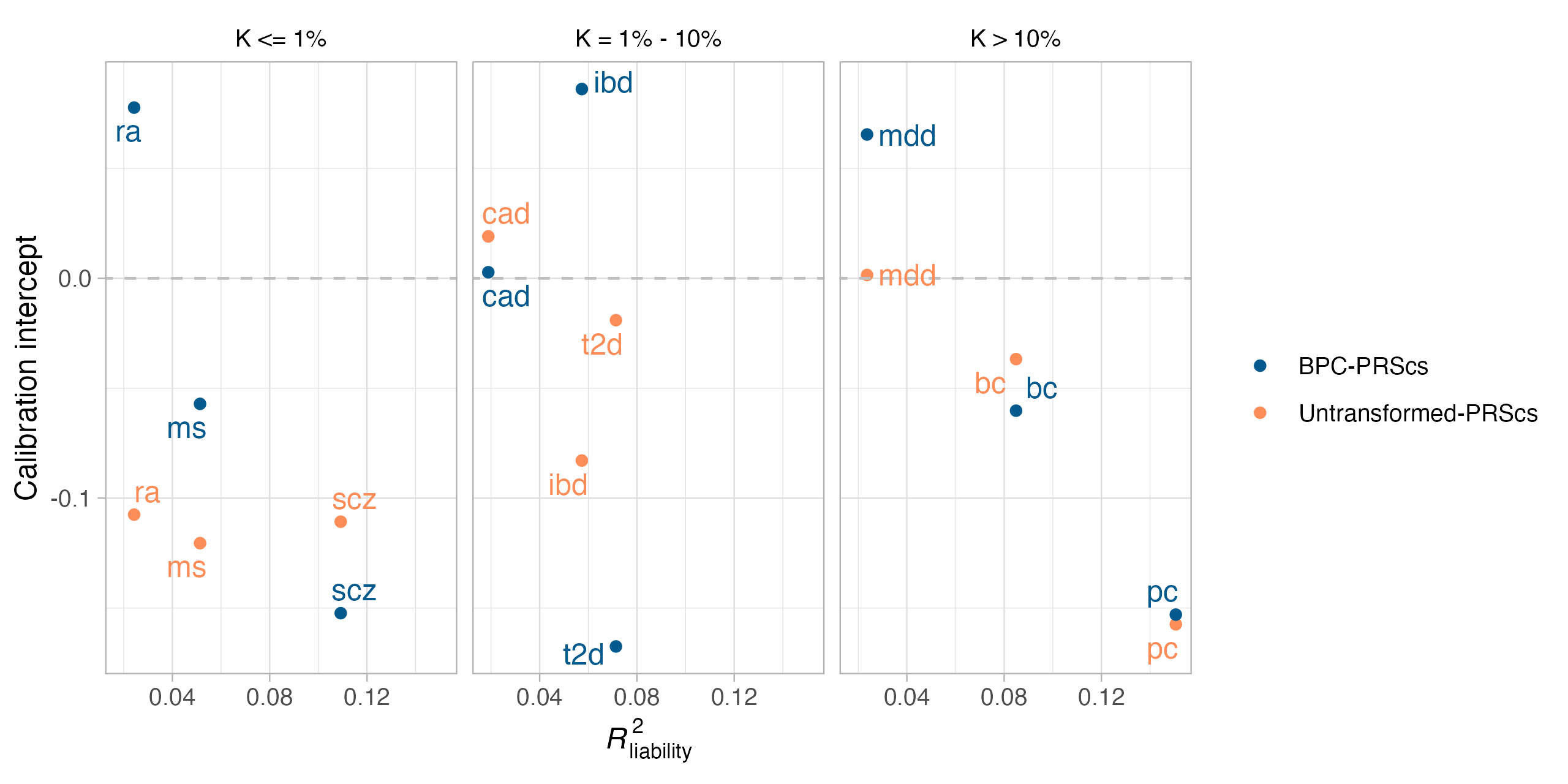

**Supplementary Figure 26 Calibration intercept of untransformed Bayesian PGSs.**

Evaluating the calibration of untransformed PRScs PGSs for binary traits using the intercept from a regression of standardized disorder status on the PGS in empirical analyses of nine disorders.

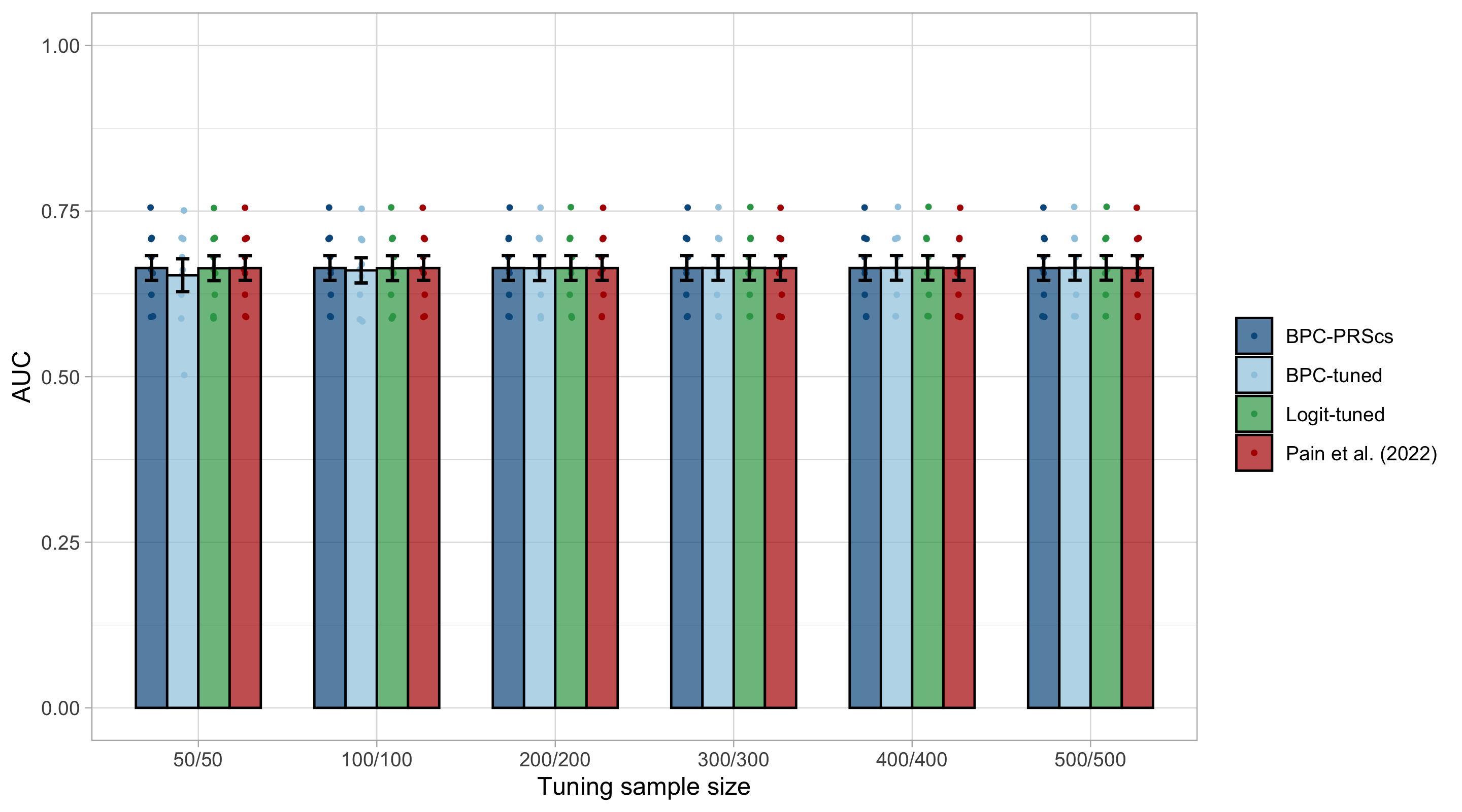

**Supplementary Figure 27** **AUC of tuning approaches in empirical analyses of nine disorders.**

AUCs of the BPC-PRScs, BPC-tuned, Logit-tuned and the Pain et al. (2022) approach are depicted for nine disorders. BPC-tuned and Logit -tuned use tuning samples that include genotype and phenotype data to estimate disorder probabilities. Tuning sample sizes are presented as (N_case_/N_control_). Error bars represent standard errors. The AUCs are nearly identical for all approaches, except for Coronary Artery Disease when the tuning sample size is 50/50. This is because the empirically estimated variances and means become unstable at low sample sizes, such that, in this case, the PGS mean is larger in controls than in cases.

### Supplementary author lists

**Major Depressive Disorder Working Group of the Psychiatric Genomics Consortium (2023)**

Mark J Adams 1 *

Fabian Streit 2 *

Swapnil Awasthi 3 *

Brett N Adey 4

Karmel W Choi 5, 6

V Kartik Chundru 7

Jonathan RI Coleman 4, 8

Jerome C Foo 2

Olga Giannakopoulou 9

Alisha S M Hall 2, 10

Jens Hjerling-Leffler 11

David M Howard 4

Christopher Hübel 4, 12, 13

Alex S F Kwong 1, 14

Bochao Danae Lin 15

Xiangrui Meng 9

Guiyan Ni 16

Oliver Pain 17

Gita A Pathak 18, 19

Eva C Schulte 20, 21, 22, 23

Jackson G Thorp 24

Alicia Walker 16

Shuyang Yao 25

Jian Zeng 16

Johan Zvrskovec 4, 8

Dag Aarsland 26

Ky'Era V Actkins 27

Mazda Adli 3, 28

Esben Agerbo 12, 29, 30

Mareike Aichholzer 31

Tracy M Air 32

Allison Aiello 33

Thomas D Als 30, 34, 35

Evelyn Andersson 36

Till F M Andlauer 37, 38

Volker Arolt 39

Helga Ask 40, 41

Sunita Badola 42

Clive Ballard 43

Karina Banasik 44

Nicholas J Bass 9

Aartjan T F Beekman 45

Sintia Belangero 46

Elisabeth B Binder 38, 47

Ottar Bjerkeset 48, 49

Gyda Bjornsdottir 50

Julia Boberg 36

Sigrid Børte 51, 52, 53

Emma Bränn 54

Alice Braun 55

Thorsten Brodersen 56

Søren Brunak 44

Mie T Bruun 57

Pichit Buspavanich 58, 59

Jonas Bybjerg-Grauholm 60, 61

Enda M Byrne 62

Archie Campbell 63, 64

Megan L. Campbell 65

Enrique Castelao 66

Jorge Cervilla 67, 68

Boris Chaumette 69

Chia-Yen Chen 70

Zhengming Chen 71, 72

Sven Cichon 73, 74, 75, 76

Lucía Colodro-Conde 24

Anne Corbett 43

Elizabeth C Corfield 40, 77

Baptiste Couvy-Duchesne 78

Nick Craddock 79, 80

Udo Dannlowski 39

Gail Davies 81

EJC de Geus 82

Ian J Deary 81

Franziska Degenhardt 76, 83

Abbas Dehghan 84, 85

J Raymond DePaulo 86

Michael Deuschle 87

Maria Didriksen 88

Khoa Manh Dinh 89

Nese Direk 90

Srdjan Djurovic 91, 92

Anna R Docherty 93, 94, 95

Katharina Domschke 96

Joseph Dowsett 88

Ole Kristian Drange 49, 97, 98, 99

Erin C Dunn 6, 100

Gudmundur Einarsson 50

Thalia C Eley 4

Samar S M Elsheikh 101

Jan Engelmann 102

Michael E Benros 60, 103, 104

Christian Erikstrup 89

Valentina Escott-Price 80

Chiara Fabbri 4, 105

Yu Fang 106

Sarah Finer 107

Josef Frank 2

Robert C Free 108

He Gao 109

Michael Gill 110

Maria Gilles 87

Fernando S Goes 86

Scott Douglas Gordon 24

Jakob Grove 30, 34, 35, 111

Daniel F Gudbjartsson 50, 112

Blanca Gutierrez 67, 68

Tim Hahn 39

Lynsey S Hall 80

Thomas F Hansen 44, 60, 113

Magnus Haraldsson 114

Catherina A Hartman 115

Alexandra Havdahl 40

Caroline Hayward 116

Stefanie Heilmann-Heimbach 76

Stefan Herms 74, 76

Ian B Hickie 117

Henrik Hjalgrim 118

Per Hoffmann 74, 76

Georg Homuth 119

Carsten Horn 120

Jouke-Jan Hottenga 82

David M Hougaard 60, 61

Iiris Hovatta 121

Qin Qin Huang 7

Floris Huider 82

Karen A Hunt 122

Marcus Ising 123

Erkki Isometsä 124

Rick Jansen 45

Yunxuan Jiang 125

Ian Jones 80

Lisa A Jones 126

Lina Jonsson 127

Robert Karlsson 25

Siegfried Kasper 128

Kenneth S Kendler 129

Ronald C Kessler 130

Stefan Kloiber 101, 123, 131, 132

James A Knowles 133

Nastassja Koen 65

Julia Kraft 55

Henry R Kranzler 134, 135

Kristi Krebs 136

Theodora Kunovac Kallak 137

Zoltán Kutalik 138, 139, 140

Elisa Lahtela 141

Margit Hørup Larsen 88

Eric J Lenze 142

Daniel F Levey 143, 144

Melissa Lewins 1

Glyn Lewis 9

Liming Li 145, 146

Kuang Lin 71

Penelope A Lind 24

Donald J MacIntyre 1, 147, 148

Dean F MacKinnon 86

Hermine HM Maes 149, 150

Wolfgang Maier 151

Victoria S Marshe 101, 152

Hamdi Mbarek 82

Peter McGuffin 4

Sarah E Medland 24

Susanne Meinert 39, 153

Susan Mikkelsen 89

Christina Mikkelsen 88, 154

Yuri Milaneschi 45

Iona Y Millwood 71, 72

Brittany L Mitchell 24

Esther Molina 67, 155

Francis M Mondimore 86

Preben Bo Mortensen 12, 29, 30

Benoit H Mulsant 101, 131

Joonas Naamanka 121

Jake M Najman 156

Matthias Nauck 157, 158

Igor Nenadić 159

Kasper R Nielsen 160

Ilja M Nolte 161

Merete Nordentoft 60, 103, 104

Markus M Nöthen 76

Mette Nyegaard 30, 162, 163, 164

Michael C O'Donovan 80

Asmundur Oddsson 50

Catherine M Olsen 165, 166

Hogni Oskarsson 167

Sisse Rye Ostrowski 88, 168

Vanessa K Ota 46

Michael J Owen 80

Richard Packer 169

Teemu Palviainen 141

Pedro M Pan 170

Carlos N Pato 171

Michele T Pato 171

Nancy L Pedersen 25

Ole Birger Pedersen 172

Roseann E Peterson 129, 173

Wouter J Peyrot 45

James B Potash 86

Martin Preisig 66

Jorge A Quiroz 174

Charles F Reynolds III 175

John P Rice 142

Giovanni A Salum 176

Robert A Schoevers 177, 178

Andrew Schork 30, 179, 180

Thomas G Schulze 2, 21, 86, 181, 182

Tabea S Send 87

Jianxin Shi 183

Engilbert Sigurdsson 114

Kritika Singh 27

Grant C B Sinnamon 184

Lea Sirignano 2

Olav B Smeland 185, 186

Daniel J Smith 187

Erik Sørensen 88

Sundararajan Srinivasan 188

Hreinn Stefansson 50

Kari Stefansson 50, 189

Dan J. Stein 190

Frederike Stein 191

André Tadic 102, 192

Henning Teismann 193

Alexander Teumer 194

Anita Thapar 80, 195

Pippa A Thomson 64

Lise Wegner Thørner 88

Apostolia Topaloudi 196

Ioanna Tzoulaki 84, 85, 197

Monica Uddin 198

André G Uitterlinden 199

Henrik Ullum 88, 200, 201

Daniel Umbricht 202

Robert J Ursano 203

Sandra Van der Auwera 204

David A van Heel 122

Albert M van Hemert 205

Abirami Veluchamy 188

Alexander Viktorin 25

Henry Völzke 194

Agaz Wani 198

G Bragi Walters 50

Robin G Walters 71, 72

Sylvia Wassertheil-Smoller 206

Myrna M Weissman 207, 208

Jürgen Wellmann 193

David C Whiteman 165

Derek Wildman 198

Gonneke Willemsen 82

Alexander T Williams 169

Bendik S Winsvold 51, 52, 209

Stephanie H Witt 2

Ying Xiong 25

Lea Zillich 2

John-Anker Zwart 51, 52, 53

23andMe Research Team 125

Estonian Biobank Research Team 136

HUNT All-In Psychiatry 210

China Kadoorie Biobank Collaborative Group 211

Genes & Health Research Team 212

Ole A Andreassen 185, 186, 213

Bernhard T Baune 214, 215, 216

Klaus Berger 193

Dorret I Boomsma 82

Anders D Børglum 30, 34, 35

Gerome Breen 4, 8

Na Cai 217, 218, 219

Hilary Coon 94

William E Copeland 220

Byron Creese 43

Lea K Davis 27

Eske M Derks 24

Enrico Domenici 221

Paul Elliott 84, 85, 197, 222

Andreas J Forstner 73, 76

Micha Gawlik 223

Joel Gelernter 19, 143, 224

Hans J Grabe 204

Steven P Hamilton 225

Kristian Hveem 226, 227, 228

Catherine John 169, 229

Jaakko Kaprio 141

Tilo Kircher 159

Marie-Odile Krebs 230

Karoline Kuchenbaecker 9, 71

Mikael Landén 25, 127

Kelli Lehto 136

Douglas F Levinson 231

Qingqin S Li 232

Klaus Lieb 102

Yi Lu 25

Susanne Lucae 123

Jurjen J Luykx 15, 233

Patrik K Magnusson 25

Nicholas G Martin 24

Hilary C Martin 7

Andrew McQuillin 9

Christel M Middeldorp 62, 234

Lili Milani 136

Ole Mors 30, 235

Daniel J Müller 101, 131, 132, 236

Bertram Müller-Myhsok 38, 237, 238

Albertine J Oldehinkel 115

Sara A Paciga 239

Colin NA Palmer 188

Peristera Paschou 196

Brenda WJH Penninx 45

Roy H Perlis 5, 6, 240

Giorgio Pistis 66

Renato Polimanti 18, 19

David J Porteous 64

Danielle Posthuma 241, 242

Ted Reichborn-Kjennerud 40

Andreas Reif 31

Frances Rice 80, 243

Roland Ricken 3

Marcella Rietschel 2

Margarita Rivera 67, 244

Christian Rück 245

Catherine Schaefer 246

Srijan Sen 106, 247

Alessandro Serretti 105

Alkistis Skalkidou 137

Jordan W Smoller 5, 248, 249

Frederike Stein 191

Murray B Stein 250, 251, 252

Patrick F Sullivan 25, 253

Martin Tesli 40

Thorgeir E Thorgeirsson 50

Henning Tiemeier 254, 255

Nicholas J Timpson 14

Rudolf Uher 256

Jens R Wendland 42

Thomas Werge 60, 179, 201, 257, 258

Naomi R Wray 16, 259 **

Stephan Ripke 3, 248 **

Cathryn M Lewis 4, 260 **

Andrew M McIntosh 1, 261 **

* Joint Lead Authors

** Joint Last Authors

1, Division of Psychiatry, University of Edinburgh, Edinburgh, UK

2, Department of Genetic Epidemiology in Psychiatry, Central Institute of Mental Health, Medical Faculty Mannheim, Heidelberg University, Mannheim, BW, DE

3, Department of Psychiatry and Psychotherapy, Charité – Universitätsmedizin Berlin, Berlin, BE, DE

4, Social, Genetic and Developmental Psychiatry Centre, King's College London, London, UK

5, Department of Psychiatry, Massachusetts General Hospital, Boston, MA, US

6, Department of Psychiatry, Harvard Medical School, Boston, MA, US

7, Human Genetics, Wellcome Sanger Institute, Hinxton, UK

8, NIHR Maudsley Biomedical Research Centre, King's College London, London, UK

9, Division of Psychiatry, University College London, London, UK

10, Department of Clinical Medicine, Aarhus University, Aarhus, DK

11, Department of Medical Biochemistry and Biophysics, Karolinska Institutet, Stockholm, SE

12, National Centre for Register-based Research, Aarhus University, Aarhus, DK

13, Department of Pediatric Neurology, Charité – Universitätsmedizin Berlin, Berlin, BE, DE

14, MRC Integrative Epidemiology Unit, University of Bristol, Bristol, UK

15, Department of Psychiatry and Neuropsychology, School for Mental Health and Neuroscience, Maastricht University Medical Centre, Maastricht, NL

16, Institute for Molecular Bioscience, University of Queensland, Brisbane, QLD, AU

17, Maurice Wohl Clinical Neuroscience Institute, Department of Basic and Clinical Neuroscience, King's College London, London, UK

18, Veterans Affairs Connecticut Healthcare System, West Haven, CT, US

19, Department of Psychiatry, Yale University School of Medicine, New Haven, CT, US

20, Department of Psychiatry, University of Munich, Munich, BY, DE

21, Institute of Psychiatric Phenomics and Genomics, University of Munich, Munich, BY, DE

22, Department of Psychiatry and Psychotherapy, University Hospital Bonn, Medical Faculty, University of Bonn, Bonn, DE

23, Institute of Human Genetics, University Hospital Bonn, Medical Faculty, University of Bonn, Bonn, DE

24, Mental Health and Neuroscience, QIMR Berghofer Medical Research Institute, Brisbane, QLD, AU

25, Department of Medical Epidemiology and Biostatistics, Karolinska Institutet, Stockholm, SE

26, Old Age Psychiatry, King's College London, London, UK

27, Department of Medicine, Division of Genetic Medicine, Vanderbilt University Medical Center, Nashville, TN, US

28, Department of Psychiatry and Psychotherapy, Fliedner Klinik Berlin, Berlin, BE, DE

29, Centre for Integrated Register-based Research, Aarhus University, Aarhus, DK

30, iPSYCH, The Lundbeck Foundation Initiative for Integrative Psychiatric Research, Aarhus, DK

31, Department of Psychiatry, Psychosomatic Medicine and Psychotherapy, Goethe University Frankfurt - University Hospital, Frankfurt am Main, DE

32, Discipline of Psychiatry, University of Adelaide, Adelaide, SA, AU

33, Department of Epidemiology, Columbia University Mailman School of Public Health, New York, NY, US

34, Department of Biomedicine and Centre for Integrative Sequencing, iSEQ, Aarhus University, Aarhus, DK

35, Center for Genomics and Personalized Medicine, Aarhus University, Aarhus, DK

36, Department of Clinical Neuroscience, Karolinska Institutet,, SE

37, Department of Neurology, Klinikum rechts der Isar, Technical University of Munich, Munich, BY, DE

38, Department of Translational Research in Psychiatry, Max Planck Institute of Psychiatry, Munich, BY, DE

39, Institute for Translational Psychiatry, University of Münster, Münster, NRW, DE

40, Department of Mental Disorders, Norwegian Institute of Public Health, Oslo, NO

41, PROMENTA Research Center, Department of Psychology, University of Oslo, Oslo, NO

42, Research and Development, Takeda Pharmaceutical Company Limited, Cambridge, MA, US

43, Faculty of Health and Life Sciences, University of Exeter, Exeter, UK

44, Novo Nordisk Center for Protein Research, Department of Health Sciences, University of Copenhagen, Copenhagen, DK

45, Department of Psychiatry, Amsterdam Public Health and Amsterdam Neuroscience, Amsterdam UMC, Vrije Universiteit Amsterdam, Amsterdam, NL

46, Morphology and Genetics, Universidade Federal de Sao Paulo, Sao Paulo, SP, BR

47, Department of Psychiatry and Behavioral Sciences, Emory University School of Medicine, Atlanta, GA, US

48, Faculty of Nursing and Health Sciences, NORD University, Levanger, NO

49, Department of Mental Health, Faculty of Medicine and Health Sciences, Norwegian University of Science and Technology (NTNU), Trondheim, TRD, NO

50, deCODE Genetics / Amgen, Reykjavik, IS

51, K. G. Jebsen Center for Genetic Epidemiology, Department of Public Health and Nursing, Faculty of Medicine and Health Sciences, Norwegian University of Science and Technology (NTNU), Trondheim, TRD, NO

52, Department of Research and Innovation, Division of Clinical Neuroscience, Oslo University Hospital, Oslo, NO

53, Institute of Clinical Medicine, Faculty of Medicine, University of Oslo, Oslo, NO

54, Institute of Environmental Medicine, Unit of Integrative Epidemiology, Karolinska Institutet, Stockholm, SE

55, Department of Psychiatry and Psychotherapy, Charité – Universitätsmedizin Berlin, Berlin, DE

56, Department of Clinical Immunology, Roskilde University/Næstved Hospital, Roskilde, DK

57, Department of Clinical Immunology, Odense University Hospital, Odense, DK

58, Department of Psychiatry, Psychotherapy and Psychosomatics, Brandenburg Medical School Theodor Fontane, Neuruppin, BB, DE

59, Department of Psychiatry and Psychotherapy, Gender Research in Medicine, Institute of Sexology and Sexual Medicine, Charité – Universitätsmedizin Berlin, Berlin, BE, DE

60, iPSYCH, The Lundbeck Foundation Initiative for Integrative Psychiatric Research, Copenhagen, DK

61, Center for Neonatal Screening, Department for Congenital Disorders, Statens Serum Institut, Copenhagen, DK

62, Child Health Research Centre, University of Queensland, Brisbane, QLD, AU

63, Centre for Medical Informatics, Usher Institute, University of Edinburgh, Edinburgh, UK

64, Centre for Genomic & Experimental Medicine, Institute for Genetics and Cancer, University of Edinburgh, Edinburgh, UK

65, Department of Psychiatry and Mental Health, University of Cape Town, Cape Town, SA

66, Department of Psychiatry, Lausanne University Hospital and University of Lausanne, Prilly, VD, CH

67, Instituto de Investigación Biosanitaria ibs.GRANADA, Granada, ES

68, Department of Psychiatry, Faculty of Medicine and Institute of Neurosciences, Biomedical Research Centre (CIBM), University of Granada, Granada, ES

69, Université de Paris Cité, INSERM U1266, Institute of Psychiatry and Neuroscience of Paris, GHU Paris Psychiatry and Neuroscience, Paris, FR

70, Translational Biology, Biogen, Cambridge, MA, US

71, Nuffield Department of Population Health, University of Oxford, Oxford, UK

72, MRC Population Health Research Unit, University of Oxford, Oxford, UK

73, Institute of Neuroscience and Medicine (INM-1), Research Center Juelich, Juelich, DE

74, Human Genomics Research Group, Department of Biomedicine, University of Basel, Basel, CH

75, Institute of Medical Genetics and Pathology, University Hospital Basel, University of Basel, Basel, CH

76, Institute of Human Genetics, University of Bonn, School of Medicine & University Hospital Bonn, Bonn, DE

77, Nic Waals Institute, Lovisenberg Diakonale Hospital, Oslo, NO

78, Centre for Advanced Imaging, University of Queensland, Saint Lucia, QLD, AU

79, Psychological Medicine, Cardiff University, Cardiff, WLS, UK

80, Centre for Neuropsychiatric Genetics and Genomics, Cardiff University, Cardiff, WLS, UK

81, The Lothian Birth Cohorts, University of Edinburgh, Edinburgh, UK

82, Department of Biological Psychology & Amsterdam Public Health Research Institute, Vrije Universiteit Amsterdam, Amsterdam, NL

83, Department of Child and Adolescent Psychiatry, Psychosomatics and Psychotherapy, University Hospital Essen, Unversity of Duisburg-Essen, Duisburg, DE

84, MRC Centre for Environment and Health, School of Public Health, Imperial College London, London, UK

85, Imperial College Dementia Research Institute, Imperial College London, London, UK

86, Department of Psychiatry and Behavioral Sciences, Johns Hopkins University School of Medicine, Baltimore, MD, US

87, Department of Psychiatry and Psychotherapy, Research Group Stress Related Disorders, Central Institute of Mental Health, Medical Faculty Mannheim, Heidelberg University, Mannheim, BW, DE

88, Department of Clinical Immunology, Copenhagen University Hospital, Rigshospitalet, Copenhagen, CPH, DK

89, Department of Clinical Immunology, Aarhus University Hospital, Aarhus, DK

90, Department of Psychiatry, Istanbul University, Istanbul, TR

91, Department of Medical Genetics, Oslo University Hospital, Oslo, OSL, NO

92, NORMENT, Department of Clinical Science, University of Bergen, Bergen, NO

93, Virginia Institute for Psychiatric & Behavioral Genetics, Virginia Commonwealth University, Richmond, VA, US

94, Psychiatry Department / Huntsman Mental Health Institute, University of Utah School of Medicine, Salt Lake City, UT, US

95, Center for Genomic Research, University of Utah School of Medicine, Salt Lake City, UT, US

96, Department of Psychiatry and Psychotherapy, Medical Center, University of Freiburg, Faculty of Medicine, University of Freiburg, Freiburg, DE

97, Division of Mental Health Care, St. Olavs Hospital, Trondheim University Hospital, Trondheim, TRD, NO

98, Department of Psychiatry, Sørlandet Hospital, Kristiansand, AG, NO

99, University of Oslo, NORMENT Centre, Institute of Clinical Medicine, Oslo, OSL, NO

100, Center for Genomic Medicine, Massachusetts General Hospital, Boston, MA, US

101, Centre for Addiction and Mental Health, Toronto, ON, CA

102, Department of Psychiatry and Psychotherapy, University Medical Center of the Johannes Gutenberg University Mainz, Mainz, DE

103, Mental Health Center Copenhagen, Mental Health Services Capital Region of Denmark, Copenhagen, DK

104, Faculty of Health Science, Department of Clinical Medicine, University of Copenhagen, Copenhagen, DK

105, Department of Biomedical and Neuromotor Sciences, University of Bologna, Bologna, IT

106, Michigan Neuroscience Institute, University of Michigan, Ann Arbor, MI, US

107, Wolfson Institute of Population Health, Queen Mary University of London, London, UK

108, School of Computing and Mathematical Sciences, University of Leicester, Leicester, UK

109, Department of Epidemiology and Biostatistics, Imperial College London, London, UK

110, Discipline of Psychiatry, School of Medicine, Trinity College Dublin, Dublin, IE

111, Bioinformatics Research Centre, Aarhus University, Aarhus, DK

112, School of Engineering, University of Iceland, Reykjavik, IS

113, Danish Headache Centre, Department of Neurology, Rigshospitalet, Glostrup, DK

114, Faculty of Medicine, Department of Psychiatry, University of Iceland, Reykjavik, IS

115, Department of Psychiatry, University of Groningen, University Medical Center Groningen, Groningen, NL

116, MRC Human Genetics Unit, Institute for Genetics and Cancer, University of Edinburgh, Edinburgh, UK

117, Brain and Mind Centre, University of Sydney, Sydney, NSW, AU

118, Department of Epidemiology Research, Statens Serum Institut, Copenhagen, DK

119, Interfaculty Institute for Genetics and Functional Genomics, Department of Functional Genomics, University Medicine Greifswald, Greifswald, MV, DE

120, Roche Pharmaceutical Research and Early Development, Pharmaceutical Sciences, Roche Innovation Center Basel, F. Hoffmann-La Roche Ltd, Basel, CH

121, SleepWell Research Program and Department of Psychology and Logopedics, University of Helsinki, Helsinki, FI

122, Blizard Institute, Barts and the London School of Medicine and Dentistry, Queen Mary University of London, London, UK

123, Max Planck Institute of Psychiatry, Munich, BY, DE

124, Department of Psychiatry, University of Helsinki, Helsinki, FI

125, 23andMe Research Team, 23andMe, Inc., Sunnyvale, CA, US

126, Department of Psychological Medicine, University of Worcester, Worcester, UK

127, Institution of Neuroscience and Physiology, University of Gothenburg, Gothenburg, SE

128, Department of Psychiatry and Psychotherapy, Medical University of Vienna, Vienna, AT

129, Department of Psychiatry, Virginia Commonwealth University, Richmond, VA, US

130, Health Care Policy, Harvard Medical School, Boston, MA, US

131, Department of Psychiatry, University of Toronto, Toronto, ON, CA

132, Department of Pharmacology & Toxicology, University of Toronto, Toronto, ON, CA

133, Department of Genetics, Rutgers University, Piscataway, NJ, US

134, Department of Psychiatry, Perelman School of Medicine, University of Pennsylvania, Philadelphia, PA, US

135, Mental Illness Research, Education and Clinical Center, Crescenz VA Medical Center, Philadelphia, PA, US

136, Estonian Genome Centre, Institute of Genomics, University of Tartu, Tartu, EE

137, Department of Women's and Children's Health, Uppsala University, Uppsala, SE

138, Department of Epidemiology and Health Systems, Center for Primary Care and Public Health, Lausanne, VD, CH

139, Swiss Institute of Bioinformatics, Lausanne, VD, CH

140, Department of Computational Biology, University of Lausanne, Lausanne, VD, CH

141, Institute for Molecular Medicine Finland - FIMM, University of Helsinki, Helsinki, FI

142, Department of Psychiatry, Washington University School of Medicine in St. Louis, St. Louis, MO, US

143, Psychiatry, Veterans Affairs Connecticut Healthcare System, West Haven, CT, US

144, Department of Psychiatry, Yale University, New Haven, CT, US

145, Department of Epidemiology and Biostatistics, School of Public Health, Peking University, Beijing, CN

146, Peking University Center for Public Health and Epidemic Preparedness & Response, Peking University, Beijing, CN

147, Mental Health, NHS 24, Glasgow, UK

148, Royal Edinburgh Hospital, NHS Lothian, Edinburgh, UK

149, Department of Human and Molecular Genetics, Virginia Commonwealth University, Richmond, VA, USA

150, Virginia Institute for Psychiatric and Behavioral Genetics, Virginia Commonwealth University, Richmond, VA, USA

151, Department of Psychiatry and Psychotherapy, University of Bonn, Bonn, DE

152, Center for Translational and Computational Neuroimmunology, Columbia University Medical Center, New York, NY, US

153, Institute for Translational Neuroscience, University of Münster, Münster, NRW, DE

154, Novo Nordisk Foundation Center for Basic Metabolic Research, Faculty of Health Science, Copenhagen University, Copenhagen, DK

155, Department of Nursing, Faculty of Health Sciences and Institute of Neurosciences, Biomedical Research Centre (CIBM), University of Granada, Granada, ES

156, School of Public Health, University of Queensland, Brisbane, QLD, AU

157, DZHK (German Centre for Cardiovascular Research), Partner Site Greifswald, Greifswald, MV, DE

158, Institute of Clinical Chemistry and Laboratory Medicine, University Medicine Greifswald, Greifswald, MV, DE

159, Department of Psychiatry, University of Marburg, Marburg, DE

160, Department of Clinical Immunology, Aalborg University Hospital, Aalborg, DK

161, Department of Epidemiology, University of Groningen, University Medical Center Groningen, Groningen, NL

162, Department of Health, Science and Technology, Aalborg University, Aalborg, DK

163, Centre for Integrative Sequencing, iSEQ, Aarhus University, Aarhus, DK

164, Department of Biomedicine-Human Genetics, Aarhus University, Aarhus, DK

165, Population Health, QIMR Berghofer Medical Research Institute, Brisbane, QLD, AU

166, The Fraser Institute, Faculty of Medicine, University of Queensland, Brisbane, QLD, AU

167, Humus, Reykjavik, IS

168, Department of Clinical Medicine, University of Copenhagen, Copenhagen, CPH, DK

169, Department of Population Health Sciences, University of Leicester, Leicester, UK

170, Department of Psychiatry, Universidade Federal de Sao Paulo, Sao Paulo, SP, BR

171, Department of Psychiatry, Rutgers University, Piscataway, NJ, US

172, Department of Clinical Immunology, Zealand University Hospital, Køge, DK

173, Department of Psychiatry and Behavioral Sciences, SUNY Downstate Health Sciences University, Brooklyn, NY, US

174, NMD Pharma, Lexington, MA, US

175, Psychiatry, University of Pittsburgh Medical Centre, Pittsburgh, PA, US

176, Psychiatry, Universidade Federal do Rio Grande do Sul, Porto Alegre, BR

177, Department of Psychiatry, University Medical Center Groningen, Groningen, NL

178, Research School of Behavioural and Cognitive Neurosciences (BCN), University of Groningen, Groningen, NL

179, Institute of Biological Psychiatry, Mental Health Center Sct. Hans, Mental Health Services Capital Region of Denmark, Copenhagen, DK

180, Neurogenomics Division, The Translational Genomics Research Institute (TGEN), Phoenix, AZ, US

181, Human Genetics Branch, NIMH Division of Intramural Research Programs, Bethesda, MD, US

182, Department of Psychiatry and Psychotherapy, University Medical Center Göttingen, Goettingen, NI, DE

183, Division of Cancer Epidemiology and Genetics, National Cancer Institute, Bethesda, MD, US

184, School of Medicine and Dentistry, James Cook University, Townsville, QLD, AU

185, Division of Mental Health and Addiction, Oslo University Hospital, Oslo, OSL, NO

186, NORMENT, Institute of Clinical Medicine, University of Oslo, Oslo, OSL, NO

187, Institute of Health and Wellbeing, University of Glasgow, Glasgow, UK

188, Division of Population Health and Genomics, Ninewells Hospital and School of Medicine, University of Dundee, Dundee, UK

189, Faculty of Medicine, University of Iceland, Reykjavik, IS

190, SAMRC Unit on Risk & Resilience in Mental Disorders, Department of Psychiatry and Mental Health, University of Cape Town, Cape Town, SA

191, Department of Psychiatry and Psychotherapy, University of Marburg, Marburg, HE, DE

192, Department of Psychiatry, Psychotherapy and Psychosomatics, Dr. Fontheim Mentale Gesundheit, Liebenburg, DE

193, Institute of Epidemiology and Social Medicine, University of Münster, Münster, NRW, DE

194, Institute for Community Medicine, University Medicine Greifswald, Greifswald, MV, DE

195, Wolfson Centre for Young People's Mental Health, Division of Psychological Medicine and Clinical Neurosciences, Cardiff University, Cardiff, WLS, UK

196, Department of Biological Sciences, Purdue University, West Lafayette, IN, US

197, Imperial College BHF Centre for Research Excellence, Imperial College London, London, UK

198, Genomics Program, University of South Florida College of Public Health, Tampa, FL, US

199, Department of Internal Medicine, Erasmus University Medical Center Rotterdam, Rotterdam, NL

200, Management Section, Statens Serum Institut, Copenhagen, DK

201, Department of Clinical Medicine, University of Copenhagen, Copenhagen, DK

202, Xperimed LLC, Basel, CH

203, Psychiatry, USUHS, Bethesda, US

204, Department of Psychiatry and Psychotherapy, University Medicine Greifswald, Greifswald, MV, DE

205, Department of Psychiatry, Leiden University Medical Center, Leiden, NL

206, Department of Epidemiology and Population Health, Albert Einstein College of Medicine, Bronx, NY, US

207, Department of Psychiatry, Columbia University College of Physicians and Surgeons, New York, NY, US

208, Division of Epidemiology, New York State Psychiatric Institute, New York, NY, US

209, Department of Neurology, Oslo University Hospital, Oslo, NO

210, HUNT All-In Psychiatry

211, China Kadoorie Biobank Collaborative Group

212, Genes & Health Research Team

213, KG Jebsen Centre for Neurodevelopmental Research, University of Oslo, Oslo, OSL, NO

214, Department of Psychiatry, University of Melbourne, Melbourne, VIC, AU

215, Florey Institute of Neuroscience and Mental Health, University of Melbourne, Melbourne, VIC, AU

216, Department of Psychiatry, University of Münster, Münster, NRW, DE

217, Computational Health Centre, Helmholtz Zentrum München, Neuherberg, DE

218, School of Medicine, Technical University of Munich, Munich, BY, DE

219, Helmholtz Pioneer Campus, Helmholtz Zentrum München, Neuherberg, DE

220, Department of Psychiatry, University of Vermont, Burlington, VT, US

221, Department of Cellular, Computational and Integrative Biology, Università degli Studi di Trento, Trento, IT

222, Imperial College Biomedical Research Centre, Imperial College London, London, UK

223, Department of Psychiatry, Psychosomatics and Psychotherapy, Julius-Maximilians-Universität Würzburg, Würzburg, DE

224, Department of Genetics, Department of Neuroscience, Yale University School of Medicine, New Haven, CT, US

225, Psychiatry, Kaiser Permanente Northern California, San Francisco, CA, US

226, K. G. Jebsen Center for Genetic Epidemiology, Department of Public Health and Nursing, Faculty of Medicine and Health Sciences, Norwegian University of Science and Technology (NTNU), Trondheim, NO

227, HUNT Research Center, Department of Public Health and Nursing, Faculty of Medicine and Health Sciences, Norwegian University of Science and Technology (NTNU), Trondheim, NO

228, Department of Research, Innovation and Education, St. Olavs Hospital, Trondheim University Hospital, Trondheim, NO

229, NIHR Leicester Biomedical Research Centre, Glenfield Hospital, Leicester, UK

230, Pathophysiology of Psychiatric Diseases, INSERM, Univ Paris Cité, GHU Paris, Paris, FR

231, Department of Psychiatry & Behavioral Sciences, Stanford University, Stanford, CA, US

232, Neuroscience Therapeutic Area, Janssen Research and Development, LLC, Titusville, NJ, US

233, Second Opinion Outpatient Clinic, GGNet Mental Health, Warnsveld, NL

234, Child and Youth Mental Health Service, Children's Health Queensland Hospital and Health Service, Brisbane, QLD, AU

235, Psychosis Research Unit, Aarhus University Hospital-Psychiatry, Aarhus, DK

236, Department of Psychiatry, Psychosomatics and Psychotherapy, University Hospital of Würzburg, Würzburg, DE

237, Munich Cluster for Systems Neurology (SyNergy), Munich, BY, DE

238, University of Liverpool, Liverpool, UK

239, Human Genetics and Computational Biomedicine, Pfizer Global Research and Development, Groton, CT, US

240, Centre for Quantitative Health, Massachusetts General Hospital, Boston, MA, US

241, Child and Adolescent Psychiatry, Amsterdam UMC, Vrije Universiteit Amsterdam, Amsterdam, NL

242, Complex Trait Genetics, Vrije Universiteit Amsterdam, Amsterdam, NL

243, Wolfson Centre for Young People's Mental Health, Division of Psychological Medicine and Clinical Neurosciences, Cardiff University, Cardiff, UK

244, Department of Biochemistry and Molecular Biology II, Faculty of Pharmacy and Institute of Neurosciences, Biomedical Research Centre (CIBM), University of Granada, Granada, ES

245, Department of Clinical Neuroscience, Karolinska Institutet, Stockholm, SE

246, Division of Research, Kaiser Permanente Northern California, Oakland, CA, US

247, Department of Psychiatry, University of Michigan, Ann Arbor, MI, US

248, Stanley Center for Psychiatric Research, Broad Institute of MIT and Harvard, Cambridge, MA, US

249, Psychiatric and Neurodevelopmental Genetics Unit, Massachusetts General Hospital, Boston, MA, US

250, Psychiatry, UCSD School of Medicine, La Jolla, CA, US

251, Public Health, UCSD School of Public Health, La Jolla, CA, US

252, Psychiatry, Veterans Affairs San Diego Healthcare System, San Diego, CA, US

253, Departments of Genetics and Psychiatry, University of North Carolina at Chapel Hill, Chapel Hill, NC, US

254, Child and Adolescent Psychiatry, Erasmus University Medical Center Rotterdam, Rotterdam, NL

255, Social and Behavioral Science, Harvard T.H. Chan School of Public Health, Boston, MA, US

256, Psychiatry, Dalhousie University, Halifax, NS, CA

257, Institute of Biological Psychiatry, Mental Health Center Sct. Hans, Copenhagen University Hospital, Mental Health Services, Copenhagen, DK

258, GLOBE Institute, Lundbeck Foundation Centre for Geogenetics, University of Copenhagen, Copenhagen, DK

259, Queensland Brain Institute, University of Queensland, Brisbane, QLD, AU

260, Department of Medical & Molecular Genetics, King's College London, London, UK

261, Institute for Genomics and Cancer, University of Edinburgh, Edinburgh, UK

**Schizophrenia Working Group of the Psychiatric Genomics Consortium**

Vassily Trubetskoy 1, Antonio F Pardiñas 2, Georgia Panagiotaropoulou 1, Swapnil Awasthi 1, Tim B Bigdeli 3, 267, 397, Charlotte A Dennison 2, Lynsey S Hall 2, Max Lam 4, 268, 398, Oleksandr Frei 5, 269, 399, Alexander L Richards 2, Jakob Grove 6, 270, 400, Zhiqiang Li 7, 271, Mark Adams 8, Ingrid Agartz 5, 272, 401, Elizabeth G Atkinson 9, 273, Esben Agerbo 10, 6, Mariam Al Eissa 11, Margot Albus 12, Madeline Alexander 13, Behrooz Z Alizadeha 14, 274, Köksal Alptekin 15, 275, Thomas D Als 6, 270, 400, Farooq Amin 16, Volker Arolt 17, Manuel Arrojo 18, Lavinia Athanasiu 5, 269, Maria Helena Azevedo 19, Silviu A Bacanu 20, Nicholas J Bass 11, Martin Begemann 21, Richard A Belliveau 22, Judit Bene 23, Beben Benyamin 24, 276, 402, Sarah E Bergen 25, Giuseppe Blasi 26, Julio Bobes 27, 277, 403, Stefano Bonassi 28, Alice Braun 1, Rodrigo Affonseca Bressan 29, 278, Evelyn J Bromet 30, Richard Bruggeman 14, 279, Peter F Buckley 31, Randy L Buckner 32, Jonas Bybjerg-Grauholm 33, 280, Wiepke Cahn 34, 281, Murray J Cairns 35, 282, 404, Monica E Calkins 36, Vaughan J Carr 37, 283, 405, David Castle 38, 284, Stanley V Catts 39, 285, Kimberley D Chambert 40, Raymond CK Chan 41, 286, Boris Chaumette  42, 287, Wei Cheng 43, Eric FC Cheung 44, Siow Ann Chong 45, 288, David Cohen 46, 289, 406, Angèle Consoli 46, 290, Quirino Cordeiro 47, Javier Costas 48, Charles Curtis 49, 291, Michael Davidson 50, Kenneth L Davis 51, Lieuwe de Haan 52, 292, Franziska Degenhardt 53, Lynn E DeLisi 54, 293, Ditte Demontis 6, 270, 400, Faith Dickerson 55, Dimitris Dikeos 56, Timothy Dinan 57, 294, Srdjan Djurovic 58, 295, Jubao Duan 59, 296, Giuseppe Ducci 60, Johan G Eriksson 61, 297, 407, Lourdes Fañanás 62, 298, Stephen V Faraone 63, Alessia Fiorentino 11, Andreas Forstner 53, 299, Josef Frank 64, Nelson B Freimer 65, 300, Menachem Fromer 66, Alessandra Frustaci 67, Ary Gadelha 29, 278, Giulio Genovese 22, Elliot S Gershon 68, Marianna Giannitelli 69, 289, Ina Giegling 70, Paola Giusti-Rodríguez 71, Stephanie Godard 72, Jacqueline I Goldstein 73, Javier González Peñas 74, 301, Ana González-Pinto 75, 301, Srihari Gopal 76, Jacob Gratten 77, 302, Michael F Green 78, 303, Tiffany A Greenwood 79, Olivier Guillin 80, 304, 408, Sinan Gülöksüz 81, 305, Raquel E Gur 36, Ruben C Gur 36, Blanca Gutiérrez 82, Eric Hahn 83, Hakon Hakonarson 84, Vahram Haroutunian 51, 306, 409, Annette M Hartmann 70, Carol Harvey 38, 307, Caroline Hayward 85, Frans A Henskens 86, Stefan Herms 87, Per Hoffmann 87, Daniel P Howrigan 73, 308, Masashi Ikeda 88, Conrad Iyegbe 89, Inge Joa 90, Antonio Julià 91, Anna K Kähler 25, Tony Kam-Thong 92, Yoichiro Kamatani 93, 309, Sena Karachanak-Yankova 94, 310, Oussama Kebir 42, Matthew C Keller 95, Brian J Kelly 86, Andrey Khrunin 96, Sung-Wan Kim 97, Janis Klovins 98, Nikolay Kondratiev 99, Bettina Konte 70, Julia Kraft 1, 311, Michiaki Kubo 100, Vaidutis Kučinskas 101, Zita Ausrele Kučinskiene 101, Agung Kusumawardhani 102, Hana Kuzelova-Ptackova 103, Stefano Landi 104, Laura C Lazzeroni 105, 312, Phil H Lee 106, 22, Sophie E Legge 2, Douglas S Lehrer 107, Rebecca Lencer 17, Bernard Lerer 108, Miaoxin Li 109, Jeffrey Lieberman 110, Gregory A Light 111, 79, Svetlana Limborska 96, Chih-Min Liu 112, 212, Jouko Lönnqvist 113, 313, Carmel M Loughland 114, Jan Lubinski 115, Jurjen J Luykx 116, 314, 410, Amy Lynham 2, Milan Macek Jr 117, Andrew Mackinnon 118, 315, Patrik KE Magnusson 25, Brion S Maher 119, Wolfgang Maier 120, Dolores Malaspina 51, 316, Jacques Mallet 121, Stephen R Marder 122, Sara Marsal 91, Alicia R Martin 73, 317, 411, Lourdes Martorell 123, Manuel Mattheisen 124, 318, 412, Robert W McCarley 125, 319, Colm McDonald 126, John J McGrath 10, 320, 413, Helena Medeiros 127, 321, Sandra Meier 128, 322, Bela Melegh 129, Ingrid Melle 5, 269, Raquelle I Mesholam-Gately 130, 323, Andres Metspalu 131, Patricia T Michie 132, Lili Milani 133, Vihra Milanova 134, Marina Mitjans 21, Espen Molden 135, 324, Esther Molina 136, María Dolores Molto 137, 301, 414, Valeria Mondelli 138, 245, Carmen Moreno 74, 301, Christopher P Morley 139, Gerard Muntané 123, 325, Kieran C Murphy 140, Inez Myin-Germeys 141, Igor Nenadić 142, 326, Gerald Nestadt 143, Liene Nikitina-Zake 98, Cristiano Noto 29, 278, Keith H Nuechterlein 78, Niamh Louise O'Brien 11, F Anthony O'Neill 144, Sang-Yun Oh 145, 327, Ann Olincy 146, Vanessa Kiyomi Ota 147, 278, Christos Pantelis 148, 328, 307, George N Papadimitriou 56, Mara Parellada 74, 301, Tiina Paunio 149, 329, Renata Pellegrino 84, Sathish Periyasamy 150, 330, Diana O Perkins 151, Bruno Pfuhlmann 152, Olli Pietiläinen 153, 331, 415, Jonathan Pimm 154, David Porteous 155, John Powell 156, Diego Quattrone 49, 291, 416, Digby Quested 157, 332, Allen D Radant 158, 333, Antonio Rampino 26, Mark H Rapaport 159, Anna Rautanen 92, Abraham Reichenberg 51, Cheryl Roe 160, Joshua L Roffman 161, Julian Roth 162, Matthias Rothermundt 17, Bart PF Rutten 163, Safaa Saker-Delye 164, Veikko Salomaa 165, Julio Sanjuan 166, 301, 414, Marcos Leite Santoro 147, 278, Adam Savitz 76, Ulrich Schall 167, 334, Rodney J Scott 35, 335, 417, Larry J Seidman 168, 336, Sally Isabel Sharp 11, Jianxin Shi 169, Larry J Siever 51, 337, Kang Sim 170, 338, 418, Nora Skarabis 1, Petr Slominsky 96, Hon-Cheong So 171, 339, Janet L Sobell 127, Erik Söderman 172, Helen J Stain 173, 340, Nils Eiel Steen 174, 269, Agnes A. Steixner-Kumar 21, Elisabeth Stögmann 175, William S Stone 176, 341, Richard E Straub 177, Fabian Streit 178, Eric Strengman 179, T Scott Stroup 110, Mythily Subramaniam 45, 342, Catherine A Sugar 180, 78, Jaana Suvisaari 165, Dragan M Svrakic 181, Neal R Swerdlow 79, Jin P Szatkiewicz 71, Thi Minh Tam Ta 182, 343, Atsushi Takahashi 163, 183, Chikashi Terao 183, Florence Thibaut 184, 344, Draga Toncheva 94, 345, Paul A Tooney 35, 282, 404, Silvia Torretta 26, Sarah Tosato 185, Gian Battista Tura 186, Bruce I Turetsky 36, Alp Üçok 187, Arne Vaaler 188, 346, Therese van Amelsvoort 163, 245, Ruud van Winkel 189, 163, Juha Veijola 190, 347, John Waddington 191, Henrik Walter 192, Anna Waterreus 193, 348, Bradley T Webb 20, Mark Weiser 194, Nigel M Williams 2, Stephanie H Witt 64, Brandon K Wormley 20, Jing Qin Wu 195, Zhida Xu 196, Robert Yolken 197, Clement C Zai 198, 349, Wei Zhou 199, Feng Zhu 200, 350, Fritz Zimprich 201, Eşref Cem Atbaşoğlu 202, 316, Muhammad Ayub 203, Alessandro Bertolino 26, Donald W Black 204, Nicholas J Bray 2, Gerome Breen 49, Nancy G Buccola 205, William F Byerley 206, Wei J Chen 207, 351, C Robert Cloninger 181, Benedicto Crespo-Facorro 208, 352, Gary Donohoe 209, Robert Freedman 146, Cherrie Galletly 210, 353, 419, Massimo Gennarelli 211, 354, David M Hougaard 33, 280, Hai-Gwo Hwu 212, 355, Assen V Jablensky 213, Steven A McCarroll 22, Jennifer L Moran 40, 356, Ole Mors 6, 357, Preben B Mortensen 10, 6, Bertram Müller-Myhsok 214, 358, 420, Amanda L Neil 215, Merete Nordentoft 33, 359, Michele T Pato 216, 360, Tracey L Petryshen 106, Ann E Pulver 143, Thomas G Schulze 217, 361, 421, Jeremy M Silverman 51, 337, Jordan W Smoller 106, 362, Eli A Stahl 66, 363, 422, Debby W Tsuang 218, 364, Elisabet Vilella 123, Shi-Heng Wang 219, Shuhua Xu 220, 365, 423, Rolf Adolfsson 221, Celso Arango 74, 301, Bernhard T Baune 17, 366, 328, Sintia Iole Belangero 147, 278, Anders D Børglum 6, 270, 400, David Braff 79, 367, Elvira Bramon 222, Joseph D Buxbaum 51, Dominique Campion 80, 304, Jorge A Cervilla 223, Sven Cichon 224, 368, 424, David A Collier 225, Aiden Corvin 226, Marta Di Forti 49, 245, 416, Enrico Domenici 227, Hannelore Ehrenreich 21, Valentina Escott-Price 228, 369, Tõnu Esko 229, 370, Ayman H Fanous 230, 371, 425, Anna Gareeva 231, 372, Micha Gawlik 232, Pablo V Gejman 59, 296, Michael Gill 226, Stephen J Glatt 233, Vera Golimbet 99, Kyung Sue Hong 234, Christina M Hultman 25, Steven E Hyman 40, 373, Nakao Iwata 88, Erik G Jönsson 172, 374, René S Kahn 235, 34, James L Kennedy 198, 349, Elza Khusnutdinova 236, 375, George Kirov 2, James A Knowles 237, 376, Marie-Odile Krebs 42, Claudine Laurent-Levinson 46, 377, Jimmy Lee 238, 378, Todd Lencz 239, 379, 426, Douglas F Levinson 240, Qingqin S Li 76, Jianjun Liu 241, 380, Anil K Malhotra 239, 379, 426, Dheeraj Malhotra 242, Andrew McIntosh 8, Andrew McQuillin 154, Paulo R Menezes 243, Vera A Morgan 244, 381, Derek W Morris 126, Bryan J Mowry 150, 330, Robin M Murray 245, 382, Vishwajit Nimgaonkar 246, Markus M Nöthen 53, Roel A Ophoff 247, 65, 427, Sara A Paciga 248, Aarno Palotie 249, 383, 428, Carlos N Pato 216, 360, Shengying Qin 199, 384, Marcella Rietschel 64, Brien P Riley 20, Margarita Rivera 250, 385, Dan Rujescu 70, Meram C Saka 251, Alan R Sanders 59, 386, Sibylle G Schwab 252, 387, Alessandro Serretti 253, Pak C Sham 254, 388, 429, Yongyong Shi 255, 389, David St Clair 256, Ming T Tsuang 257, 390, Jim van Os 258, 391, Marquis P Vawter 259, Daniel R Weinberger 177, Thomas Werge 260, 392, 430, Dieter B Wildenauer 261, Xin Yu 262, 393, Weihua Yue 262, 393, 431, Peter A Holmans 2, Panos Roussos 263, 394, Evangelos Vassos 49, 245, 432, Danielle Posthuma 264, Ole A Andreassen 5, 269, Kenneth S Kendler 20, Michael J Owen 2, Naomi R Wray 265, 150, Mark J Daly 73, 395, 433, Hailiang Huang 73, 317, 411, Benjamin M Neale 73, 317, Patrick F Sullivan 266, 71, 434, Stephan Ripke 1, 396, 435, James TR Walters 2, Michael C O'Donovan 2

1: Dept. of Psychiatry and Psychotherapy, Charité - Universitätsmedizin, Berlin 10117, Germany, 2: MRC Centre for Neuropsychiatric Genetics and Genomics, Division of Psychiatry and Clinical Neurosciences, Cardiff University, Hadyn Ellis Building, Maindy Road, Cardiff, CF24 4HQ, 3: Department of Psychiatry and the Behavioral Sciences, State University of New York, Downstate Medical Center, 450 Clarkson Ave, Brooklyn, NY, 11203, 4: Stanley Center for Psychiatric Research, Broad Institute of MIT and Harvard, Cambridge, MA 02142, 5: NORMENT Centre, Division of Mental Health and Addiction, University of Oslo, 0424 Oslo, Norway, 6: The Lundbeck Foundation Initiative for Integrative Psychiatric Research (iPSYCH), Aarhus, Denmark, 7: Affiliated Hospital of Qingdao University & Biomedical Sciences Institute of Qingdao University, Qingdao University, 8: Division of Psychiatry, Centre for Clinical Brain Sciences, University of Edinburgh, Royal Edinburgh Hospital, Edinburgh, EH10 5HF, UK, 9: Analytic and Translational Genetics Unit, Massachusetts General Hospital, Boston, MA, 10: National Centre for Register-based Research, Aarhus University, DK-8210 Aarhus, Denmark, 11: Molecular Psychiatry Laboratory, Division of Psychiatry, University College London, London, WC1E 6BT, 12: Comedicum Lindwurmhof, Lindwurmstr. 88, 80337, Munich, Germany, 13: Center for Depression, Anxiety and Stress Research, McLean Hospital, Belmont, MA, USA, 14: University Medical Center Groningen, University Center for Psychiatry, Rob Giel Research Center, Unviersity of Groningen, Groningen, The Netherlands, 15: Department of Psychiatry, Dokuz Eylül University School of Medicine, Izmir, Turkey, 16: Department of Psychiatry and Behavioral Sciences, Emory University, Atlanta, Georgia 30322, USA, 17: Department of Psychiatry, University of Münster, Münster, Germany, 18: Servizo de Psiquiatría, Complexo Hospitalario Universitario de Santiago de Compostela, Servizo Galego de Saúde (SERGAS), Santiago de Compostela, Galicia, Spain, 19: Institute of Medical Psychology, Faculty of Medicine, University of Coimbra, Coimbra, PT, Portugal., 20: Virginia Institute for Psychiatric and Behavioral Genetics, Department of Psychiatry, Virginia Commonwealth University, Richmond, Virginia 23298, USA, 21: Clinical Neuroscience, Max Planck Institute of Experimental Medicine, Göttingen 37075, Germany, 22: Stanley Center for Psychiatric Research, Broad Institute of MIT and Harvard, Cambridge, Massachusetts 02142, USA, 23: Department of Medical Genetics, Medical School, University of Pécs, Pécs, Hungary, 24: Australian Centre for Precision Health, University of South Australia Cancer Research Institute, University of South Australia, Adelaide, Australia, 25: Department of Medical Epidemiology and Biostatistics, Karolinska Institutet, Stockholm SE-17177, Sweden, 26: Department of Basic Medical Science, Neuroscience and Sense Organs, University of Bari 'Aldo Moro', Bari, Italy, 27: Área de Psiquiatría-Universidad de Oviedo, Hospital Universitario Central de Asturias (HUCA), Asturias, Spain, 28: Unit of Clinical and Molecular Epidemiology, IRCCS San Raffaele Pisana, and San Raffaele University, Rome, Italy, 29: Department of Psychiatry, Universidade Federal de Sao Paulo, Rua Major Maragliano, 241- CEP 04017-030, Sao Paulo, SP, Brazil, 30: Department of Psychiatry and Behavioural Health, Stony Brook University, HSC, Level T-10, Stony Brook, NY, 31: School of Medicine, Virginia Commonwealth University,, Richmond VA, USA, 32: Department of Psychology, Harvard University, Cambridge, Massachusetts 02138, USA, 33: The Lundbeck Foundation Initiative for Integrative Psychiatric Research (iPSYCH), Denmark, 34: University Medical Center Utrecht, Department of Psychiatry, Rudolf Magnus Institute of Neuroscience, 3584 Utrecht, The Netherlands, 35: School of Biomedical Sciences and Pharmacy, University of Newcastle, Callaghan NSW 2308, Australia, 36: Department of Psychiatry, University of Pennsylvania, 3400 Spruce Street, Philadelphia, PA 19104, 37: School of Psychiatry, University of New South Wales, Sydney, Australia, 38: Department of Psychiatry, The University of Melbourne, Parkville, Victoria 3010, Australia, 39: Brain and Mind Centre, The University of Sydney, Sydney, NSW, Australia, 40: Stanley Center for Psychitric Research, Broad Institute of MIT and Harvard, Cambridge, MA 02142, 41: Institute of Psychology, Chinese Academy of Science, Beijing 100101, China, 42: INSERM U1266, Institute of Psychiatry and Neuroscience of Paris, Université de Paris, GHU Paris Psychiatrie & Neurosciences, 108 rue de la Santé, 75014 Paris, France, 43: Department of Computer Science, University of North Carolina, Chapel Hill, North Carolina 27514, USA, 44: Castle Peak Hospital, Hong Kong, China, 45: Research Division, Institute of Mental Health, Singapore, 46: Faculté de Médecine Sorbonne Université, Groupe de Recherche Clinique n°15 - Troubles Psychiatriques et Développement (PSYDEV), Department of Child and Adolescent Psychiatry, Hôpital Universitaire de la Pitié-Salpêtrière, 47-83 Boulevard de l’Hôpital, 75651 Paris Cedex 13, France., 47: Department of Psychiatry, Irmandade da Santa Casa de Misericórdia de São Paulo, Rua Dona Veridiana, 55 - São Paulo - SP - CEP 01238-010, São Paulo, SP, Brazil, 48: Instituto de Investigación Sanitaria (IDIS) de Santiago de Compostela, Complexo Hospitalario Universitario de Santiago de Compostela (CHUS), Servizo Galego de Saúde (SERGAS), Santiago de Compostela, Galicia, Spain., 49: Social, Genetic and Developmental Psychiatry Centre, Institute of Psychiatry, Psychology and Neuroscience, King’s College London, London SE5 8AF, UK, 50: University of Nicosia Medical School, Nicosia, Cyprus, 51: Department of Psychiatry, Icahn School of Medicine at Mount Sinai, New York, New York 10029, USA, 52: Department of Psychiatry, Academic Medical Centre, University of Amsterdam, Amsterdam, The Netherlands, 53: Institute of Human Genetics, University of Bonn, D-53127 Bonn, Germany, 54: Cambridge Health Alliance, Cambridge, MA 02139, USA, 55: Sheppard Pratt Health System, Baltimore, MD, USA, 56: First Department of Psychiatry, Medical School, National and Kapodistrian University of Athens, Eginition Hospital, Athens 11528, Greece, 57: Department of Psychiatry and Neurobehavioural Sciences, University College Cork, Cork, Ireland, 58: NORMENT Centre, Department of Clinical Science, University of Bergen, Bergen, Norway, 59: Center for Psychiatric Genetics, NorthShore University HealthSystem, Evanston, IL 60201, USA., 60: Department of Mental Health, ASL Rome 1, 00135 Rome, Italy, 61: Department of General Practice and Primary Health Care, University of Helsinki and Helsinki University Hospital, Helsinki, Finland, 62: Department of Evolutionary Biology, Ecology and Environmental Sciences, Faculty of Biology, University of Barcelona, Spain, 63: Departments of Psychiatry and Neuroscience and Physiology, SUNY Upstate Medical University, Syracuse, New York, USA, 64: Department of Genetic Epidemiology in Psychiatry, Central Institute of Mental Health, Medical Faculty Mannheim, University of Heidelberg, Heidelberg, D-68159 Mannheim, Germany, 65: Department of Human Genetics, David Geffen School of Medicine, University of California, Los Angeles, California 90095, USA, 66: Division of Psychiatric Genomics, Department of Psychiatry, Icahn School of Medicine at Mount Sinai, New York, New York 10029, USA, 67: Barnet, Enfield and Haringey Mental Health NHS Trust, St.Ann’s Hospital, St.Ann’s Road, London, N15 3 TH, UK, 68: Departments of Psychiatry and Human Genetics, University of Chicago, Chicago, Illinois 60637, USA, 69: Faculté de Médecine Sorbonne Université, Groupe de Recherche Clinique n°15 - Troubles Psychiatriques et Développement (PSYDEV), Department of Child and Adolescent Psychiatry,Hôpital Universitaire de la Pitié-Salpêtrière, 47-83 Boulevard de l’Hôpital, 75651 Paris Cedex 13, France., 70: Department of Psychiatry and Psychotherapy, Medical University of Vienna, Vienna, Austria, 71: Department of Genetics, University of North Carolina, Chapel Hill, North Carolina 27599-7264, USA, 72: Departments of Psychiatry and Human and Molecular Genetics, INSERM, Institut de Myologie, Hôpital de la Pitiè-Salpêtrière, Paris, 75013, France, 73: Analytic and Translational Genetics Unit, Massachusetts General Hospital, Boston, Massachusetts 02114, USA, 74: Department of Child and Adolescent Psychiatry, Hospital General Universitario Gregorio Marañón, School of Medicine, Universidad Complutense, Investigación Sanitaria del Hospital Gregorio Marañón, Madrid, Spain, 75: BIOARABA Health Research Institute. OSI Araba. University Hospital, University of the Basque Country, Vitoria, Spain, 76: Neuroscience Therapeutic Area, Janssen Research and Development, Titusville, New Jersey 08560, USA, 77: Mater Research Institute, University of Queensland, Brisbane, Queensland 4102, Australia, 78: Department of Psychiatry and Biobehavioral Sciences, Geffen School of Medicine, University of California Los Angeles, 10833 Le Conte Ave, Los Angeles, CA 90095, 79: Department of Psychiatry, University of California San Diego, 9500 Gilman Drive, La Jolla, CA 92093-0804, 80: INSERM U1245, 76000 Rouen, Normandie, France, 81: Department of Psychiatry and Neuropsychology, School for Mental Health and Neuroscience, Maastricht University Medical Centre, Maastricht, the Netherlands, 82: Department of Psychiatry, Faculty of Medicine and Biomedical Research Centre (CIBM), University of Granada, Granada, Spain., 83: Department of Psychiatry, Charité - Universitätsmedizin, Berlin, Campus Benjamin Franklin, Germany, 84: Children's Hospital of Philadelphia, Leonard Madlyn Abramson Research Center,3615 Civic Center Boulevard, Suite 1216, Philadelphia, PA , USA 19104-4318, 85: MRC Human Genetics Unit, University of Edinburgh, Institute of Genetics and Molecular Medicine, Western General Hospital, Edinburgh, UK, 86: School of Medicine and Public Health, University of Newcastle, Newcastle NSW 2308, Australia, 87: Division of Medical Genetics, Department of Biomedicine, University of Basel, Basel, CH-4058, Switzerland, 88: Department of Psychiatry, Fujita Health University School of Medicine, Toyoake, Aichi, 470-1192, Japan, 89: Department of Psychosis Studies, Institute of Psychiatry, Psychology and Neuroscience, King’s College London, London SE5 8AF, UK, 90: Regional Centre for Clinical Research in Psychosis, Department of Psychiatry, Stavanger University Hospital, 4011 Stavanger, Norway, 91: Rheumatology Research Group, Vall d'Hebron Research Institute, Barcelona, 08035, Spain, 92: Roche Pharma Research and Early Development, Pharmaceutical Sciences, Roche Innovation Center Basel, F. Hoffman-La Roche Ltd, Grenzacherstrasse 124, 4070 Basel, Switzerland, 93: Laboratory of Complex Trait Genomics, Department of Computational Biology and Medical Sciences, Graduate School of Frontier Sciences, The University of Tokyo, Tokyo,108-8639, Japan., 94: Department of Medical Genetics, Medical University, Sofia 1431, Bulgaria, 95: Institute for Behavioural Genetics, University of Colorado Boulder, Boulder, Colorado 80309, USA, 96: Institute of Molecular Genetics of National Research Centre “Kurchatov Institute”, Moscow, Russia, 97: Department of Psychiatry, Chonnam National University Medical School, Gwangju, Korea, 98: Latvian Biomedical Research and Study Centre, Riga, LV-1067, Latvia, 99: Mental Health Research Center, Moscow, Russian Federation, 100: RIKEN Center for Integrative Medical Sciences, Yokohama, Kanagawa, 230-0045, Japan, 101: Faculty of Medicine, Vilnius University, LT-01513 Vilnius, Lithuania, 102: Psychiatry Department, University of Indonesia - Cipto Mangunkusumo National General Hospital, Jakarta Pusat, DKI Jakarta, 10430, Indonesia, 103: Department of Psychiatry, 1st Faculty of Medicine and General University Hospital, Prague, Czech Republic, 104: Dipartimento di Biologia, Universita' di Pisa, Pisa, Italy, 105: Departments of Psychiatry and Behavioral Sciences, Stanford University, 450 Serra Mall, Stanford, CA 94305, 106: Psychiatric and Neurodevelopmental Genetics Unit, Department of Psychiatry and Center for Genomic Medicine, Massachusetts General Hospital, Harvard Medical School, Boston, MA, USA, 107: Department of Psychiatry, Wright State University, 3640 Colonel Gleen Hwy, Dayton, OH, 108: Department of Psychiatry, Hadassah-Hebrew University Medical Center, Jerusalem 91120, Israel, 109: Zhongshan School of Medicine and Key Laboratory of Tropical Diseases Control (SYSU), Sun Yat-sen University, Guangzhou 510080, China, 110: Department of Psychiatry, Columbia University, New York, New York 10032, USA, 111: VISN 22, Mental Illness Research, Education & Clinical Center (MIRECC), VA San Diego Healthcare System, 3350 La Jolla Village Drive, San Diego, CA 92161, 112: Department of Psychiatry, National Taiwan University Hospital, Taipei, Taiwan, 113: Mental Health Unit, Department of Public Health Solutions, National Institute for Health and Welfare, PO Box 30, FI-00271, Helsinki, Finland, 114: Hunter New England Health & University of Newcastle, Newcastle NSW 2308, Australia, 115: Department of Genetics and Pathology, International Hereditary Cancer Center, Pomeranian Medical University in Szczecin, 70-453 Szczecin, Poland, 116: Department of Psychiatry, UMC Utrecht Brain Center, University Medical Centre Utrecht, Utrecht University, Utrecht, The Netherlands, 117: Department of Biology and Medical Genetics, 2nd Faculty of Medicine and University Hospital Motol, 150 06 Prague, Czech Republic, 118: Black Dog Institute, University of new South Wales, NSW, Australia, 119: Department of Mental Health, Bloomberg School of Public Health, Johns Hopkins University, Baltimore, Maryland 21205, USA, 120: Department for Neurodegenerative Diseases and Geriatric Psychiatry, University Hospital Bonn, Bonn, Germany , 121: Asfalia Biologics, iPEPS-ICM, Hôpital Universitaire de la Pitié-Salpêtrière, AP-HP, Paris 75013, France, 122: Semel Institute for Neurosciene, University of California, Los Angeles, 11301 Wilshire Blvd, Los Angeles, CA, 123: Hospital Universitari Institut Pere Mata, IISPV, Universitat Rovira i Virgili, CIBERSAM, Reus, Spain, 124: Department of Psychiatry, Dalhousie University, Halifax NS, Canada, 125: VA Boston Health Care System, Brockton, Massachusetts 02301, USA, 126: Centre for Neuroimaging, Cognition and Genomics (NICOG), National University of Ireland Galway, Galway, Ireland., 127: Department of Psychiatry and the Behavioral Sciences, Keck School of Medicine, University of Southern California, Los Angeles, CA, USA, 128: Department of Psychiatry, Dalhousie University, 5850/5980 University Avenue | PO Box 9700, Halifax, Nova Scotia | B3K 6R8, 129: Department of Medical Genetics, University of Pécs, Pécs H-7624, Hungary, 130: Massachusetts Mental Health Center Public Psychiatry Division of the Beth Israel Deaconess Medical Center, Boston, Massachusetts 02114, USA, 131: Estonian Genome Center, Institute of Genomics, University of Tartu, Tartu 50090, Estonia, 132: School of Psychology, University of Newcastle, Newcastle NSW 2308, Australia, 133: Estonian Genome Center, Institute of Genomics, University of Tartu, Tartu 51010, Estonia, 134: First Psychiatric Clinic, Medical University, Sofia 1431, Bulgaria, 135: Department of Pharmacy, University of Oslo, Oslo, Norway, 136: Department of Nursing, Faculty of Health Sciences and Biomedical Research Centre (CIBM), University of Granada, Granada, Spain., 137: Department of Genetics, Faculty of Biological Sciences, Campus of Burjassot, Universidad de Valencia, Valencia, Spain, 138: Department of Psychological Medicine, Institute of Psychiatry, Psychology, and Neuroscience, King's College London, London, SE5 8AF, UK, 139: Departments of Public Health and Preventive Medicine, Family Medicine, and Psychiatry and Behavioral Sciences, State University of New York, Upstate Medical University, Syracuse, NY, 140: Department of Psychiatry, Royal College of Surgeons in Ireland, Dublin, Ireland, 141: Department for Neurosciences, Center for Contextual Psychiatry, KU Leuven, Leuven, Belgium, 142: Cognitive Neuropsychiatry Lab, Department of Psychiatry and Psychotherapy, Philipps Universität Marburg, Marburg, Germany, 143: Department of Psychiatry and Behavioral Sciences, Johns Hopkins University School of Medicine, Baltimore, Maryland 21205, USA, 144: Centre for Public Health, Institute of Clinical Sciences, Queen's University Belfast, Belfast BT12 6AB, UK, 145: Department of Statistics and Applied Probability, University of California at Santa Barbara, Santa Barbara, California, USA., 146: Department of Psychiatry, University of Colorado Denver, Aurora, Colorado 80045, USA, 147: Department of Morphology and Genetics, Universidade Federal de Sao Paulo, Rua Botucatu, 740, Laboratorio de Genetica, CEP 04023-900, Sao Paulo, SP, Brazil, 148: Melbourne Neuropsychiatry Centre, University of Melbourne & Melbourne Health, Melbourne VIC 3053, Australia, 149: Department of Public Health Solutions, Genomics and Biomarkers Unit, National Institute for Health and Welfare, PO Box 30, FI-00271, Helsinki, 150: Queensland Brain Institute, The University of Queensland, Brisbane, QLD 4072, Australia, 151: Department of Psychiatry, University of North Carolina, Chapel Hill, North Carolina 27599-7160, USA, 152: Clinic of Psychiatry and Psychotherapy, Weißer Hirsch, Dresden, Germany, 153: Department of Stem Cell and Regenerative Biology, Harvard University, Cambridge, MA 02138, USA , 154: Molecular Psychiatry Laboratory, Division of Psychiatry, University College London, London WC1E 6JJ, UK, 155: Centre for Genomic and Experimental Medicine, Institute of Genetics and Molecular Medicine, University of Edinburgh, Western General Hospital, Crewe Road, Edinburgh, EH4 2XU, UK, 156: Department of Basic and Clinical Neuroscience, Institute of Psychiatry, Psychology and Neuroscience, King's College London, London SE5 8AF, UK, 157: Oxford Health NHS Foundation Trust, Warneford Hospital, Oxford, UK, 158: Department of Psychiatry and Behavioral Sciences, University of Washington, 2815 Eastlake Ave E #200, Seattle, WA 98102, 159: Huntsman Mental Health Institute, Department of Psychiatry, University of Utah School of Medicine, Salt Lake City, UT, USA, 160: SUNY Upstate Medical University, Syracuse, New York, USA, 161: Department of Psychiatry, Massachusetts General Hospital, Boston, Massachusetts 02114, USA, 162: Department of Psychiatry, Psychosomatics and Psychotherapy, Julius-Maximilians-Universität Würzburg, Wuerzburg, Germany, 163: Maastricht University Medical Center, Department of Psychiatry and Neuropsychology, School for Mental Health and Neuroscience, Maastricht, The Netherlands, 164: Généthon, 1 bis, Rue de l’Internationale, 91000 Evry, France., 165: THL-Finnish Institute for Health and Welfare, P.O BOX 30, Mannerheimintie 166, FI-00271 Helsinki, Finland, 166: Department of Psychiatry, School of Medicine, University of Valencia, Hospital Clínico Universitario de Valencia, Spain, 167: Priority Centre for Brain & Mental Health Research, The University of Newcastle, Mater Hospital, McAuley Centre, Waratah, New South Wales 2298, Australia, 168: Department of Psychiatry, Harvard Medical School, 25 Shattuck St, Boston, MA 02115, 169: Division of Cancer Epidemiology and Genetics, National Cancer Institute, Bethesda, Maryland 20892, USA, 170: West Region, Institute of Mental Health, Singapore, 171: School of Biomedical Sciences, The Chinese University of Hong Kong, Hong Kong, China, 172: Centre for Psychiatry Research, Department of Clinical Neuroscience, Karolinska Institutet & Stockholm Health Care Services, Stockholm Region, SE-171 77 Stockholm, Sweden, 173: School of Social and Health Sciences, Leeds Trinity University, Leeds, 174: NORMENT Centre, Institute of Clinical Medicine, University of Oslo, 0424 Oslo, Norway, 175: Department of Clinical Neurology, Medical University of Vienna, 1090 Wien, Austria, 176: Harvard Medical School Department of Psychiatry at Beth Israel Deaconess Medical Center, 330 Brookline Ave, Boston MA 02215, 177: Lieber Institute for Brain Development, Baltimore, Maryland 21205, USA, 178: Department of Genetic Epidemiology in Psychiatry, Central Institute of Mental Health, Medical Faculty Mannheim, University of Heidelberg, Heidelberg,D-68159 Mannheim, Germany, 179: Department of Medical Genetics, University Medical Centre Utrecht, Universiteitsweg 100, 3584 CG, Utrecht, The Netherlands, 180: Department of Biostatistics, Fielding School of Public Health, University of California Los Angeles, 650 Charles E. Young Dr. South, Los Angeles, CA 90095, 181: Department of Psychiatry, Washington University, St. Louis, Missouri 63110, USA, 182: Department of Psychiatry, Campus Benjamin Franklin, Charité – Universitätsmedizin Berlin, 183: Laboratory for Statistical and Translational Genetics, RIKEN Center for Integrative Medical Sciences, Yokohama, Kanagawa, 230-0045, Japan, 184: Université de Paris, Faculté de médecine, Hôpital Cochin-Tarnier, Paris 75006, France, 185: Department of Neuroscience, Biomedicine and Movement Sciences, Section of Psychiatry, University of Verona, 37134 Verona, Italy, 186: Psychiatry Unit, IRCCS Istituto Centro San Giovanni di Dio Fatebenefratelli, Brescia, Italy, 187: Department of Psychiatry, Faculty of Medicine, Istanbul University, Istanbul, Turkey, 188: Division of Mental Health, St. Olav’s Hospital, Trondheim University Hospital, Trondheim, Norway, 189: KU Leuven, Department of Neurosciences, Center for Clinical Psychiatry, Leuven, Belgium, 190: Department of Psychiatry, Research Unit of Clinical Neuroscience, University of Oulu, Oulu, Finland, 191: Molecular and Cellular Therapeutics, Royal College of Surgeons in Ireland, Dublin 2, Ireland, 192: Department for Psychiatry and Psychotherapy, CCM Charité Universitätsmedizin Berlin, corporate member of Freie Universität Berlin, Humboldt-Universität zu Berlin, and Berlin Institute of Health, Berlin, Germany, 193: Neuropsychiatric Epidemiology Research Unit, School of Population and Global Health, University of Western Australia, Perth, Australia, 194: Sheba Medical Center, Tel Hashomer 52621, Israel, 195: School of Life and Environmental Sciences, University of Sydney, Sydney, NSW 2006, Australia, 196: Department of Psychiatry, GGz Centraal, The Netherlands, 197: Stanley Neurovirology Laboratory, Johns Hopkins University School of Medicine, Baltimore, MD, 21287, USA., 198: Campbell Family Mental Health Research Institute, Centre for Addiction and Mental Health, Toronto, Ontario, M5T 1R8, Canada, 199: Bio-X Institutes, Key Laboratory for the Genetics of Developmental and Neuropsychiatric Disorders, Ministry of Education, Shanghai Jiao Tong University, Shanghai 200030, PR China, 200: Department of Psychiatry, The First Affiliated Hospital of Xi'an Jiaotong University, 277 Yanta West Road, Xi’an 710061, China., 201: Department of Neurology, Medical University of Vienna, Austria, 202: Department of Psychiatry, Ankara University Faculty of Medicine, Ankara, Turkey, 203: Department of Psychiatry, Queens University Kingston, 191 Portsmouth Avenue, Kingston ON Canada K7M 8A6, 204: Department of Psychiatry, University of Iowa Carver College of Medicine, Iowa City, Iowa 52242, USA, 205: School of Nursing, Louisiana State University Health Sciences Center, New Orleans, Louisiana 70112, USA, 206: Department of Psychiatry, University of California San Francisco, San Francisco, California, 94143 USA, 207: Center for Neuropsychiatric Research, National Health Research Institutes, Zhunan Town, Miaoli County, Taiwan, 208: University of Sevilla, CIBERSAM, Sevilla, Spain, 209: Centre for Neuroimaging, Cognition and Genomics (NICOG), National University of Ireland Galway, Galway, Ireland, 210: Discipline of Psychiatry, Adelaide Medical School, University of Adelaide, Adelaide, SA, Australia., 211: Department of Molecular and Translational Medicine, University of Brescia, Brescia, Italy, 212: Neurobiology and Cognitive Science Center, National Taiwan University, Taipei, Taiwan, 213: Centre for Clinical Research in Neuropsychiatry, The University of Western Australia, Perth, WA, Australia, 214: Max Planck Institute of Psychiatry, 80804 Munich, Germany, 215: Menzies Institute for Medical Research, University of Tasmania, 216: Rutgers University, Robert Wood Johnson Medical School, New Brunswick NJ, USA, 217: Institute of Psychiatric Phenomics and Genomics (IPPG), University Hospital, LMU Munich, Munich, Germany, 218: VA Puget Sound Health Care System, 1660 S. Columbian Way, Seattle, WA 98108D WA 98102, 219: College of Public Health, China Medical University, Taichung, Taiwan, 220: State Key Laboratory of Genetic Engineering and Ministry of Education (MOE) Key Laboratory of Contemporary Anthropology, Collaborative Innovation Center of Genetics and Development, Human Phenome Institute, School of Life Sciences, Fudan University, Shanghai, China, 221: Department of Clinical Sciences, Psychiatry, Umeå University, SE-901 87 Umeå, Sweden, 222: Division of Psychiatry, Department of Mental Health Neuroscience, University College London, London WC1E 6BT, UK, 223: Department of Psychiatry, San Cecilio University Hospital, University of Granada, Granada, Spain., 224: Institute of Medical Genetics and Pathology, University Hospital Basel, Schönbeinstrasse 40, CH-4031 Basel, Switzerland, 225: Eli Lilly and Company, Erl Wood Manor, Sunninghill Road, Windlesham, Surrey, GU20 6PH, UK, 226: Neuropsychiatric Genetics Research Group, Department of Psychiatry, Trinity College Dublin, Dublin, Ireland, 227: Department of Cellular, Computational and Integrative Biology, University of Trento, Trento, Italy, 228: Dementia Research Institute, Cardiff University, Hadyn Ellis Building, Maindy Road, Cardiff, CF24 4HQ, 229: Program in Medical and Population Genetics, The Broad Institute of MIT and Harvard, Cambridge, MA, USA, 230: Department of Psychiatry, Phoenix VA Healthcare System, Phoenix AZ, USA, 231: Department of Human Molecular Genetics of the Institute of Biochemistry and Genetics of the Ufa Federal Research Center of the Russian Academy of Sciences. (IBG UFRC RAS) Ufa, 450054, Prospekt Octyabrya, 71, 232: Department of Psychiatry, Psychosomatics and Psychotherapy, University of Würzburg, Margarete-Höppel-Platz 1, 97080, Würzburg, Germany., 233: Psychiatric Genetic Epidemiology and Neurobiology Laboratory (PsychGENe lab), Department of Psychiatry and Behavioral Sciences, SUNY Upstate Medical University, Syracuse, NY, USA, 234: Department of Psychiatry, Sungkyunkwan University School of Medicine, Samsung Medical Center, Seoul, Korea, 235: Department of Psychiatry, Icahn School of Medicine at Mount Sinai, New York NY, 10029, USA, 236: Institute of Biochemistry and Genetics of the Ufa Federal Research Center of the Russian Academy of Sciences (IBG UFRC RAS), Ufa, Russia, 237: Department of Psychiatry and Zilkha Neurogenetics Institute, Keck School of Medicine at University of Southern California, Los Angeles, California 90089, USA, 238: Department of Psychosis, Institute of Mental Health, Singapore, 239: Division of Psychiatry Research, Zucker Hillside Hospital, 75-59 263rd Street, Glen Oaks, NY, 11004, USA, 240: Department of Psychiatry, Stanford University, 401 Quarry Rd., Stanford, CA 94305-5797, USA., 241: Human Genetics, Genome Institute of Singapore, A*STAR, Singapore 138672, Singapore, 242: Roche Pharma Research and Early Development,  Roche Innovation Center Basel, F. Hoffman-La Roche Ltd, Basel, Switzerland, 243: Department of Preventative Medicine, Faculdade de Medicina FMUSP, University of Sao Paulo, Brazil;, 244: Neuropsychiatric Epidemiology Research Unit, School of Population and Global Health M431, University of Western Australia, Perth, Australia 6009, 245: National Institute for Health Research (NIHR) Mental Health Biomedical Research Centre at South London and Maudsley NHS Foundation Trust and King's College London, UK, 246: Department of Psychiatry, University of Pittsburgh, 3811 O'Hara St., Pittsburgh PA 15213, 247: Center for Neurobehavioral Genetics, Semel Institute for Neuroscience and Human Behavior, University of California, Los Angeles, California 90095, USA, 248: Early Clinical Development, Pfizer Worldwide Research and Development, Groton, Connecticut 06340, USA, 249: Institute for Molecular Medicine Finland (FIMM), University of Helsinki, Helsinki, Finland, 250: Department of Biochemistry and Molecular Biology II, Faculty of Pharmacy, University of Granada, Granada, Spain, 251: Department of Psychiatry, School of Medicine, Ankara University, Ankara, Turkey, 252: Faculty of Science, Medicine and Health, School of Chemistry and Molecular Bioscience, University of Wollongong, Wollongong NSW 2522, Australia, 253: Department of Biomedical and Neuromotor Sciences, University of Bologna, Italy., 254: Centre for PanorOmic Sciences, LKS Faculty of Medicine, The University of Hong Kong, 255: Bio-X Institutes, Key Laboratory for the Genetics of Developmental and Neuropsychiatric Disorders (Ministry of Education), Collaborative Innovation Center for Brain Science, Shanghai Jiao Tong University, Shanghai, China., 256: Institute of Medical Sciences, University of Aberdeen, Aberdeen, AB25 2ZD, UK, 257: Center for Behavioral Genomics; Department of Psychiatry; University of California, San Diego; La Jolla, CA 92039; U.S.A., 258: University Medical Center Utrecht, Department of Psychiatry, PO BOX 85500, 3584 GA Utrecht, The Netherlands, 259: Department of Psychiatry & Human Behavior, School of Medicine, University of California, Irvine CA 92697-4260, USA, 260: Institute of Biological Psychiatry, Mental Health Services, Copenhagen University Hospital, Copenhagen, Denmark, 261: School of Psychiatry and Clinical Neurosciences, The University of Western Australia, Perth WA 6009, Australia, 262: Peking University Sixth Hospital, Peking University Institute of Mental Health, Beijing, 100191, China;, 263: Department of Psychiatry, Pamela Sklar Division of Psychiatric Genomics, Friedman Brain Institute, Department of Genetics and Genomic Science and Institute for Data Science and Genomic Technology, Icahn School of Medicine at Mount Sinai, New York, NY, 10029, USA, 264: Department of Functional Genomics, Center for Neurogenomics and Cognitive Research, Neuroscience Campus Amsterdam, VU University, Amsterdam 1081, The Netherlands, 265: Institute for Molecular Bioscience, The University of Queensland, Brisbane, QLD 4067, Australia, 266: Department of Medical Epidemiology and Biostatistics, Karolinska Institutet, Stockholm, Sweden, 267: Institute for Genomic Health, SUNY Downstate Medical Center, Brooklyn, NY, USA, 268: Research Division, Institute of Mental Health, Singapore, Republic of Singapore, 269: Division of Mental Health and Addiction, Oslo University Hospital, 0424 Oslo, Norway, 270: Department of Biomedicine and Centre for Integrative Sequencing (iSEQ), Aarhus University, Aarhus, Denmark., 271: Bio-X Institutes, Key Laboratory for the Genetics of Developmental and Neuropsychiatric Disorders (Ministry of Education), Collaborative Innovation Centre for Brain Science, Shanghai Jiao Tong University, 272: Department of Psychiatric Research, Diakonhjemmet Hospital, Oslo, Norway., 273: Stanley Center for Psychiatric Research, Broad Institute of MIT and Harvard, Cambridge, MA, 274: Department of Epidemiology, University Medical Center Groningen, Unviersity of Groningen, Groningen, The Netherlands, 275: Department of Neuroscience, Dokuz Eylül Univ Graduate School of Health Sciences, 276: UniSA Allied Health & Human Performance, University of South Australia, Adelaide, Australia, 277: Instituto de Investigación Sanitaria del Principado de Asturias (ISPA), Asturias, Spain, 278: Laboratory of Integrative Neuroscience, Universidade Federal de Sao Paulo, Rua Pedro de Toledo, 669 - 3° andar fundos, CEP 04039-032, Sao Paulo, SP, Brazil, 279: University of Groningen, Department of Clinical and Developmental Neuropsychology, Groningen, The Netherlands, 280: Center for Neonatal Screening, Department for Congenital Disorders, Statens Serum Institut, Copenhagen, Denmark, 281: Altrecht, General Menthal Health Care, Utrecht, The Netherlands, 282: Hunter Medical Research Institute, Newcastle, New South Wales, Australia, 283: Department of Psychiatry, Monash University, Melbourne, Australia, 284: St Vincent's Hospital, 41 Victoria Parade, Fitzroy, Victoria3065, Australia., 285: School of Medicine, University of Queensland, Herston, QLD Australia, 286: Department of Psychology, University of Chinese Academy of Sciences, Beijing, China, 287: Department of Psychiatry, McGill University, Montreal, Canada, 288: Saw Swee Hock School of Public Health, University of Singapore, Singapore, 289: Centre de Référence des Maladies Rares à Expression Psychiatrique, Department of Child and Adolescent Psychiatry, AP-HP Sorbonne Université, Hôpital Universitaire de la Pitié-Salpêtrière, 47 - 83 Boulevard de l’Hôpital, 75651 Paris Cedex 13, France, 290: Centre de Référence des Maladies Rares à Expression Psychiatrique, Department of Child and Adolescent Psychiatry, AP-HP Sorbonne Université, Hôpital Universitaire de la Pitié-Salpêtrière, 47 - 83 Boulevard de l’Hôpital, 75651 Paris Cedex 13, France;, 291: National Institute for Health Research (NIHR) Maudsley Biomedical Research Centre at South London and Maudsley NHS Foundation Trust and King's College London, UK, 292: Arkin, Institute for Mental Health, Amsterdam, The Netherlands, 293: Department of Psychiatry, Harvard Medical School, Boston, Massachusetts 02115, USA, 294: APC Microbiome Ireland, University College Cork, Cork, Ireland, 295: Department of Medical Genetics, Oslo University Hospital, 0424 Oslo, Norway, 296: Department of Psychiatry and Behavioral Neurosciences, The University of Chicago, Chicago, IL 60637, USA., 297: Folkhälsan Research Center, Helsinki, Finland, 298: Centro de Investigación Biomédica en Red en Salud Mental (CIBERSAM), Madrid, Spain, 299: Centre for Human Genetics, University of Marburg, D-35033 Marburg, Germany, 300: Department of Psychiatry and Biobehavioral Sciences, University of California, Los Angeles, Los Angeles, CA, USA, 301: Centro de Investigación Biomédica en Red de Salud Mental, Spain (CIBERSAM), 302: Institute for Molecular Biology, University of Queendland, Brisbane, Queensland 4072, Australia, 303: VA Greater Los Angeles Healthcare System, 11301 Wilshire Boulevard, Los Angeles, CA90073, 304: Centre Hospitalier du Rouvray, Rouen 76000 France, 305: Department of Psychiatry, Yale School of Medicine, New Haven, CT, USA, 306: Department of Neuroscience, Icahn School of Medicine at Mount Sinai, New York, New York 10029, USA, 307: NorthWestern Mental Health, Melbourne, Vic, Australia, 308: Broad Institute of MIT and Harvard, Cambridge, Massachusetts, USA, 309: Laboratory for Statistical Analysis, RIKEN Center for Integrative Medical Sciences, Yokohama, Kanagawa, 230-0045, Japan, 310: Department of Genetics, Faculty of Biology, Sofia University "St. Kliment Ohridski", Sofia 1164, Bulgaria, 311: Berlin School of Mind and Brain, Humboldt-Universität zu Berlin, Berlin, Germany, 312: Department of Biomedical Data Science, Stanford University, CA, 313: Department of Psychiatry, University of Helsinki, PO Box 22, FI-00014, Helsinki, Finland, 314: Department of Translational Neuroscience, UMC Utrecht Brain Center, University Medical Center Utrecht, Utrecht University, Utrecht, The Netherlands., 315: Melbourne School of Population and Global health, University of Melbourne, Victoria, Australia, 316: Department of Genetics & Genomics, Icahn School of Medicine at Mount Sinai, Mount Sinai, NY, USA, 317: Stanley Center for Psychiatric Research, the Broad Institute of MIT and Harvard, Cambridge, MA, USA, 318: Department of Community Health and Epidemiology, Dalhousie University, Halifax Nova Scotia, Canada, 319: Deceased, 320: Queensland Brain Institute, University of Queensland, St Lucia, Queensland, Australia, 321: College of Medicine, SUNY Downstate Health Sciences University, Brooklyn, NY, USA, 322: Department of Biomedicine, Aarhus University, DK-8000 Aarhus C, Denmark, 323: Department of Psychiatry, Harvard Medical School, Boston MA, USA, 324: Center for Psychopharmacology, Diakonhjemmet Hospital, Oslo, Norway, 325: Institut de Biologia Evolutiva (UPF-CSIC), Departament de Ciències Experimentals i de la Salut, Universitat Pompeu Fabra, PRBB, Barcelona, Spain, 326: Department of Psychiatry and Psychotherapy, Jena University Hospital, 07743 Jena, Germany, 327: Computational Research Division, Lawrence Berkeley National Laboratory, Berkeley, California, USA, 328: The Florey Institute of Neuroscience and Mental Health, The University of Melbourne, Parkville, VIC, Australia, 329: Department of Psychiatry and SleepWell Research Program, Faculty of medicine, University of Helsinki and Helsinki University Central Hospital, 330: Queensland Centre for Mental Health Research, The University of Queensland, Brisbane, QLD, Australia, 331: The Stanley Center for Psychiatric Research, The Broad Institute, Cambridge, MA 02142, USA, 332: Department of Psychiatry, University of Oxford, Oxford, OX3 7JX, UK, 333: VA Puget Sound Health Care System, 1660 S. Columbian Way, Seattle, WA 98108, 334: Hunter Medical Research Institute, New Lambton Heights, New South Wales 2305, Australia, 335: Division of Molecular Medicine, NSW Health Pathology North, Newcastle, NSW 2305, Australia, 336: Massachusetts Mental Health Center Public Psychiatry Division of the Beth Israel Deaconess Medical Center, 330 Brookline Ave, Boston, MA 02215, 337: James J. Peters VA Medical Center, 130 W Kingsbridge Road, Bronx, NY 10468, 338: Yoo Loo Lin School of Medicine, National University of Singapore, 339: Department of Psychiatry, The Chinese University of Hong Kong, Hong Kong, China, 340: TIPS - Network for Clinical Research in Psychosis; Stavanger University Hospital, Stavanger, Norway, 341: Massachusetts Mental Health Center, 75 Fenwood Road, MA 02115, 342: Saw Swee Hock School of Public Health, National University of Singapore, 343: Berlin Institute of Health (BIH), 10178 Berlin, Germany, 344: INSERM U1266, Institut de psychiatrie et de neurosciences, Paris, France, 345: Bulgarian Academy of Science, 346: Department of Mental Health, Norwegian University of Science and Technology, Trondheim, Norway, 347: Medical Research Center Oulu, Oulu University Hospital and University of Oulu, Oulu, Finland, 348: Centre for Clinical Research in Neuropsychiatry, Division of Psychiatry, Medical School, University of Western Austral, 349: Department of Psychiatry, University of Toronto, Toronto, ON, Canada, 350: Center for Translational Medicine, The First Affiliated Hospital of Xi'an Jiaotong University, 277 Yanta West Road, Xi’an 710061, China., 351: Institute of Epidemiology and Preventive Medicine, College of Public Health, National Taiwan University, Taipei, Taiwan, 352: Hospital Universitario Virgen del Rocio, Department of Psychiatry, Universidad del Sevilla, Sevilla, Spain, 353: Ramsay Health Care (SA) Mental Health, 354: Genetic Unit, IRCCS Istituto Centro San Giovanni di Dio Fatebenefratelli, Brescia, Italy, 355: Department of Psychiatry, College of Medicine and National Taiwan University Hospital, National Taiwan University, Taipei, Taiwan , 356: Department of Psychiatry, Massachusetts General Hospital, Boston, MA 02114, USA, 357: Psychosis Research Unit, Aarhus University Hospital, Aarhus, Denmark, 358: Munich Cluster for Systems Neurology, Munich, Germany, 359: Mental Health Services in the Capital Region of Denmark, Mental Health Center Copenhagen, University of Copenhagen, Copenhagen, Denmark, 360: Rutgers University, New Jersey Medical School, Newark NJ, USA, 361: Department of Psychiatry and Behavioral Sciences, SUNY Upstate Medical University, Syracuse, NY, USA, 362: Stanley Center for Psychiatric Research, Broad Institute of MIT and Harvard, Cambridge, MA 02138, USA, 363: Medical and Population Genetics, The Broad Institute of MIT and Harvard, Cambridge, MA, USA, 364: Department of Psychiatry and Behavioral Sciences, University of Washington, Seattle,, 365: School of Life Science and Technology, ShanghaiTech University, Shanghai, China, 366: Department of Psychiatry, Melbourne Medical School, The University of Melbourne, Parkville, VIC, Australia, 367: VISN 22, Mental Illness Research, Education & Clinical Center (MIRECC) , VA San Diego Healthcare System, 3350 La Jolla Village Drive, San Diego, CA 92161, 368: Department of Biomedicine, University of Basel, Basel, Switzerland, 369: MRC Centre for Neuropsychiatric Genetics and Genomics, Cardiff University, Hadyn Ellis Building, Cardiff, CF24 4HQ, 370: Estonian Genome Center, Institute of Genomics, University of Tartu, Tartu, 51010, Estonia, 371: Banner-University Medical Center, Phoenix AZ, USA, 372: Federal State Educational Institution of Highest Education Bashkir State Medical University of Public Health Ministry of Russian Federation (BSMU). Lenina, 3. 450008 Ufa, Russia, 373: Department of Stem Cell and Regnerative Biology, Harvard University, 374: NORMENT, Institute of Clinical Medicine, University of Oslo, 0424 Oslo, Norway, 375: Federal State Educational Institution of Highest Education Bashkir State Medical University of Public Health Ministry of Russian Federation (BSMU), Ufa, Russia, 376: Department of Cell Biology, State University of New York, Downstate Medical Center, 450 Clarkson Ave, Brooklyn NY, 377: Centre de Référence des Maladies Rares à Expression Psychiatrique, Department of Child and Adolescent Psychiatry,AP-HP Sorbonne Université, Hôpital Universitaire de la Pitié-Salpêtrière, 47 - 83 Boulevard de l’Hôpital, 75651 Paris Cedex 13, France., 378: Neuroscience and Mental Health, Lee Kong Chian School of Medicine, Nanyang Technological University, Singapore, 379: Institute of Behavioral Science, Feinstein Institutes for Medical Research, 350 Community Drive, Manhasset, NY, 11030, USA, 380: Department of Medicine, Yong Loo Lin School of Medicine, National University of Singapore, Singapore, 381: Centre for Clinical Research in Neuropsychiatry, Division of Psychiatry, Medical School, University of Western Australia, Perth, Australia 6009, 382: Department of Psychosis Studies, Institute of Psychiatry, King’s College London, De Crespigny Park, Denmark Hill, London SE5 8AF, UK, 383: Analytic and Translational Genetics Unit, Department of Medicine, Department of Neurology and Department of Psychiatry Massachusetts General Hospital, Boston, MA, USA., 384: Shanghai Key Laboratory of Psychotic Disorders, Shanghai Mental Health Center, Shanghai Jiao Tong University School of Medicine, Shanghai, China., 385: Institute of Neurosciences, Biomedical Research Center (CIBM), University of Granada, Granada, Spain, 386: Department of Psychiatry and Behavioral Neurosciences, The University of Chicago, Chicago IL, USA, 387: Illawarra Health and Medical Research Institute, Wollongong NSW 2522, Australia, 388: State Key Laboratory of Brain and Cognitive Sciences, LKS Faculty of Medicine, The University of Hong Kong, 389: Affiliated Hospital of Qingdao University and Biomedical Sciences Institute of Qingdao University (Qingdao Branch of SJTU Bio-X Institutes), Qingdao University, Qingdao, China., 390: Institute of Genomic Medicine, University of California, San Diego; La Jolla, CA 92039; U.S.A, 391: King’s College London, King’s Health Partners, Department of Psychosis Studies, Institute of Psychiatry, London, United Kingdom, 392: Department of Clinical Medicine, University of Copenhagen, Copenhagen, Denmark, 393: National Clinical Research Center for Mental Disorders & NHC Key Laboratory of Mental Health (Peking University) & Chinese Academy of Medical Sciences Research Unit (No.2018RU006), Beijing, 100191, China., 394: Mental Illness Research, Education, and Clinical Center (VISN 2 South), James J. Peters VA Medical Center, Bronx, NY, USA, 395: Broad Institute of MIT and Harvard, Cambridge, Massachusetts, USA., 396: Analytic and Translational Genetics Unit, Massachusetts General Hospital, Boston MA 02114, USA, 397: Department of Psychiatry, Veterans Affairs New York Harbor Healthcare System, Brooklyn, NY, USA, 398: Division of Psychiatry Research, Zucker Hillside Hospital, Glen Oaks NY, USA, 399: Center for Bioinformatics, Department of Informatics, University of Oslo, PO box 1080, Blindern, 0316 Oslo, Norway, 400: Center for Genomics and Personalized Medicine, Aarhus, Denmark, 401: Centre for Psychiatry Research, Department of Clinical Neuroscience, Karolinska Institutet & Stockholm Health Care Services, Stockholm County Council, Stockholm, Sweden., 402: South Australian Health and Medical Research Institute, Adelaide, South Australia, Australia, 403: Centro de Investigación Biomédica en Red de Salud Mental, Oviedo, Asturias, Spain, 404: Centre for Brain & Mental Health Research, The University of Newcastle, Callaghan, NSW, Australia, 405: Neuroscience Research Australia, Sydney, Australia, 406: Institut des Systèmes Intelligents et de Robotique (ISIR), CNRS UMR7222, Sorbonne Université, Campus Pierre et Marie Curie, Faculté des Sciences et Ingénierie, Pyramide, Tour 55, Boîte courrier 173, 4 Place Jussieu, 75252 Paris Cedex 05, France, 407: Department of Obstetrics & Gynecology, Yong Loo Lin School of Medicine, National University of Singapore, 408: UFR santé, Université de Rouen Normandie, Rouen, France, 409: Mental Illness Research Clinical and Education Center (MIRECC), JJ Peters VA Medical Center, Bronx, NY 10468, USA, 410: Second opinion outpatient clinic, GGNet mental health, Warnsveld, The Netherlands, 411: Department of Medicine, Harvard Medical School, Boston, MA, USA, 412: Department of Biomedicine and Centre for Integrative Sequencing (iSEQ), Aarhus University, Aarhus, Denmark, 413: Queensland Centre for Mental Health Research, The Park Centre for Mental Health, Queensland, Australia, 414: Biomedical Research Institute INCLIVA, Valencia, Spain, 415: Institute for Molecular Medicine Finland, FIMM, University of Helsinki, P.O. BOX 20 FI-00014, Helsinki, Finland, 416: South London and Maudsley NHS Mental Health Foundation Trust, 417: Hunter Medical Research Institute, Newcastle NSW, Australia, 418: Lee Kong Chian School of Medicine, Nanyang Technological University, Singapore, 419: Northern Adelaide Local Health Network, 420: Department of Health Data Science, University of Liverpool, Liverpool, UK, 421: Department of Psychiatry and Psychotherapy, University Medical Center Göttingen, Göttingen, Germany, 422: Regeneron Genetics Center, 423: Center for Excellence in Animal Evolution and Genetics, Chinese Academy of Sciences, Kunming, China, 424: Institute of Neuroscience and Medicine (INM-1), Research Center Juelich, Juelich, Germany, 425: Department of Psychiatry, Veterans Affairs New York Harbor Healthcare System, Brooklyn, NY, USA., 426: Department of Psychiatry, Zucker School of Medicine at Hofstra/Northwell, 500 Hofstra University, Hempstead, NY, 11549, USA, 427: Department of Psychiatry, Erasmus University Medical Center, Rotterdam, The Netherlands, 428: The Stanley Center for Psychiatric Research and Program in Medical and Population Genetics, The Broad Institute of MIT and Harvard, Cambridge, MA, USA., 429: Department of Psychiatry, LKS Faculty of Medicine, The University of Hong Kong, 430: Center for GeoGenetics, GLOBE Institute, University of Copenhagen, Copenhagen, Denmark, 431: PKU-IDG/McGovern Institute for Brain Research, Peking University, Beijing, 100871, China, 432: Oxford Health NHS Foundation Trust, 433: Institute for Molecular Medicine Finland (FIMM), Helsinki, Finland, 434: Department of Psychiatry, University of North Carolina, Chapel Hill, North Carolina 27599-7264, USA, 435: Stanley Center for Psychiatric Research, Broad Institute of MIT and Harvard, Cambridge MA 02142, USA
